# Egg-derived anti-SARS-CoV-2 immunoglobulin Y (IgY) with broad variant activity as intranasal prophylaxis against COVID-19

**DOI:** 10.1101/2022.01.07.22268914

**Authors:** Lyn R. Frumkin, Michaela Lucas, Curtis L. Scribner, Nastassja Ortega-Heinly, Jayden Rogers, Gang Yin, Trevor J Hallam, Alice Yam, Kristin Bedard, Rebecca Begley, Courtney A. Cohen, Catherine V. Badger, Shawn A. Abbasi, John M. Dye, Brian McMillan, Michael Wallach, Traci L. Bricker, Astha Joshi, Adrianus C.M. Boon, Suman Pokhrel, Benjamin R. Kraemer, Lucia Lee, Stephen Kargotich, Mahima Agogiya, Tom St. John, Daria Mochly-Rosen

## Abstract

COVID-19 emergency use authorizations and approvals for vaccines were achieved in record time. However, there remains a need to develop additional safe, effective, easy-to-produce, and inexpensive prevention to reduce the risk of acquiring SARS-CoV-2 infection. This need is due to difficulties in vaccine manufacturing and distribution, vaccine hesitancy, and, critically, the increased prevalence of SARS-CoV-2 variants with greater contagiousness or reduced sensitivity to immunity. Antibodies from eggs of hens (immunoglobulin Y; IgY) that were administered receptor-binding domain (RBD) of the SARS-CoV-2 spike protein were developed as nasal drops to capture the virus on the nasal mucosa. Although initially raised against the 2019 novel coronavirus index strain (2019- nCoV), these anti-SARS-CoV-2 RBD IgY surprisingly had indistinguishable enzyme-linked immunosorbent assay binding against variants of concern that have emerged, including Alpha (B.1.1.7), Beta (B.1.351), Delta (B.1.617.2), and Omicron (B.1.1.529). This is distinct for sera from immunized or convalescent patients. Culture neutralization titers against available Alpha, Beta, and Delta were also indistinguishable from the index SARS-CoV-2 strain. Efforts to develop these IgY for clinical use demonstrated that the intranasal anti-SARS-CoV-2 RBD IgY preparation showed no binding (cross-reactivity) to a variety of human tissues and had an excellent safety profile in rats following 28-day intranasal delivery of the formulated IgY. A double-blind, randomized, placebo-controlled phase 1 study evaluating single-ascending and multiple doses of anti-SARS-CoV-2 RBD IgY administered intranasally for 14 days in 48 healthy adults also demonstrated an excellent safety and tolerability profile, and no evidence of systemic absorption. As these antiviral IgY have broad selectivity against many variants of concern, are fast to produce, and are a low-cost product, their use as prophylaxis to reduce SARS-CoV-2 viral transmission warrants further evaluation. (ClinicalTrials.gov: NCT04567810, https://www.clinicaltrials.gov/ct2/show/NCT04567810)

## 1. INTRODUCTION

As of March 15, 2022, over 456 million persons with coronavirus disease 2019 (COVID-19) had been identified in 222 countries and territories with an estimated 6 million deaths (1). It is estimated that only 57% of the world population has been fully vaccinated (2), a figure dramatically lower than the 70%-80% believed needed to reach herd immunity to stop the pandemic (3, 4).

Furthermore, only 14% of people in low-income countries have received at least one vaccine dose to date (2).

The fast emergency regulatory authorizations and approvals of COVID-19 vaccines in various countries were a critical turning point in slowing the spread of the pandemic. However, there remains a global need to develop additional safe, effective, easy-to-produce, and inexpensive prophylaxis to prevent or reduce the risk of acquiring severe acute respiratory syndrome coronavirus 2 (SARS-CoV-2) infection (5, 6). This need is due in part to the global shortage of essential components necessary for manufacturing the intramuscular mRNA vaccines and the requirement for cold chain storage for distribution. In addition, novel means of prevention are of heightened importance as variants of SARS-CoV-2 such as Delta (B.1.617.20) and Omicron (B.1.1.529) increase contagiousness and evade immunity produced by existing vaccines and previous infection (7–12) and where COVID-19 vaccination is unavailable, especially in resource-poor settings. The emergence of these variants has occurred even in populations with high vaccine uptake and are now the most prevalent COVID-19 strains globally (13).

The main entry route for SARS-CoV-2 is the nasal mucosa, which has high levels of the human angiotensin-converting enzyme 2 (hACE2) receptor that is used by the virus to gain cellular entry (14). Viral binding to the hACE2 receptor is mediated by the spike (S) protein on the surface of the viral envelope (15) for all SARS-CoV-2 variants identified so far; even the highly mutated Omicron variant, with 15 mutations in the receptor-binding domain (RBD) of the S protein, is still dependent on hACE2 for its infectivity (16). Therefore, the nasal mucosa is an excellent site as a critical barrier to reduce SARS-CoV-2 entry; antibodies against the SARS-CoV-2 RBD can compete with viral binding to the hACE2 receptor. In addition, antibodies on epithelial surfaces can greatly inhibit lateral viral motility, agglutinate viral particles, and anchor the virus to the extracellular matrix (17, 18), thus making intranasally administered antibodies a potentially important antiviral strategy. Indeed, intranasal antibody prophylaxis has been an effective means against multiple viral pathogens in humans and veterinary applications, including respiratory tract viruses (18). Thus, covering the nasal mucosa with anti-SARS-CoV-2 antibodies could prevent SARS-CoV-2 infection in naïve individuals and may also reduce viral transmission from an infected individual by reducing levels of active virus.

Because the RBD remains essential for SARS-CoV-2 infection, even for variants of concern, we chose recombinant RBD of the S protein (amino acids 328-533) as the immunogen. We next considered the optimal species to raise anti-SARS-CoV-2 polyclonal antibodies and chose to immunize egg-laying hens to enable fast, low-cost, and high-volume production. Antibodies generated in commercial hens (immunoglobulin Y; IgY) are concentrated in their eggs to 50-100 mg/egg (thus yielding approximately 35 g of IgY per year) within 2-3 weeks following vaccination (19–21). This yield can be up to five times higher (175 g of IgY in one year) when using specific pathogen-free (SPF) hens.

Here, we describe the production of anti-SARS-CoV-2 RBD IgY of the S protein in SPF hens and the characterization of these IgY, including in culture neutralization efficacy against current pathogenic viral variants and a phase 1 study that evaluated the safety, tolerability, and pharmacokinetics of anti-SARS-CoV-2 RBD IgY given by intranasal drops in healthy humans.

## 2. MATERIALS AND METHODS

### 2.1 Study Design

The experiments were conducted in 4 parts: 1) production of immunogen (recombinant RBD), immunization of 12 SPF hens, IgY collection from egg yolks, and in vitro characterization of the IgY anti-SARS CoV-2 RBD, 2) Good Laboratory Practice (GLP) blinded safety studies in rat, treated intranasally twice daily for 28 days with a total of 16 mg/kg IgY or vehicle, 3) a preliminary efficacy study of hamsters treated with IgY or phosphate-buffered saline (PBS) for 4 hours before viral challenge, and 4) a placebo-controlled, double-blind phase 1 safety, tolerability, and pharmacokinetic (PK) study conducted in healthy humans using intranasal IgY or vehicle in single-ascending doses followed by multiple doses (3-times daily every 4 hours) for 14 days. Both the single-ascending and multiple-dose parts were followed by a 7-day nontreatment period to further evaluate safety.

### 2.2 Recombinant SARS-CoV-2 RBD

SARS-CoV-2 RBD (residues 328-533) of the 2019 novel coronavirus index virus (2019- nCoV) was expressed in cell-free protein synthesis reactions at Sutro Biopharma, Inc. (South San Francisco, CA) using the XpressCFTM platform (22, 23) and was constructed as a carboxy-terminal fusion to a small ubiquitin-related modifier (SUMO) sequence to enable the production of a tagless RBD protein post enzymatic cleavage. The his-SUMO tag was constructed as previously described with an N-terminal his6-tag followed by a GGS-linker and the yeast SUMO protein Smt3 for Ulp1- enabled cleavage (22, 23).

Briefly, cell-free reactions were prepared by the addition of 37.5% v/v iodoacetamide-treated S30 extract, 5 µg/mL plasmid, and a supermix containing amino acids, nano-microspheres, and small molecules for energy generation (23). T7 RNA polymerase was over-expressed in Escherichia coli and added to the cell-free reaction as a reagent lysate at <1% v/v. Reactions were carried out in a DASbox stirred tank (Eppendorf) at 250 mL volume with pH, dissolved oxygen, and temperature control. Reactions were run for 16 hours at a temperature of 25°C, pH was controlled at 7.0 using 1 M citrate and 1 M potassium hydroxide, and dissolved oxygen was maintained at 20%.

The XpressCF reaction of SARS-CoV-2-RBD 328-533 was clarified by centrifugation at 10,000 rpm for 20 minutes (Beckman, JLA-10.500 rotor) and filtered through a 0.22-μm membrane filter. The clarified material was loaded onto a 5-mL his-Trap Excel affinity column equilibrated with binding buffer (15 mM Tris-HCl, 500 mM NaCl, pH 8.0). After 20 column volumes were applied to wash unbound impurities, the bound proteins were eluted with 20 mM Tris-HCl, 300 mM imidazole, pH 8.0. The eluted fractions were then analyzed by 4-12% sodium dodecyl sulfate-polyacrylamide gel electrophoresis (SDS-PAGE) and protein concentrations were determined by measured absorbance at 280 nm by NanoDrop (Thermo Fisher Scientific). The his-SUMO tag was removed by Ulp1 protease digestion for 1 hour at room temperature. The digested reaction was analyzed by 4- 12% SDS-PAGE to verify full cleavage of the his-SUMO tag before flow-through mode purification by Capto Q (Cytiva). Twenty mM Tris-HCl, pH 8.0, was used to equilibrate the column, and the flow-through containing the cleaved protein was collected. To further purify the CoV-2-RBD, the Capto Q flow-through fraction was bound to a Capto SP ImpRes cation column equilibrated with 20 mM Tris-HCl, pH 8.0. The bound protein was eluted with a 30-column volume linear gradient using elution buffer (20 mM Tris-HCl, 300 mM NaCl, pH 8.0). An Amicon Ultra-15, 3kD centrifugal filter was used to concentrate and buffer exchange the target peak fraction into 6% sucrose in PBS at pH 7.2. The final purity of the product was demonstrated by high-performance liquid chromatography (HPLC) analysis.

#### 2.2.1. Characterization of the cell-free expressed recombinant RBD using surface plasmon resonance

Binding kinetic of cell-free expressed SARS-CoV-2 RBD construct or mammalian expressed his-tagged RBD control (ACROBiosystems SPD-C52H1) were then measured on a Biacore T200 instrument, using Fc-tagged hACE2 receptor protein (ACROBiosystems AC2-H5257). To this end, an anti-human Fc antibody (Jackson ImmunoResearch Labs) was immobilized on all flow cells of a CM5 chip (GE Healthcare). Fc-tagged human ACE2 receptor protein (ACROBiosystems AC2-H5257) was captured at ∼100 replication units (RU). Binding of cell-free expressed SARS-CoV-2 S protein RBD construct or mammalian expressed RBD control (ACROBiosystems SPD-C52H1) was measured at concentrations up to 100 nM. Kinetic experiments were performed at 25°C at a flow rate of 50 μL/min. RBD samples were diluted in HBS-EP buffer (Teknova) and injected over the chip for 180 seconds followed by a 420-second dissociation. The chip was regenerated with 10 mM glycine pH 1.5 after each injection. Affinities were calculated using Biacore T200 Evaluation Software.

### 2.3 Hen immunization and IgY purification and characterization

Nine SPF hens were obtained from Charles River Laboratories and housed in a filtered air, positive pressure barrier room (3 more hens from the same lot were added to the study after 5 months) at Avian Vaccine Services, Charles River Laboratories (Storrs, CT). Each hen was caged individually with access to feed and water. Upon receipt, hens were immunized with an inoculum containing 50 µg of recombinant cell-free expressed RBD fragment derived from S1 spike protein and water-in-oil adjuvant. Test bleeds were taken before the first immunization and every 2 weeks after the first immunization throughout the project. Serum samples from each hen were tested using an indirect enzyme-linked immunoassay (ELISA) and Western blot. Hens received a boost 14 days after immunization and every 4 weeks after the previous immunization, unless otherwise indicated.

Eggs were collected weekly. Yolks in batches up to 100 eggs/batch were separated and IgY was purified. ELISA titration of IgY binding to full-length S1 (ACROBiosystems, S1N-C5255) expressed in human 293 cells and the cell-free expressed RBD of the SARS-CoV-2 spike protein was performed in 96-well plates. IgY was then purified from yolks. Eggs were collected weekly and egg white was separated from the egg yolk and discarded. Yolks were stored frozen at -20°C. For purification of IgY, yolks were thawed and diluted 1:10 in sterile PBS. The pH was reduced to 5.0 and the yolk solution was incubated at 4°C overnight. Yolk solution was then centrifuged for 10 minutes at 9000 rpm. Supernatant was removed, pooled, pH was increased to 7.0, and final filtration (0.45 µm) was performed.

Hen sera and purified IgY were tested by ELISA and Western blot analysis using both the immunogen (RBD) and full length glycosylated S1 protein (ACROBiosystems, cat# S1N-C5255). Lot Y0120 of purified IgY was derived from 50 yolks, starting 2 weeks after the primary immunization. Lot Y0130 was derived from the next 50 yolks, starting 3 weeks after the primary immunization. Lot Y0140 and above were derived from the next 100 yolks collected over the preceding 2 weeks. Lots of purified IgY were tested by indirect ELISA and Western blot to determine antibody response levels within the yolk after purification as well as affinity to the cell-free expressed RBD fragment of S1 spike protein and the entire glycosylated S1 protein (ACROBiosystems, cat# S1N-C5255). Protein concentration was also measured using the Bradford assay. IgY purity was determined using SDS-PAGE under reducing conditions and stained with Coomassie Blue.

ELISA titration of IgY binding to full length and the RBD of the SARS-CoV-2 spike protein was performed as follows: 96-well clear flat bottom polystyrene high-binding ELISA plates (Corning, Cat# 9018) were coated with 100 μl/well of 5 μg/mL recombinant his-tagged (Sino Biological) RBD protein overnight at 4°C. Plates were then washed twice with PBS and blocked with 1% bovine serum albumin (BSA)-PBS. After blocking, 100 μL of serial dilutions of IgY was added to the wells and incubated at room temperature for 2 hours. The plates were then washed twice and incubated with horseradish peroxidase (HRP)-conjugated mouse rat anti-chicken IgY (Sapphire, Cat# LO-IgY-16-P-0.5 ) 1:2000 diluted in 0.5% BSA and 0.05% Tween-20 in PBS) for 60 minutes. After 3 final washes using 0.05% PBS-Tween, the plates were read using chemiluminescent substrate (Pierce TMB Substrate Kit, Thermo Fisher, Cat# 34021). Luminescence was read on a plate reader and analyzed with SoftMaxPro 6.0 (Molecular Devices).

Hen sera were diluted with 1% BSA at various dilutions and tested in ELISA. One hundred microliters were added to wells and the plate was incubated at room temperature for 1 hour. Plate wells were then rinsed with 1% Tween 20 wash solution, as indicated above. Anti-chicken HRP conjugate (Invitrogen, cat# SA-19509) diluted 1:5000 was added to each well. The plate was incubated at room temperature for 1 hour, followed by another 1% Tween 20 wash. 3,3′,5,5′- tetramethylbenzidine substrate (KPL, cat# 5120-0050) was added to each well and the plate was incubated at room temperature for 15 minutes. Optical density was measured using an ELISA reader (Molecular Devices) at 650 nm.

Western blot analysis was performed with the cell-free expressed RBD fragment of the index S1 protein or the entire mammalian-expressed S1 protein (ACROBiosystems). One microgram of denatured protein was added to Mini-PROTEAN TGX 4-20% SDS-PAGE. Gel was run at 200V for 45 minutes in Tris-glycine buffer and stained using Simply Blue Safestain (Invitrogen) at room temperature for 1 hour. Gels were destained twice using deionized water.

An unstained SDS-PAGE gel was transferred to the polyvinylidene difluoride membrane using Trans-Blot Turbo Transfer System (Bio-Rad Laboratories) and cut into strips. Strips were then stored at -20°C until ready to use. Five percent nonfat dry milk was added to strips and incubated at room temperature for 30-60 minutes. Test bleed sera and IgY were diluted 1:2000 and purified IgY was diluted 1:500 in 0.2 µm-filtered PBS and added to strips. Strips were incubated at room temperature for 2 hours followed by 3 washes in filtered PBS. Rabbit anti-chicken IgY (H+L) secondary antibody, HRP (Invitrogen) diluted 1:3000 was added to each strip and incubated at room temperature for 1-2 hours. Strips were washed once again in filtered PBS. Opti-4CN substrate (Bio-Rad Laboratories) was added to each strip and incubated at room temperature for up to 30 minutes in a rocking incubator. Strips were washed twice in deionized water and images were taken using Bio-Rad EZ Gel Imager.

### 2.4 Evaluation of IgY

#### 2.4.1. ELISA evaluation of IgY titer against SARS-CoV-2 variants of concern

ELISA titer of the final IgY preparation used in the clinical studies against the Alpha, Beta, Delta, and Omicron-derived RBD [amino acids 319-537; ACROBiosystems SPD-C52H1(index), SPD-C52Hn (Alpha), SPD-C52Hp (Beta), SPD-C525e (Delta), and SPD-C522e (Omicron)]) was carried out as described above.

#### 2.4.2 In culture viral neutralization studies using pseudovirus

The neutralization assays using pseudovirus were performed at RetroVirox (San Diego, CA).

The assay used 3 non-replicative vesicular stomatitis virus (VSV) pseudoviruses carrying a firefly luciferase reporter gene and expressing the following S protein of SARS-CoV-2 on the surface: the index SARS-CoV-2 Hu-1 spike, a truncated spike with a C-terminal 19 amino acid deletion; the Beta variant (K417N/ E484K / N501Y / D614G full-length spike); and a D614G spike variant with a full-length sequence of the index spike protein. The IgY preparation or plasma was diluted in Dulbecco’s Modified Eagle Medium (DMEM) with 5% fetal bovine serum (FBS) to perform pre-incubation with pseudovirus before addition to target cells. The neutralization assay was performed with HEK 293T- hACE2, a human embryonic kidney cell line overexpressing hACE2. Test item was preincubated for 60 minutes at 37°C with a previously titrated inoculum of pseudovirus. The mixture was then added to 293T-hACE2 cells and infection was allowed for 24 hours. Pseudovirus infection was determined by measuring firefly luciferase activity (RLU, relative light unit ratio) 24 hours after infection.

Putative neutralizing antibodies (human sera, purified IgY, or vehicle) were present in the cell culture for the entire duration of the experiment. The ability of the test item to neutralize pseudovirus carrying SARS-CoV-2 spike was compared with samples treated with vehicle alone. Nine concentrations of the IgY sample were tested in duplicates with each variant in 5-fold serial dilutions starting at 10,000 μg/mL. The half-maximal inhibitory concentration (IC50) and 50% neutralization titer (NT50) values for the IgY and positive control human serum were determined using GraphPad Prism software (GraphPad Software).

Quality controls for the pseudovirus neutralization assays were performed to determine: 1) signal-to-background values; 2) variation of the assay, estimated as the average of the coefficient of variation for data points for which 50% or greater infection (RLUs) was observed compared to cells with pseudovirus in the presence of vehicle alone, and 3) neutralization with plasma from an individual who received 2 doses of the Moderna (mRNA-1273) COVID-19 vaccine. All controls worked as anticipated for each assay.

#### 2.4.3 In culture neutralization studies using live virus

At the United States Army Medical Research Institute of Infectious Diseases (USAMARIID; Frederick, MD), all work with authentic (live) SARS-CoV-2 was completed in Biosafety Level 3 laboratories following federal and institutional biosafety standards and regulations. Vero-76 cells were inoculated with SARS-CoV-2/Was1 (MT020880.1) at a multiplicity of infection of 0.01 and incubated at 37°C with 5% CO2 and 80% humidity. At 50 hours post-infection, cells were frozen at - 80°C for 1 hour, allowed to thaw at room temperature, and supernatants were collected and clarified by centrifugation at ∼2,500 ×g for 10 minutes. Clarified supernatant was aliquoted and stored at - 80°C.

Authentic (live) SARS-CoV-2/B.1.617.2 at a multiplicity of infection of 1 was incubated for 1 hour at 37°C with serially diluted antibodies. Vero-E6 monolayers were exposed to the antibody-virus mixture at 37°C for 1 hour. Following incubation, viral inoculum was removed and fresh cell culture media was added for an additional 23 hours at 37°C. Cells were washed with PBS, fixed in 10% formalin, and permeabilized with 0.2% Triton-X for 10 minutes. Detection of infection was accomplished using an anti-SARS-CoV-2 nucleocapsid protein detection antibody (Sino Biological), and a goat α-rabbit secondary antibody conjugated to AlexaFluor488. Infected cells were identified using the PerkinElmer Operetta high-content imaging instrument and data analysis was performed using the Harmony software (PerkinElmer).

At RetroVirox (San Diego, CA), purified IgY (lot Y0180) was tested against three live SARS-CoV-2 clinical isolates: MEX-BC15/2021 (lineage B.1.617.2, Delta), USA/SD-RVX01/2022 (lineage B.1.529, Omicron) and MEX-BC2/2020 (lineage B.1, carrying the D614G mutation). A virus-induced cytopathic effect (CPE)-based neutralization assay was performed by infecting Vero E6 cells in the presence or absence of test items. Vero E6 cells were maintained in DMEM with 10% FBS. Twenty-four hours after cell seeding, test samples were submitted to serial dilutions with DMEM with 2% FBS in a different plate. Then, virus diluted in the same media alone was pre-incubated with test items for 1 hour at 37°C in a humidified incubator. Following incubation, media was removed from cells. Cells were then challenged with the SARS-CoV-2/antibody pre-incubated mix. The amount of viral inoculum was previously titrated to result in a linear response inhibited by antivirals with known activity against SARS-CoV-2. Cell culture media with the virus inoculum was not removed after virus adsorption, and antibodies and virus were maintained in the media for the duration of the assay (96 hours). After this period, the extent of cell viability was monitored with the neutral red uptake assay.

The virus-induced CPE was monitored under the microscope after 3 days of infection, and cells were stained with neutral red to monitor cell viability the following day. Viable cells incorporate neutral red in their lysosomes. The uptake of neutral red relies on the ability of live cells to maintain the pH inside the lysosomes lower than in the cytoplasm, a process that requires ATP. Inside the lysosome, the dye becomes charged and is retained. After a 3-hour incubation with neutral red (0.017%), the extra dye is washed away, and the neutral red is extracted from lysosomes by incubating cells for 15 minutes with a solution containing 50% ethanol and 1% acetic acid. The amount of neutral red is estimated by measuring absorbance at 540 nm in a plate reader.

Antibodies were evaluated in triplicates using five-fold serial dilutions starting at 10.0 mg/mL. Controls included uninfected cells and infected cells treated with vehicle alone. Some cells were treated with plasma from an uninfected individual who received two doses of Moderna’s mRNA COVID-19 vaccine as a positive control.

#### 2.4.4 CPE-based neutralization assay

The average absorbance at 540 nm (A540) observed in infected cells in the presence of vehicle alone was calculated, and then subtracted from all samples to determine the inhibition of the virus-induced CPE. Data points were then normalized to the average A540 signal observed in uninfected cells after subtraction of the absorbance signal observed in infected cells. In the neutral red CPE-based neutralization assay, uninfected cells remained viable and uptake the dye at higher levels than non-viable cells. In the absence of test items, the virus-induced CPE kills infected cells and leads to lower A540 (this value equals 0% inhibition). In contrast, incubation with neutralizing agents prevents the virus-induced CPE and leads to absorbance levels similar to those observed in uninfected cells. Full recovery of cell viability in infected cells represents 100% neutralization of the virus.

### 2.5 GMP IgY formulation, analytical studies, and stability

Using Good Manufacturing Practice (GMP), anti-S1 RBD IgY was formulated for use as intranasal drops as 0 (placebo control), 5, 10, and 20 mg/mL anti-S1 RBD IgY preparations in sterile 2% microcrystalline cellulose and carboxymethylcellulose sodium at Bravado Pharmaceuticals (Lutz, FL). The suspension was packed in a 1.5-mL dropper bottle and tests for GMP drug product release complied with the relevant standards and methods, including microbiological examination of nonsterile products (USP <1111>, <61> and <62>). All formulated products were 100% stable as measured by physical and analytical properties, including HPLC, when stored for at least 6 months at 2-8°C and about 1 month when stored at room temperature. Ten and 20 mg/mL solutions stored at room temperature showed 100% analytical stabilities for 3 months. However, several samples had some physical visual abnormalities (slight discoloration and opacity) that occurred beginning at 1 month when stored at room temperature. Therefore, all formulated samples used in the subsequent studies were stored at 2-8°C. For stability determination, formulated IgY (20 mL) was chromatographed on a TSK Gel G3000swxl sizing column (with guard column) on HPLC at 0.4 mL/min at room temperature, and eluate was monitored at 214 nm. For the rat toxicity and safety study, and for the human study, we used only 20 mg/mL as the highest dose because it was readily soluble.

### 2.6 GLP rat toxicity and safety study

Thirty-eight female and 38 male >8-week-old Sprague Dawley rats were used in a GLP study conducted at Charles River Laboratories (Spencerville, OH). Rats were identified using a subcutaneously implanted electronic identification chip and were acclimated to their housing for at least 4 days before the first day of dosing. Animals were randomly assigned to groups; males and females were randomized separately and housed in accordance with the United States Department of Agriculture (USDA) Animal Welfare Act (9 CFR, Parts 1, 2, and 3) and as described in the Guide for the Care and Use of Laboratory Animals.

For psychological and environmental enrichment, a hiding device, a chewing object, and edible enrichment treats were offered throughout the study and a cycle of 12 hours light and 12 hours dark was maintained. Water and food were freely available. The experimental protocol is summarized in **Supplementary Tables S1-S3**.

Detailed clinical observation was conducted once a week and included body weight and food consumption. Ophthalmic examinations were conducted before treatment and during the last week of dosing by a veterinary ophthalmologist and included a short-acting mydriatic solution treatment to each eye to facilitate the ocular examinations. Clinical laboratory assessments included the presence of circulating IgY (in serum) and standard hematology, coagulation, clinical chemistry, and urinalysis parameters tested at pretreatment and Day 28 (last day of treatment) after 4 hours of fasting.

Hematology parameters included red blood cell count, hemoglobin concentration, hematocrit, mean corpuscular volume, red blood cell distribution width, mean corpuscular hemoglobin concentration, mean corpuscular hemoglobin, reticulocyte count (absolute), platelet count, white blood cell count, neutrophil count (absolute), lymphocyte count (absolute), monocyte count (absolute), eosinophil count (absolute), basophil count (absolute), large unstained cells (absolute) and other cells (as appropriate). Clinical chemistry parameters included alanine aminotransferase, aspartate aminotransferase, alkaline phosphatase, gamma-glutamyltransferase, creatine kinase, total bilirubin, urea nitrogen, creatinine, calcium, phosphorus, total protein, albumin, globulin (calculated), albumin/globulin ratio, glucose, cholesterol, triglycerides, sodium, potassium, chloride, and sample quality. Urinalysis included color, appearance/clarity, specific gravity, volume, pH, protein, glucose, bilirubin, ketones, and blood.

#### 2.6.1 Cytokine level measurement and analysis

This non-GLP blinded assay was performed on serum by the Immunoassay Team at the Human Immune Monitoring Center at Stanford University (Stanford, CA). Assay kits (RECYMAG65K27PMX Rat) were purchased from EMD Millipore and used according to the manufacturer’s recommendations, with modifications described as follows. Briefly, samples were mixed with antibody-linked magnetic beads on a 96-well plate and incubated overnight at 4°C while shaking. Cold and room temperature incubation steps were performed on an orbital shaker at 500-600 rpm. Plates were washed twice with wash buffer in a Biotek ELx405 washer. Following 1-hour incubation at room temperature with a biotinylated detection antibody, streptavidin-PE was added for 30 minutes while shaking. Plates were washed as described above and PBS was added to wells for reading in the Luminex FLEXMAP 3D Instrument with a lower bound of 50 beads per sample per cytokine. Each sample was measured in duplicate. Custom Assay Chex control beads were purchased from Radix BioSolutions and added to all wells.

Median fluorescence intensity data were corrected for plate and nonspecific binding artifacts (23) and regressed on time point. Regression analysis employed linear mixed models (24) in SAS®/STAT (SAS Institute) with a separate model fit for each treatment and cytokine, and with separate sets of models with and without gender as a covariate, for 108 models in total. Of the 108 models, 84% met the assumption of normally distributed residuals and 78% for random effects.

#### 2.6.2 IgY in sera of treated rats

The presence of anti-SARS-CoV-2 IgY in sera of rats was evaluated using GLP standards in a qualified ELISA assay at Charles River Laboratories (Reno, NV). ELISA 96-well plates were coated with goat anti-chicken IgY (Thermo Fisher; A16056). After blocking, the plates were incubated with the samples containing anti-SARS-CoV-2 IgY at various concentrations for 1 hour at room temperature. After washing the microplate, rabbit anti-chicken IgY (Thermo Fisher; A16130) - HRP conjugate was added and incubated for 1 hour at room temperature. The ELISA plate was washed and substrate (3,3’,5,5’-tetramethylbenzidine) was added to the wells and incubated for 20 minutes. The color development was stopped by the addition of 2N sulfuric acid and color intensity measured in a microplate reader at 450 nm. A calibration curve from the absorbance values was obtained from the standards using a 4-parameter curve fit with a weighting equation of 1/y2. The concentrations of anti-SARS-CoV-2 IgY in the samples were determined from the calibration curve. After 4 analytical method validations, lower and upper limit of detection, intra- and inter-assay precision and accuracy, dilution integrity, and short-term stability, IgY levels in the blood samples of rats before and after 28 days of treatment were below the limit of detection.

### 2.7 Human tissue cross-reactivity study

A GLP study examining human tissue reactivity of the anti-SARS-CoV-2 RBD IgY was conducted at Charles River Laboratories (Frederick, MD) using at least 3 tissues from at least 3 donors (**Supplementary Table S4**). Ten mL of 10 mg/mL or 20 mg/mL IgY control (negative control) and anti-SARS-CoV-2 RBD IgY or an anti-human hypercalcemia of malignancy peptide (amino acid residues 1-34, Sigma-Aldrich; Catalog No. H9148; positive control), each in 1% BSA, were incubated for 1 hour with acetone-fixed human tissues (normal) of at least 3 separate donors after 20 minutes incubation with block solution of PBS + 1% BSA, 0.5% casein and 5% normal rabbit serum. After PBS washes, the secondary antibody (peroxidase [HRP]-conjugated rabbit anti-chicken IgY; Testing Facility antibody tracking No. A45764; of 2 μg/mL) was added for 30 minutes. After PBS washes, DAB (3,3′-diaminobenzidine) was applied for 4 minutes as a substrate for the peroxidase reaction. All slides were rinsed with tap water, counterstained, dehydrated, and mounted for visualization. For the β2-microglobulin antibody (positive control staining) 1 μg/mL of antibodies were incubated with the slides for 1 hour and bound antibodies were detected with biotinylated secondary antibody (goat anti-rabbit IgG; 2 μg/mL) for 30 minutes, as above. The slides were then visualized on light microscopy.

### 2.8 Virus preparation for the efficacy study in a hamster model of COVID-19 and quantitation of viral load in the animals

Syrian hamsters develop mild-to-moderate disease with progressive weight loss that starts several days after SARS-CoV-2 infection by intranasal inoculation (25, 26). SARS-CoV-2 (strain 2019- nCoV/USA-WA1/2020) was propagated on Vero-TMPRSS2 cells, and the virus titer was determined by plaque assays on Vero-hACE2 and Vero-hACE2-TMPRSS2 cells. Briefly, cells were seeded in 24-well plates, and the next day, virus stocks were serially diluted 10-fold, starting at 1:10, in cell infection medium [Minimum Essential Media (MEM) containing 2% FBS, L-glutamine, penicillin, and streptomycin]. Two hundred and fifty microliters of the diluted virus were added to a single well per dilution per sample. After 1 hour at 37°C, the inoculum was aspirated and a 1% methylcellulose overlay in MEM supplemented with 2% FBS was added. Seventy-two hours after virus inoculation, the cells were fixed with 4% formalin and the monolayer was stained with crystal violet (0.5% w/v in 25% methanol in water) for 1 hour at 20°C. The number of plaques was counted and used to calculate the plaque-forming units (PFU)/mL.

#### 2.8.1 Study protocol: hamster infection with SARS-CoV-2

Five- to 6-week-old Syrian golden hamsters (Charles River Laboratories) infected with SARS-CoV-2 as previously described (26) were housed at the Washington University (St. Louis, MO) Biosafety Level 3 facility in HEPA-filtered rodent cages. Before challenge with SARS-CoV-2, all animals received 100 μL of a placebo control fluid or fluid-formulated anti-SARS-CoV-2 RBD IgY (1 mg/50 mL of the 20 mg/mL solution per nare). The amount of nasal anti-SARS-CoV-2 RBD IgY drops was limited by the volume that could be given per nare. Four hours after the delivery, animals were challenged with 10^4^ or 5 x 10^4^ PFU of SARS-CoV-2 (titer determined on Vero-hACE2; see below). The protocol included 18 hamsters in 4 planned groups: Group 1: Control with high 5 x 10^4^ PFU (4 animals); Group 2: Control with 1 x 10^4^ PFU (4 animals); Group 3: Anti-SARS-CoV-2 RBD IgY with 5 x 10^4^ PFU (5 animals); and Group 4: Anti-SARS-CoV-2 RBD IgY with 1 x 10^4^ PFU (5 animals). Note that the viral titer in Vero-hACE2-hTMPRSS2 cells, which express the essential protease to liberate the RBD from the S protein on the surface of the virus (27), was subsequently found to be almost 100-fold higher for Groups 1 and 3 (4 x 10^6^ PFU) and Groups 2 and 4 (0.8 x 10^6^ PFU).

Animal weight was recorded daily. Three days after challenge, the animals were sacrificed and lungs were collected. The left lobe was homogenized in 1.0 mL of DMEM and the clarified supernatant was used for viral load analysis by plaque assay and quantitative reverse transcription PCR.

#### 2.8.2 Virus titration assays from hamster samples

Plaque assays were performed on Vero-hACE2 and Vero-hACE2-TMPRSS2 cells in 24-well plates. Lung tissue homogenates were serially diluted 10-fold, starting at 1:10, in cell infection medium (DMEM containing 2% FBS, L-glutamine, penicillin, and streptomycin). Two hundred and fifty microliters of the diluted lung homogenates were added to a single well per dilution per sample.

After 1 hour at 37°C, the inoculum was aspirated and a 1% methylcellulose overlay in MEM supplemented with 2% FBS was added. Seventy-two hours after virus inoculation, the cells were fixed with 4% formalin, and the monolayer was stained with crystal violet (0.5% w/v in 25% methanol in water) for 1 hour at 20°C. The number of plaques was counted and used to calculate the PFU/mL. To quantify viral load in lung tissue homogenates, RNA was extracted from 100 µL samples using QIAamp viral RNA mini kit (Qiagen) and eluted with 50 µL of water. RNA (4 µL) was used for real-time quantitative reverse transcription PCR to detect and quantify N gene of SARS-CoV-2 using TaqMan™ Fast Virus 1-Step Master Mix (Applied Biosystems) or using the following primers and probes for the N-gene, forward: GACCCCAAAATCAGCGAAAT; reverse: TCTGGTTACTGCCAGTTGAATCTG, probe: ACCCCGCATTACGTTTGGTGGACC, or the 5’- UTR, forward: ACTGTCGTTGACAGGACACG, reverse: AACACGGACGAAACCGTAAG, probe: CGTCTATCTTCTGCAGGCTG. Viral RNA was expressed as gene copy numbers per mg for lung tissue homogenates and per mL for nasal swabs, based on a standard included in the assay that was created by in vitro transcription of a synthetic DNA molecule containing the target region of the N gene or 5’-UTR.

### 2.9 Statistical Analysis

Data were analyzed with GraphPad Prism 9.0 and statistical significance was assigned when P values were < 0.05. All tests and values are indicated in figure legends.

### 2.10 Phase 1 safety, tolerability, and pharmacokinetic study in humans

#### 2.10.1 Study design and dose selection

A single-center, randomized, double-blind, placebo-controlled phase 1 study of anti-SARS-CoV-2 RBD IgY given intranasally to 48 healthy adults was conducted at Linear Clinical Research-Harry Perkins Research Institute (Nedlans, WA, Australia) (see **Figure 1**). Written informed consent was obtained from all participants. The study is registered at ClinicalTrials.gov: NCT04567810. The study period was conducted between September 25, 2020 and December 14, 2020. The protocol for this trial is available as supporting information (**Supplementary Methods)**.

**Fig. 1.**
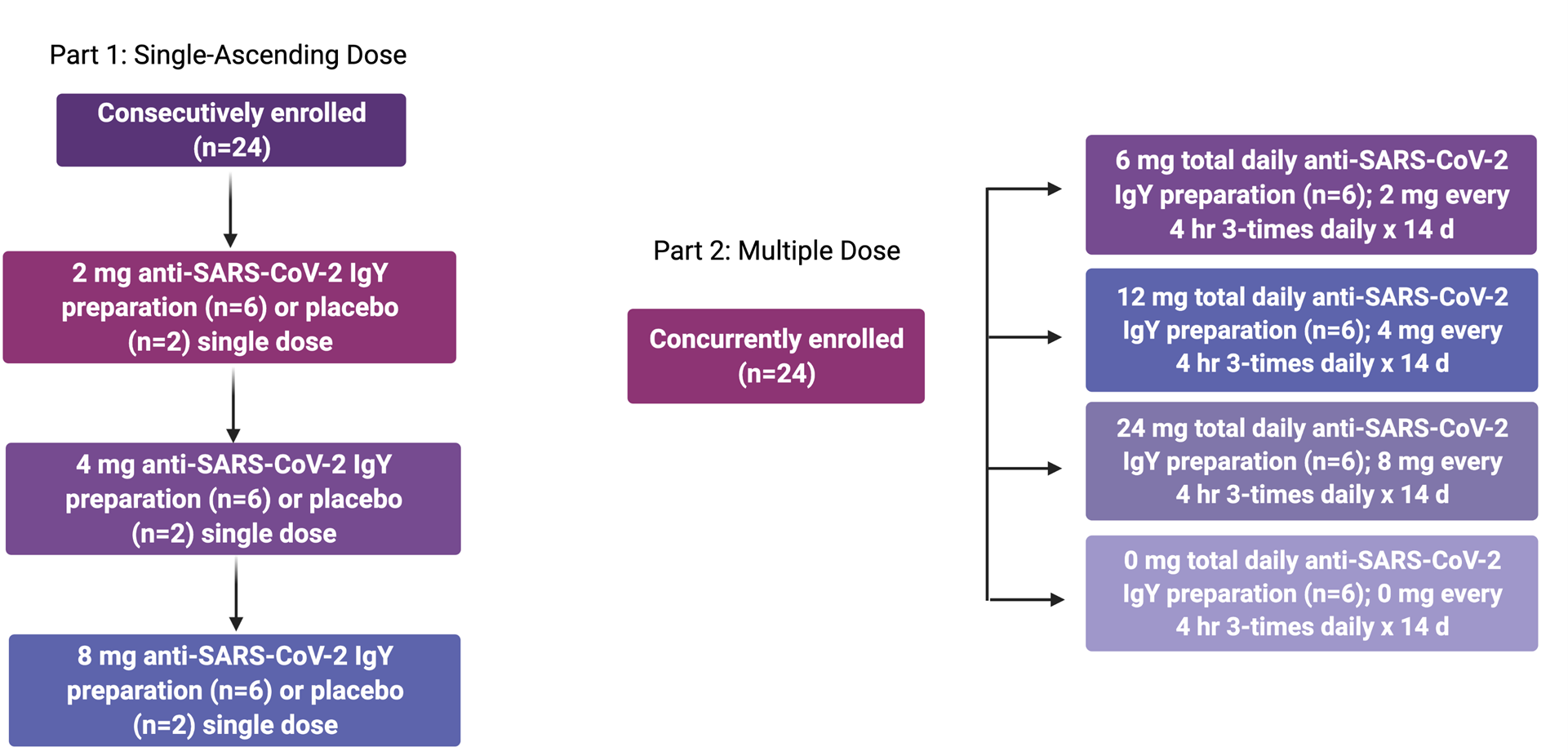
Design of phase 1 human single-ascending and multiple-dose study

Healthy male and female participants ≥18 and ≤ 45 years old with a body weight ≥ 50 kg and a body mass index ≥ 18.0 and ≤ 32.0 kg/m^2^ were eligible for this study. Females of childbearing potential who were pregnant or lactating or planning to become pregnant during the study and participants with a history of alcohol and drug abuse, current smoking, clinically significant laboratory abnormalities, history of nasal surgical procedures, frequent or recurrent nasal conditions, current use of any nasal preparations, evidence of or history of clinically significant conditions, or positive test for hepatitis B, hepatitis C, human immunodeficiency virus, or SARS-CoV-2 nucleic acid or serology were excluded from participation. Full eligibility criteria are summarized in **Supplementary Methods**.

The master randomization schedule and the associated code break envelope files were produced by an unblinded statistician using a computer-generated (SAS® v9.4 PLAN procedure) pseudo-random permutation procedure. For Part 1, the first two randomization numbers for each cohort were randomly assigned in a 1:1 ratio (anti-SARS-CoV-2 IgY: Placebo) to allow for sentinel dosing, and the remainder of the numbers for each cohort was generated in a 5:1 (anti-SARS-CoV-2 IgY: Placebo) ratio using a permuted blocked randomization with a block size of six. For Part 2, 24 numbers were generated in a 1:1:1:1 ratio (6 mg anti-SARS-CoV-2 IgY: 12 mg anti-SARS-CoV-2 IgY: 24 mg anti-SARS-CoV-2 IgY: Placebo) using a permuted blocked randomization with a block size of six. The block sizes were kept confidential during the study.

The site personnel randomized eligible participants on Day 1 by assigning the next available randomization number for the specific study part to the participant and reporting the randomization number on the case report form. Study drug was prepared by an unblinded pharmacist based on the treatment corresponding to the assigned randomization number on the randomization schedule that was only available to the pharmacist. In the event of an emergency, authorized personnel were able to unblind a participant through the code break envelope associated with the randomization number assigned to the participant.

In Part 1, participants were randomly assigned to receive a single dose of anti-SARS-CoV-2 RBD IgY antibodies or placebo in a sequential escalating manner. Three groups were sequentially dosed with 8 healthy participants per group (6 active and 2 placebo in each group). Each group in Part 1 included the initial dosing of a sentinel group (1 anti-SARS-CoV-2 RBD IgY and 1 placebo) at least 24 hours before dosing the remaining 6 participants in the cohort (5 anti-SARS-CoV-2 RBD IgY and 1 placebo). The remainder of the cohort were dosed if, in the opinion of the investigator, there were no significant safety concerns identified in the sentinel participants within the first 24 hours after administration of the dose (anti-SARS-CoV-2 RBD IgY or placebo). A Safety Monitoring Committee (SMC) reviewed safety data before each dose escalation. The following regimens were administered: 2 mg anti-SARS-CoV-2 RBD IgY preparation or placebo, 4 mg anti-SARS-CoV-2 RBD IgY preparation or placebo, and 8 mg anti-SARS-CoV-2 RBD IgY preparation or placebo. In Part 1, 2 drops were applied to each nostril as a single administration. A 7-day nontreatment follow-up period assessed safety after completion of the dosing.

In Part 2, participants were randomly assigned to receive multiple daily administrations of anti-SARS-CoV-2 RBD IgY or placebo every 4 hours (3-times daily) for 14 days in a parallel-group manner. Up to 24 healthy participants were randomized to 1 of 4 treatment regimens (6 participants per regimen). The following regimens were administered: 6 mg total daily dose anti-SARS-CoV-2 RBD IgY preparation for 14 days, 12 mg total daily dose anti-SARS-CoV-2 RBD IgY preparation for 14 days, 24 mg total daily dose anti-SARS-CoV-2 RBD IgY preparation for 14 days, and 0 mg total daily dose placebo preparation for 14 days. In Part 2, 2 drops were applied to each nostril every 4 hours (3-times daily). A 7-day nontreatment follow-up period assessed safety after completion of the dosing.

In each part, 3 groups with 8 healthy participants per group (6 active and 2 placebo in each group) were dosed. Safety and tolerability were evaluated using adverse event, physical examination (including vital signs), electrocardiogram, and clinical laboratory data. PK of anti-SARS-CoV-2 RBD IgY was evaluated by measuring serum concentrations pretreatment and at Day 14 when given as multiple doses administered intranasally for 14 days.

The investigational drug was supplied as a liquid preparation in a nose drop bottle containing 1.5 mL anti-SARS-CoV-2 RBD IgY preparation nasal suspension at 5, 10, or 20 mg/mL, or placebo, for intranasal application. Each bottle of nasal drops had enough material for one day of use. The liquid preparation contained anti-SARS-CoV-2 RBD IgY 0.5 mg/100 µL/drop, 1 mg/100 µL/drop, or 2 mg/100 µL/drop. The total maximum daily dose of anti-SARS-CoV-2 RBD IgY used in the present study (24 mg) was based in part on solubility considerations and is less than the daily dose of anti- *Pseudomonas aeruginosa* IgY previously given prophylactically as an oral (mouth rinse) treatment to prevent pulmonary infections in 17 patients with cystic fibrosis (28). For the maximum anti-SARS-CoV-2 RBD IgY dose of 4 mg/nare, we calculated a favorable ratio of IgY to viral particles, even when virus covers the nasal pathway.

All participants were provided with a Dose Administration Guide and instructed to “Gently blow your nose before using this drug. Then tilt your head back while sitting or lying down. After the study drug is administered, keep your head tilted for a few minutes. Try not to blow your nose for at least 5 minutes after study drug administration.”

#### 2.10.2 Assessments

Safety (and tolerability) were evaluated using adverse event, physical examination (including vital signs), electrocardiogram, and clinical laboratory data that included nonfasted collection of a) hematology (hemoglobin, hematocrit, erythrocyte count, mean cell volume, neutrophils, lymphocytes, monocytes, eosinophils, basophils, platelets, reticulocyte count), b) serum metabolic panel (sodium, potassium, chloride, total bilirubin, alkaline phosphatase, alanine transaminase, aspartate transaminase, blood urea nitrogen, creatinine, uric acid, phosphorous, calcium, plasma glucose, total protein, albumin, cholesterol, creatinine kinase, c) coagulation, d) urinalysis, and e) urine human chorionic gonadotropin values. Pharmacokinetics following intranasal administration of anti-SARS-CoV-2 RBD IgY were evaluated in the multiple-dose part of the study by measuring serum anti-SARS-CoV-2 RBD IgY concentration (lower limit of quantification, 30 ng/mL) at baseline and 2 hours after final dosing on Day 14, as described above (Charles River Laboratories, Reno, NV).

Serum cytokine levels for exploratory analyses were evaluated from the sera of 19 multiple-dose participants before and 2 hours after dosing on Days 1 and 2, as described above (Stanford Human Immune Monitoring Center, Stanford CA). An exploratory analysis was conducted of pretreatment serum total IgE and anti-IgE antibody (mainly anti-ovalbumin) levels in the 24 participants in the multiple-dose part of the trial.

#### 2.10.3 Changes in the Conduct of the Study

All participants were enrolled, treated, and assessed under Protocol CVR001 version 3.0, dated 29 September 2020 (**Supplementary Methods)**. The following changes were made to the conduct of the study from what was specified in the protocol:

- Participants were reconsented to allow for exploratory cytokine analyses on stored blood samples.
- Per the study protocol, serum anti-SARS-CoV-2 IgY samples were obtained from participants in the multiple-dose part of the study before dosing and at 0.5, 1, 1.5, and 2 hours after dosing on Days 1 and 14, as well as before dosing and 2 hours after dosing on Days 2, 3, and 4. Because anti-SARS-CoV-2 IgY was not measurable at the time of the theoretical maximum serum concentration at 2 hours postdose on Day 14, the remaining postbaseline PK samples were not analyzed.

#### 2.10.4 Interim Analyses

Before dose escalation in the single-ascending dose cohorts, the SMC was to review all available safety and tolerability data for a minimum of 7 participants who completed the planned safety assessments up to 48 hours after dosing. The SMC comprised three physician members (the principal investigator, sponsor medical representative, and independent medical monitor). The data was to be reviewed blinded, unless the SMC considered it necessary to unblind the data for safety concerns.

Before breaking the code per standard procedures, the potential decisions and actions were to be determined. SMC decisions on dose escalation were to be taken in consensus between the members of the SMC. The SMC decisions and their rationale were documented.

#### 2.10.5 Analyses

No formal sample size calculations were done. Based on experience for previous similar studies, the target number of participants to be enrolled was appropriate for the assessment of safety, tolerability, and PK. The planned sample size was 48 participants. A total of 48 participants were enrolled and included in the safety analyses. The analysis of safety variables included all participants who received study drug. All variables were summarized by descriptive statistics for each treatment group. The statistics for continuous variables included mean, median, standard deviation, and number of observations. Categorical variables were tabulated using frequencies and percentages. The incidence of all reported treatment-emergent adverse events and treatment-related adverse events was tabulated by treatment group. Adverse events were also classified by system organ class and preferred term using the Medical Dictionary for Regulatory Activities. Adverse events were to be listed and summarized by treatment group, preferred term, severity, seriousness, and relationship to study drug. In the event of multiple occurrences of the same adverse event with the same preferred term in one participant, the adverse event was counted once as the worst occurrence. Summary statistics for actual values and change from baseline were analyzed for laboratory results by treatment group and scheduled visit. Data summarized by treatment included adverse events, vital signs, electrocardiogram parameters, and clinical laboratory evaluations.

#### 2.10.6 Cytokine measurement and analysis

This non-GLP blinded assay was performed on sera of study participants by the Immunoassay Team at the Human Immune Monitoring Center at Stanford University (Stanford, CA), as described for the rat cytokine assay above. The levels of 80 different cytokines were determined.

#### 2.10.7 GLP bioanalytical analysis of IgY in blood samples – toxicokinetic analysis

The presence of anti-SARS-CoV-2 IgY in sera of study participants was evaluated using GLP standards in a qualified ELISA assay at Charles River Laboratories (Reno, NV). ELISA 96-well plates were coated with goat anti-chicken IgY (Thermo Fisher; A16056) and, after blocking, were incubated with the samples containing anti-SARS-CoV-2 IgY at various concentrations for 1 hour at room temperature. After washing the microplate, rabbit anti-chicken IgY (Thermo Fisher; A16130) - HRP conjugate was added and incubated for 1 hour at room temperature. The ELISA plate was washed and substrate (3,3’,5,5’-tetramethylbenzidine) was added to the wells and incubated for 20 minutes. The color development was stopped by the addition of 2N sulfuric acid and color intensity measured in a microplate reader at 450 nm. A calibration curve from the absorbance values was obtained from the standards using a 4-parameter curve fit with a weighting equation of 1/y2. The concentrations of anti-SARS-CoV-2 IgY in the samples were determined from the calibration curve. After 4 analytical method validations, lower and upper limit of detection, intra- and inter-assay precision and accuracy, dilution integrity, and short-term stability, IgY levels in study participants before and after 14 days of treatment were below the limit of detection.

### 2.11 Good Laboratory Practice

The study was performed at Charles River Laboratories following the U.S. Department of Health and Human Services, Food and Drug Administration (FDA), United States Code of Federal Regulations, Title 21, Part 58: Good Laboratory Practice for Nonclinical Laboratory Studies and as accepted by Regulatory Authorities throughout the European Union (OECD Principles of Good Laboratory Practice), Japan (MHLW), and other countries that are signatories to the OECD Mutual Acceptance of Data Agreement.

### 2.12 Animal welfare assurance and standards

The protocols and any amendment(s) or procedures involving the care and use of animals (hen immunization and rat tolerability studies) were reviewed and approved by Charles River Laboratories Institutional Animal Care and Use Committee before conduct. The hamster efficacy study was conducted following the recommendations in the Guide for the Care and Use of Laboratory Animals of the National Institutes of Health and the protocol was approved by the Institutional Animal Care and Use Committee at the Washington University School of Medicine (assurance number A3381– 01). Stanford Institutional Animal Care and Use Committee did not initially review the animal studies because they were not conducted at Stanford, but were retroactively approved.

### 2.13 Regulatory and ethics considerations

Ethical review of the clinical trial protocol and any amendments was obtained by Bellberry Human Research Ethics Committee (Australian equivalent to U.S. Institutional Review Board) and the clinical trial was conducted solely at a single investigative site, Linear Clinical Research-Harry Perkins Research Institute (Nedlands, Australia). Stanford Institutional Review Board did not review the research. The study was conducted following the protocol and ethical principles stated in the 2013 version of the Declaration of Helsinki and the applicable guidelines on Good Clinical Practice, and all applicable federal, state, and local laws, rules, and regulations.

## 3. RESULTS

### 3.1 Antigen production

The overall scheme describing the production of anti-SARS CoV-2 IgY antibody to be used as intranasal prophylaxis in humans is shown in **Figure 2A**. We produced a recombinant protein to immunize hens. A tagless RBD of SARS-CoV-2 (**Figure 2B**; index SARS-CoV-2 variant), amino acids 328-533, was produced in a cell-free protein synthesis reaction using *E.coli* extract (19,20,26). Analysis of the purified CoV-2 RBD protein yielded a single protein band with an apparent molecular weight of 23 kDa (**Figure 2C**). The purified SARS-CoV-2 RBD was eluted as a single peak by analytical size-exclusion chromatography with >95% monomer content (**Figure 2C**). Bacterial endotoxin contamination was determined to be <0.1 EU/mg by Charles River Endosafe LAL cartridge system.

**Fig. 2.**
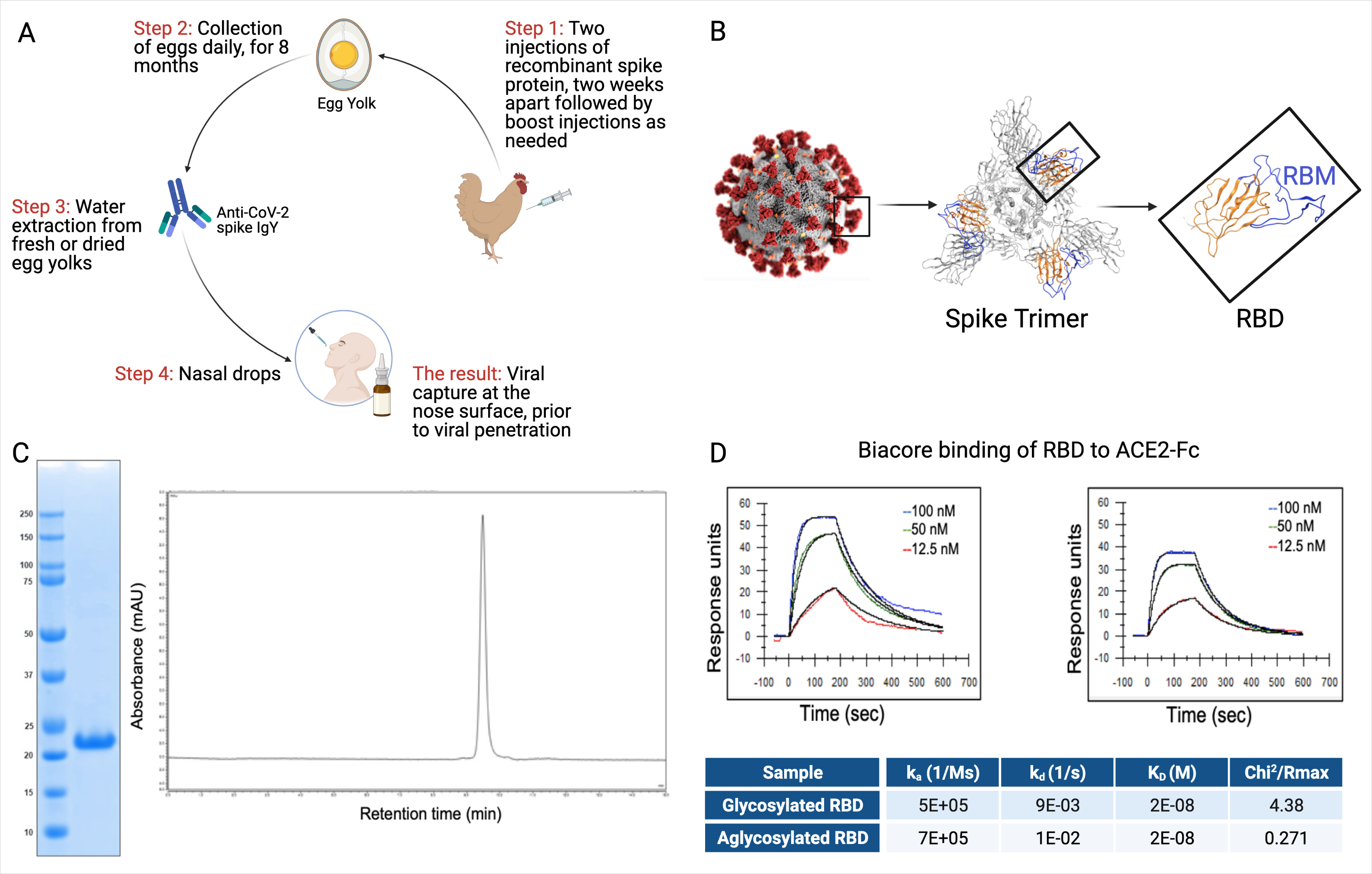
RBD and IgY preparation. **(A)** Workflow of the study. IgY preparation for intranasal drops as antiviral prophylaxis. **(B)** Cell-free expressed RBD derived from the Spike protein on the viral envelope of SARS-CoV-2. **(C)** Characterization of the recombinant protein RBD by ELISA and HPLC. **(D)** Determination of the affinity of the cell-free expressed RBD (amino acids 328-533) and mammalian-expressed full-length S1 to the hACE2 using Biacore.

Integrity of the cell-free (non-glycosylated) SARS-CoV-2 RBD was then verified by kinetic binding to the hACE2 receptor. Binding kinetics and affinity were similar to a mammalian expressed and glycosylated S1 fragment (**Figure 2D**) and were consistent with previously described binding affinities, suggesting the RBD expressed cell-free was properly folded and bioactive.

### 3.2 Hen immunization with SARS-CoV-2 RBD and IgY characterization in vitro

Cell-free expressed RBD (**Figure 2B**; 50 µg in simple oil emulsion) was injected into 9 SPF hens (46-weeks old) and IgY was extracted from egg yolks using a water-based method 2 weeks after the second immunization and thereafter. The IgY preparation was subjected to protein and Western blot analyses (**Figure 3A**). The IgY preparations were >95% pure; a quantitative Western blot analysis demonstrated that this preparation contained less than 2% ovalbumin by weight (**Figure 3A**). Chromatography of the IgY preparations on size-exclusion HPLC identified 5 peaks (**Figure 3B**); SDS-PAGE and Western blot analysis of the peaks collected between 17 and 30 minutes confirmed that these peaks all contain IgY. The anti-SARS-CoV-2 RBD IgY antibodies recognized both the immunogen, cell-free expressed RBD, and the mammalian-expressed full-length and glycosylated S1 protein (**Figure 3C**). One egg yolk of the SPF hens provided about 500 mg of purified IgY and each of >10 independent batches of IgY, purified from 100 eggs, each yielded an average of 47 ± 13 g (SD) of purified IgY (see **Figure 3D** for select examples). There was limited variability in the affinity of the various IgY batches for the S glycosylated protein as judged by ELISA (**Figure 3D**; average titer against full-length S1 was 1:18,000). Furthermore, there was almost no difference in titer of individual hens towards the full-length glycosylated S protein, suggesting minimal variability between hens (**Figures 3E, F**). Over 11 months, IgY was collected in batches of 100 eggs per preparation with a similar yield of IgY per preparation and a similar response; interruption of immunization for 3 months did not result in a drop in titer (**Figure 3F**, right panel).

**Fig. 3.**
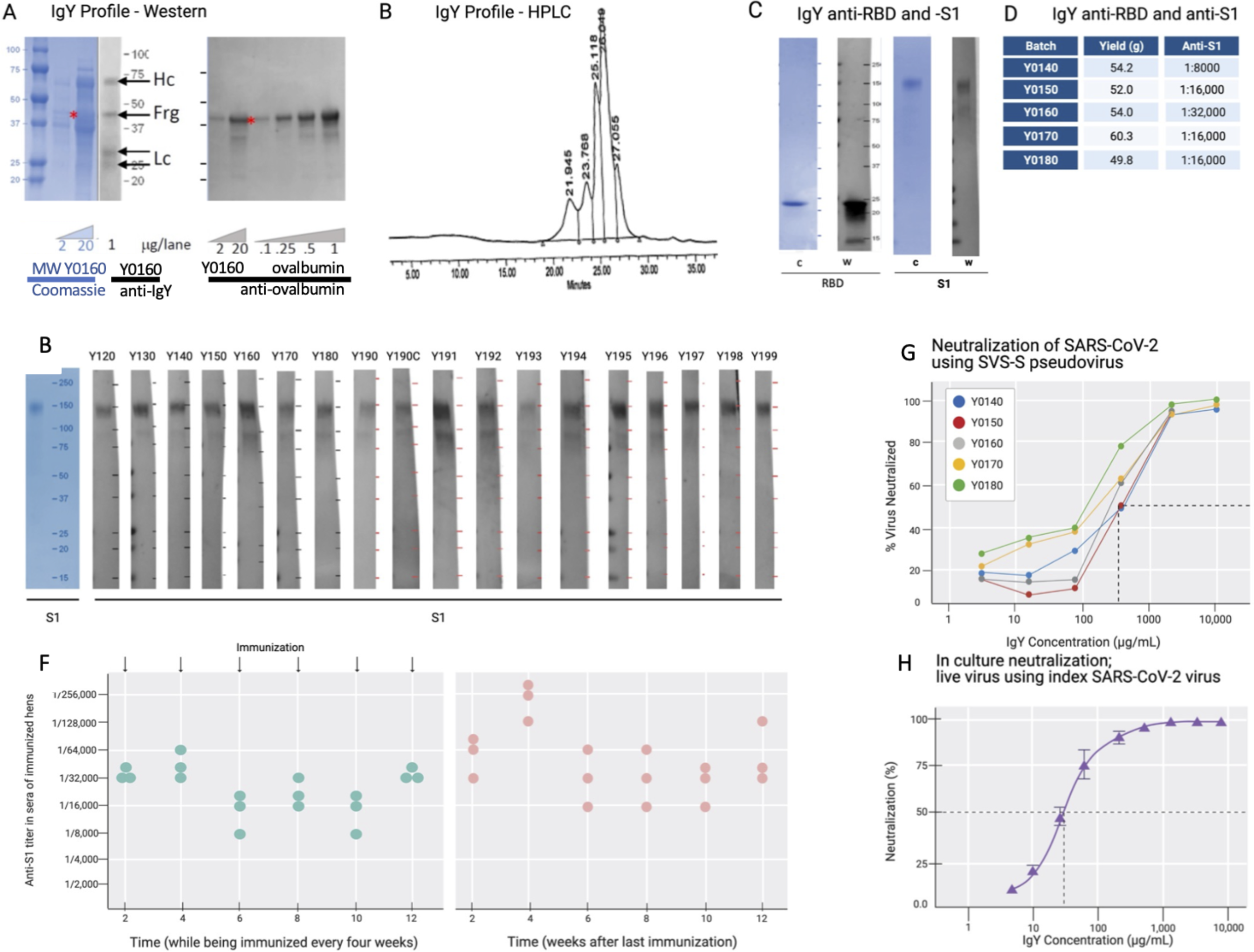
IgY purification and characterization. **(A)** Western blot analysis of the IgY preparation. **(B)** HPLC profile of the IgY preparation. **(C)** Western blot analysis of anti-SARS-CoV-2 IgY against RBD fragment and full S1 recombinant protein. **(D)** IgY yield for various batches derived from 100 eggs each. **(E)** Western blot data of different lots of anti-SARS-CoV-2 RBD IgY (Y0120-Y0199). Pools of 100 eggs laid by 9 hens over 2 weeks were used for each pool of IgY preparation between May 2020 and March 2021. IgY lot samples were diluted 1:500 followed by a 1:3000 dilution of rabbit anti-IgY HRP conjugate. First left lane shows the Coomassie stain of the same gels. **(F)** Time-dependent ELISA titers of sera from 3 individual hens following continual immunization (left); arrows indicate immunization timing. Time-dependent ELISA titer of 3 hens after immunization was stopped for up to 12 weeks (right). **(G)** Neutralization of pseudovirus SARS-CoV-2 by various lots of anti-SARS-CoV-2 RBD IgY (conducted at RetroVirox). **(H)** Neutralization of live index SARS-CoV- 2 virus by anti-SARS-CoV-2 RBD IgY (Y0180, conducted at USAMRIID).

### 3.3 Neutralization of SARS-CoV-2 index strain and variants of interest and concern with anti-SARS-CoV-2 RBD IgY

Pseudoviruses are synthetic chimeras that consist of a surrogate viral core derived from a parent virus and an envelope glycoprotein derived from a heterologous virus (29). Viral neutralization assays in culture (RetroVirox) used non-replicative VSV pseudoviruses carrying a firefly luciferase reporter gene and expressing S of SARS-CoV-2 on the surface of the virion (VSV-S). The neutralization assay was performed with HEK 293T-hACE2, a human embryonic kidney cell line overexpressing hACE2, the receptor of the SARS-CoV-2 virus. First, 5 batches of RBD IgY preparations were tested. The neutralization activity of the purified IgY, defined as the concentration inhibiting 50% of the viruses (IC50) was ∼170 µg/mL (**Figure 3G**). Importantly, ∼10-fold higher neutralization activity towards the index SARS-CoV-2 virus was observed when using a live index virus (**Figure 3H**).

Approximately 30 µg/mL IgY provided 50% neutralization of the index virus in culture (**Figure 3H**).

Because at least 13 common variants of SARS-CoV-2 with amino acid mutations in the RBD had emerged since December 2020 (30–32) (**Figure 4A**), we tested the activity of the anti-SARS-CoV-2 IgY against several variants (including Beta, Delta, and Omicron; **Figures 4A, B**) and D614G, an amino acid substitution outside the RBD that is now found in most variants. Beta, Delta, and Omicron were classified as variants of concern, associated with increased transmissibility or detrimental change in COVID-19 epidemiology, or an increase in virulence or change in clinical disease presentation.

**Fig. 4.**
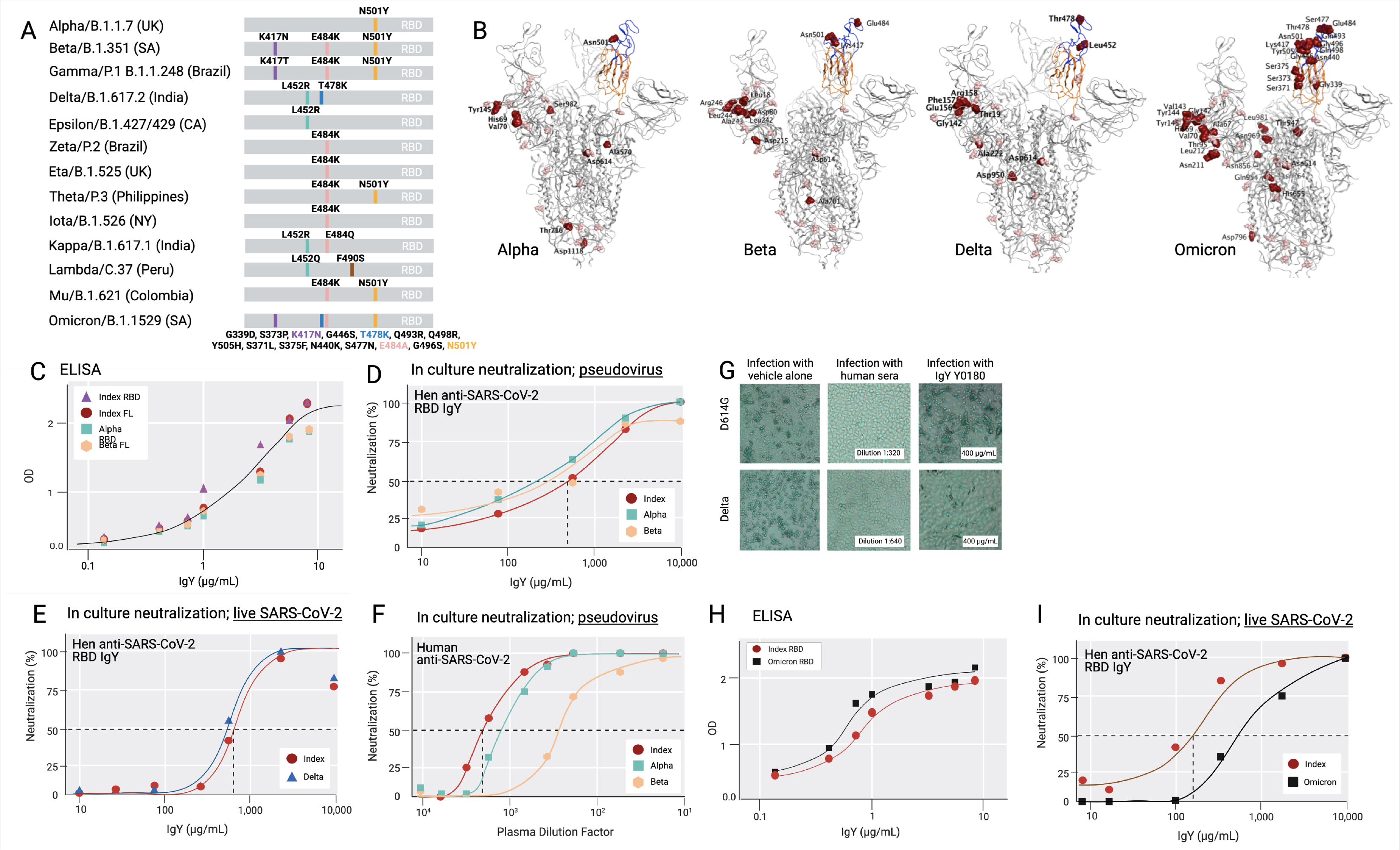
Common SARS-CoV-2 variants and anti-SARS-CoV-2 IgY interaction with them. **(A)** A scheme depicting locations of mutated amino acids in Alpha through Mu variants of SARS-CoV-2, focusing on the RBD domain only. Each color bar indicates the amino acid in the index virus that was mutated in the variant. **(B)** Spike protein of SARS-CoV-2 Alpha, Beta, Delta, and Omicron are shown from left to right. Molecular Operating Environment was used to create the figure (34). The location of mutations in the structure of the S protein trimer of SARS-CoV-2 (PDB ID: 7A98) for 4 of the common variants are indicated in red and glycosylation sites are indicated in pink throughout the S protein. Blue ribbon indicates RBD (amino acids 328-533) and the orange ribbon indicates receptor binding motif (amino acids 437-508). **(C)** Binding of anti-SARS-CoV-2 RBD IgY to recombinant S1 full length (FL) of the index virus, the RBD of the Alpha and Beta variants, and the immunizing RBD of the index virus by ELISA. **(D)** Neutralization of pseudovirus (VSV-S) SARS-CoV-2 carrying S protein of index virus, Alpha, or Beta variants by anti-RBD IgY. **(E)** Neutralization of live index or Delta viruses by anti-SARS-CoV-2 IgY against the RBD. **(F)** Neutralization of pseudoviruses listed in (D) by human serum of immunized individuals. **(G)** Neutralization of live D614G vs. Delta variants by human serum of immunized individual or by anti-SARS CoV-2 IgY. Microscopic evaluation of monolayers of Vero E6 cells after 96 hours infection with the indicated authentic (live) SARS-CoV-2 variant. Images from infected cells are shown after 4 days of infection with SARS-CoV-2 variants in the absence or presence of test items. Top three panels: Infection in the presence of MEX-BC2/2020 and bottom three panels: infection with the Delta variant each in the presence of vehicle alone, serum of a person immunized twice with the Moderna (mRNA-1273) vaccine or anti-SARS CoV-2 RBD IgY, as indicated, all at the indicated concentration (neutralization experiments in panels D-H & I were conducted by RetroVirox using pseudovirus or live virus, as indicated). **(H)** Binding of anti-SARS-CoV-2 RBD IgY to the index virus and Omicron variant (B.1.1.529) RBD domain using ELISA. **(I)** Neutralization of live index or Omicron variant of SARS-CoV-2 by anti-SARS-CoV-2 RBD IgY. Except when indicated, the studies were done over several months; therefore, the absolute titers in the ELISA and neutralization studies were not identical. However, each experiment included the same positive control; index RBD for ELISA and index virus for neutralization assays.

First, IgY antibody ELISA titer against Beta RBD was compared with the RBD of the index SARS-CoV-2 virus as well as the most common Alpha variant. ELISA with recombinant full-length S protein or the RBD of the 3 mutants as well as the immunizing RBD fragment of the index virus yielded a virtually identical titer (**Figure 4C**). Although the Omicron variant still uses the hACE2 receptor to infect human cells (10, 33), the RBD contains a total of 15 mutations compared to the index virus, 11 of which were not found in the previous variants (**Figure 4A, B**). Yet, the ELISA titer of IgY against the Omicron RBD was also equivalent or slightly better than that towards the RBD of the index virus (**Figure 4H**).

Next, we tested the neutralization titer of anti-SARS-CoV-2 RBD IgY (lot Y0180) against the RBD of the index, Alpha, and Beta variants, thus including all the amino acid substitutions within the RBD also found in the RBD of Gamma, Zeta, Eta, Theta, Iota, and Mu variants and 1 of the 2 substitutions in the Kappa variant (**Figure 4A**). The IC50 of the VSV-S pseudovirions for the index strain and Beta variant were virtually identical: 668 μg/mL for the index strain, 568 μg/mL for Beta (**Figure 4D**), and 2-fold lower for Alpha (IC50 = 302 μg/mL; **Figure 4D**). RetroVirox also provided data using plasma from a single Moderna-vaccinated individual for a titer comparison of neutralization with anti-SARS-CoV-2 RBD IgY, tested with 3 of the variants. In a side-by-side study, the IC50 generated with the human plasma from a recipient of the mRNA Moderna vaccine (2 doses; index SARS-CoV-2) showed the highest titer against the index virus, followed by a 2.8-fold drop in titer towards Alpha and a 6.7-fold lower titer for Beta (**Figure 4F**).

The anti-SARS-CoV-2 IgY preparation was similarly effective against the Delta variant compared with the index strain (**Figure 4E**). The assays show that Y0180 displays similar neutralizing activity against both isolates, with slightly reduced neutralizing activity against Omicron (**Figures 4E, I**); IC50 values generated were 635 μg/mL (Delta) and 739 μg/mL (index virus). The neutralization assay against Omicron generated IC50 values of 143 μg/mL (index) and 785 μg/mL (Omicron) (**Figure 4I**). Plasma from a Moderna-vaccinated individual was also tested in parallel with both isolates. NT50 values generated with this plasma against the variants were 1:1274 (Delta pseudovirus) and 1:1091 (index pseudovirus). The neutralization activities of the antibody and control plasma against both variants were also confirmed by microscopy evaluating the virus-induced CPE in infected cell monolayers **Figure 4G**). The human serum had a similar degree of neutralization against the index pseudovirus and Delta (**Figure 4G**) when tested at a dose that is three times higher than that required for 50% neutralization (1:640 in **Figure 4H**, middle panels, vs. 1:2,000 in **Figure 4F**). In contrast, anti-SARS-CoV-2 IgY tested at 400 µg/mL (**Figure 4G**, right panels), a dose below that required for 50% neutralization (∼650 µg/mL; **Figure 4E**), was equally effective against both variants. Together, these data indicate that the spectrum of the polyclonal anti-SARS-CoV-2 RBD IgY displays sufficient diversity so that none of the common point mutations in the RBD associated with a greater transmission rate of the virus affected the neutralization efficacy of these most common SARS-CoV-2 variants of concern.

### 3.4 Efficacy study in a hamster model of COVID-19

We did not find in vivo efficacy of anti-SARS-CoV-2 RBD IgY in the Syrian golden hamster COVID-19 model against a challenge with a titer of 0.8 or 4 x 10^6^ of SARS-CoV-2 virus, likely because the amount of virus we employed was too high. Lung viral load after 3 days was comparable between the control treatment group and the anti-SARS-CoV-2 RBD IgY treatment group, but body weight loss was more severe in control animals (**Supplementary Figure S2**).

### 3.5 Rat toxicity and safety study

All 8-week-old Sprague Dawley rats (10 males and 10 females) in the 28-day GLP safety study survived to scheduled euthanasia with no mortality, test article-related organ weight changes, or gross or microscopic findings (see **Figure 5B**). There were also no differences between female and male groups in each treatment arm. The GLP-qualified assay detected no anti-SARS-CoV-2 RBD IgY in the sera of animals after 28 days of daily treatment with 4 mg anti-SARS-CoV-2 RBD IgY (lower limit of detection of 30 ng/mL).

**Fig. 5.**
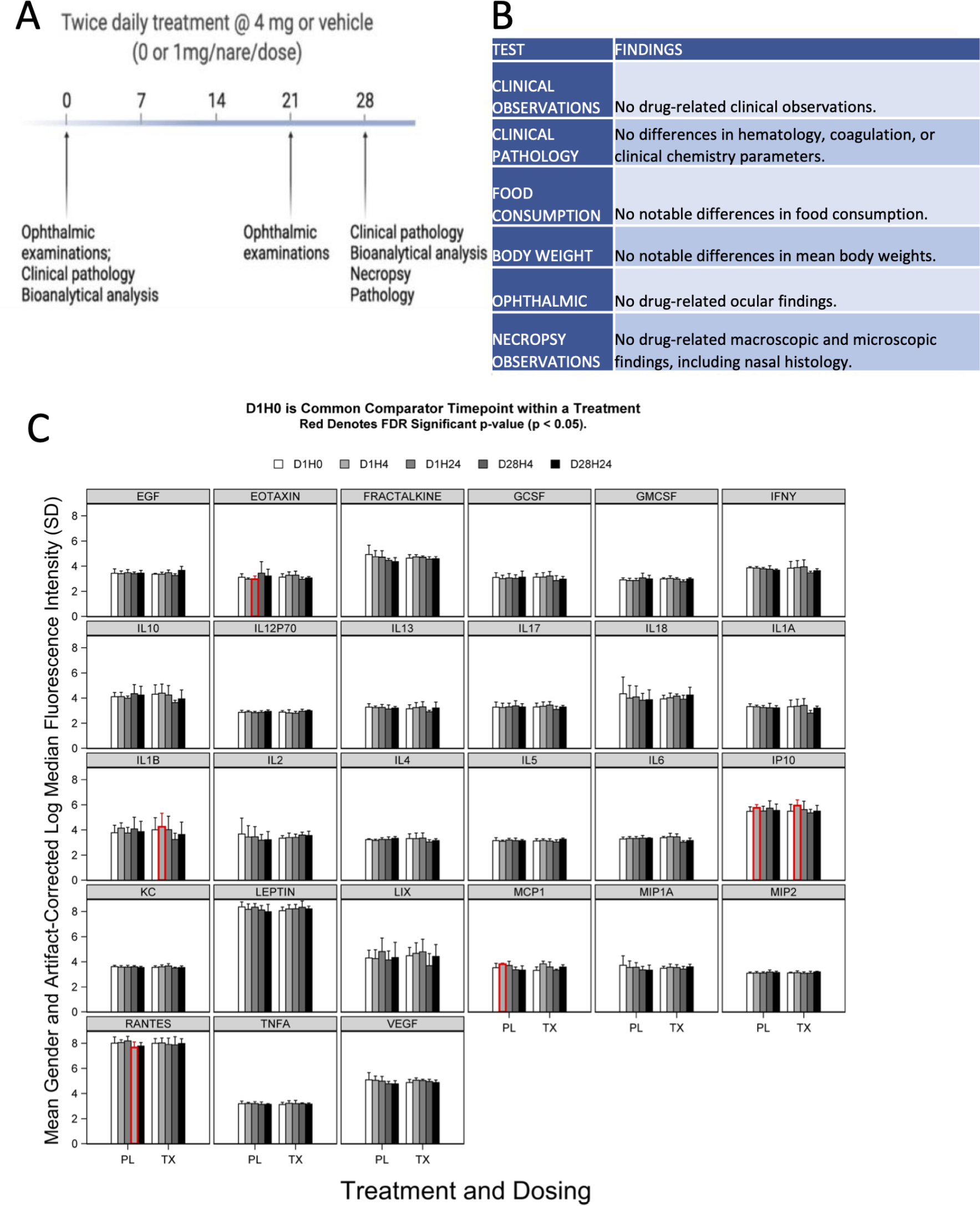
Preclinical toxicity of 28-day treatment with anti-SARS-CoV-2 RBD IgY in rats. **(A)** Study design. **(B)** Summary of findings. **(C)** Serum levels of 27 cytokines over time in rats treated with IgY (Tx) or vehicle/placebo (PL). Data are provided for Day 1 before treatment (D1H0); Day 1, 4 hours after the treatment (D1H4); 24 hours after the two treatments, 6 hours apart, at 24 hours after the first treatment (D1H24); 28 days of twice-daily treatments and 4 hours of the treatment of that day (D28H4); and 28 days of twice-daily treatments and 24 hours of the treatment of that day (D28H24). Red indicates a statistical difference with a false discovery rate (FDR) significant p-value (p < 0.05)

There was no evidence of significant systemic immune activation in the rats (20/group) treated as above by measuring changes in levels of 27 proinflammatory serum cytokines after anti-SARS-CoV-2 RBD IgY administration (**Figure 5C**). When comparing posttreatment (Day 1 at 4 hours and Day 28 at 24 hours after twice-daily treatments) to pretreatment serum levels (Day 1, time 0; D1 H0), there were no changes for most of the cytokines (22 of the 27 tested) in both placebo- and IgY-treated groups at any time. Compared with D1 H0, we detected a transient, slight increase in interleukin-1 (IL-1) beta and monocyte chemoattractant protein-1 (MCP1) on Day 1, 4 hours after anti-SARS-CoV-2 RBD IgY administration. A similar trend that did not reach significance was also observed in the placebo group at the same time; increases in IL-1 beta and MCP1 were not seen at any other time. There was a significant increase in interferon-inducible protein 10 (IP10) on Day 1, 4 hours after the first treatment compared to D1 H0, observed in both treatment and placebo group. A significant decrease in D1 H24 in the placebo arm was observed only for RANTES. Nonetheless, none of these changes in cytokines were observed in the rats at other time points. Together, our data show that long-term twice-daily administration of 4 mg/mL IgY (or 16 mg/kg) up to 28 days in rats has excellent safety and tolerability with no evidence of systemic immune activation.

### 3.6 Human tissue cross-reactivity study

We determined potential cross-reactivity of the anti-SARS-CoV-2 RBD IgY protein to a full panel of human tissues (at least 3 donors per tissue; see **Supplementary Table S4** for a list of human tissues tested for reactivity). Anti-SARS-CoV-2 RBD IgY reactivity at two concentrations (20 and 10 µg/mL) was compared with control polyclonal chicken IgY antibodies (negative control), and with anti-human macroglobulin (positive control for staining). No specific binding was observed with the anti-SARS-CoV-2 RBD IgY to any of the human tissue panels examined, including human nasal cavity and lung (see **Figure 6**).

**Fig. 6.**
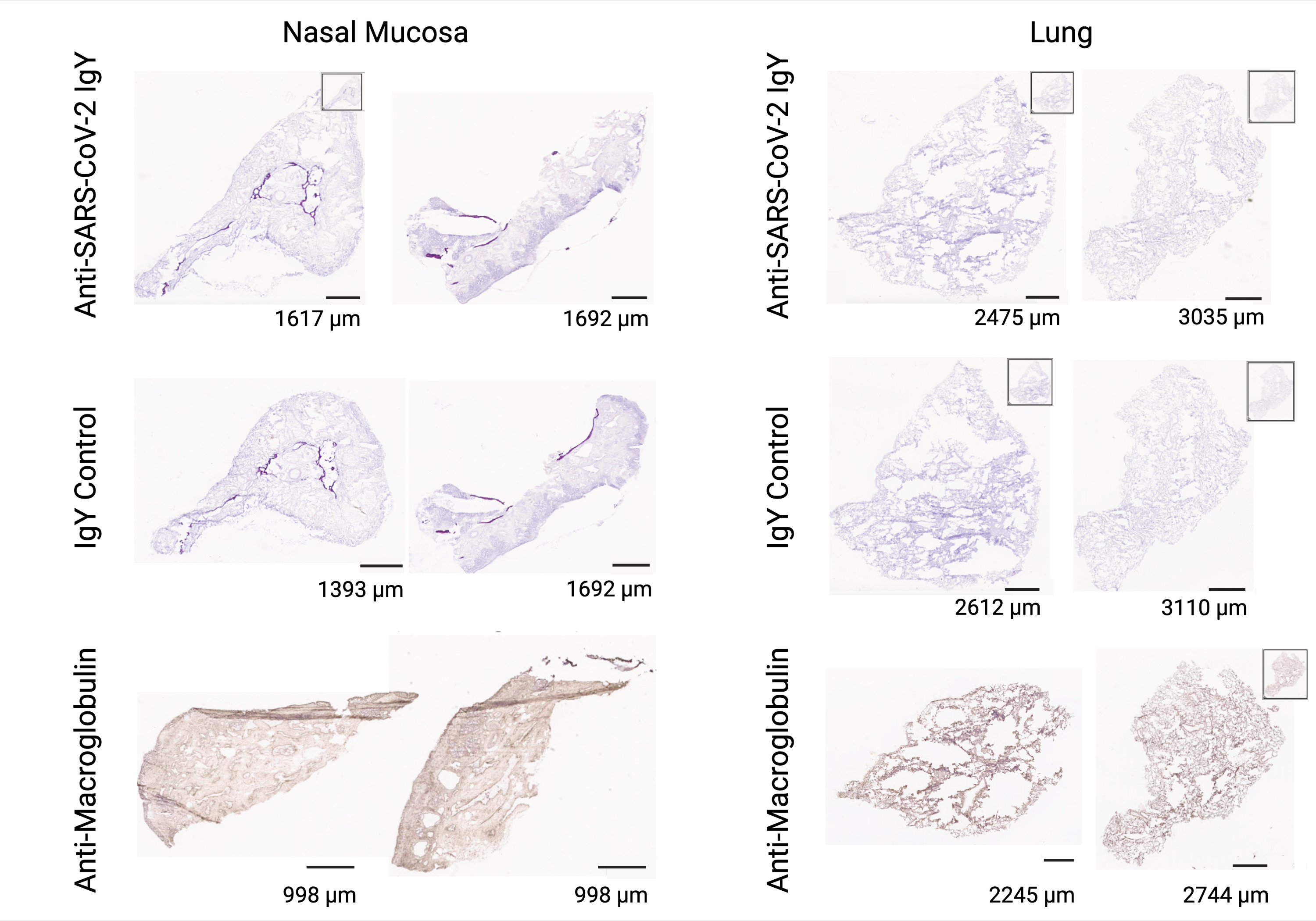
Lack of cross-reactivity of anti-SARS-CoV-2 RBD IgY with human tissues. Immunohistochemical testing of anti-SARS-CoV-2 RBD IgY (top row), control IgY (middle row; negative control), and anti-human macroglobulin antibodies (bottom row; positive control) with human nasal mucosa (left two panels) and human lungs (right two panels). Bars provide a magnification scale.

### 3.7 Phase 1 Clinical Trial Results

Forty-seven of 48 enrolled participants completed the study drug treatment period and planned study visits (**Supplementary Figure S1, CONSORT flow diagram**). One participant in the multiple-dose part was withdrawn from the study after receiving 3 doses (Day 1) of placebo due to a concurrent upper respiratory tract infection, judged to be unrelated to study drug by the investigator.

#### 3.7.1 Baseline Demographics

Study participants ranged in age from 20 to 43 years (median, 25.5) in the single-dose part of the study and 18 to 40 years (median, 23.0) in the multiple-dose part (**Tables 1, 2**). Female participants comprised 75% of the population in the single-dose study segment and 46% in the multiple-dose study segment. Demographics are summarized in **Tables 1 and 2**.

**Table 1.** Demographics of participants in single-ascending dose group (Part 1).

| Anti-SARS-CoV-2 IgY |  |  |  |  | All Participants<br>N=24 |
| --- | --- | --- | --- | --- | --- |
|  | 2 mg<br>N=6 | 4 mg<br>N=6 | 8 mg<br>N=6 | Placebo<br>N=6 |  |
| **Age, years** |  |  |  |  |  |
| Mean (SD) | 28.8 (7.99) | 27.3 (4.76) | 27.7 (5.65) | 22.7 (2.34) | 26.6 (5.72) |
| Median (range) | 26.5 (20-43) | 28.0 (21-33) | 27.5 (20-37) | 22.0 (20-27) | 25.5 (20-43) |
| **Gender, n (%)** |  |  |  |  |  |
| Female | 4 (66.7) | 4 (66.7) | 4 (66.7) | 6 (100) | 18 (75.0) |
| Male | 2 (33.3) | 2 (33.3) | 2 (33.3) | 0 | 6 (25.0) |
| **Ethnicity, n (%)** |  |  |  |  |  |
| Hispanic or Latino | 1 (16.7) | 0 | 0 | 0 | 1 (4.2) |
| Not Hispanic or Latino | 5 (83.3) | 6 (100) | 6 (100) | 6 (100) | 23 (95.8) |
| **Race, n (%)** |  |  |  |  |  |
| Asian | 2 (33.3) | 1 (16.7) | 0 | 0 | 3 (12.5) |
| Native Hawaiian or Other Pacific Islander | 0 | 0 | 0 | 1 (16.7%) | 1 (4.2%) |
| White | 4 (66.7) | 5 (83.3) | 6 (100) | 5 (83.3) | 20 (83.3) |

**Table 2:** Demographics of participants in the multiple-dose group (Part 2).

| Anti-SARS-CoV-2 IgY | Anti-SARS-CoV-2 IgY |  |  | Placebo TID<br>N=6 | All Participants<br>N=24 |
| --- | --- | --- | --- | --- | --- |
|  | 2 mg TID<br>N=6 | 4 mg TID<br>N=6 | 8 mg TID<br>N=6 |  |  |
| **Age, years** |  |  |  |  |  |
| Mean (SD) | 26.0 (8.37) | 21.8 (2.14) | 27.5 (6.92) | 25.3 (6.41) | 25.2 (6.33) |
| Median (range) | 23.0 (18-40) | 22.0 (19-25) | 25.0 (21-37) | 25.0 (18-36) | 23.0 (18-40) |
| **Gender, n (%)** |  |  |  |  |  |
| Female | 1 (16.7) | 4 (66.7) | 2 (33.3) | 4 (66.7) | 11 (45.8) |
| Male | 5 (83.3) | 2 (33.3) | 4 (66.7) | 2 (33.3) | 13 (54.2) |
| **Ethnicity, n (%)** |  |  |  |  |  |
| Hispanic or Latino | 0 | 0 | 1 (16.7) | 1 (16.7) | 2 (8.3) |
| Not Hispanic or Latino | 6 (100) | 6 (100) | 5 (83.3) | 5 (83.3) | 22 (91.7) |
| **Race, n (%)** |  |  |  |  |  |
| Asian | 1 (16.7) | 2 (33.3) | 2 (33.3) | 2 (33.3) | 7 (29.2) |
| Black or African American | 0 | 0 | 0 | 1 (16.7) | 1 (4.2) |
| White | 5 (83.3) | 4 (66.7) | 4 (66.7) | 3 (50.0) | 16 (66.7) |

tid = 3-times daily

#### 3.7.2 Safety and tolerability

The overall incidence of treatment-emergent adverse events was 29% (7 of 24 participants; **Table 3**) in the single-dose part of the study and 58% (14 of 24 participants; **Table 4**) in the multiple-dose part of the study, with similar incidence rates between anti-SARS-CoV-2 RBD IgY (42%) and placebo (50%) groups. The most frequent treatment-emergent adverse event was headache, with similar rates between placebo (17%) and anti-SARS-CoV-2 RBD IgY (14%) (**Tables 3 and 4**).

**Table 3.**
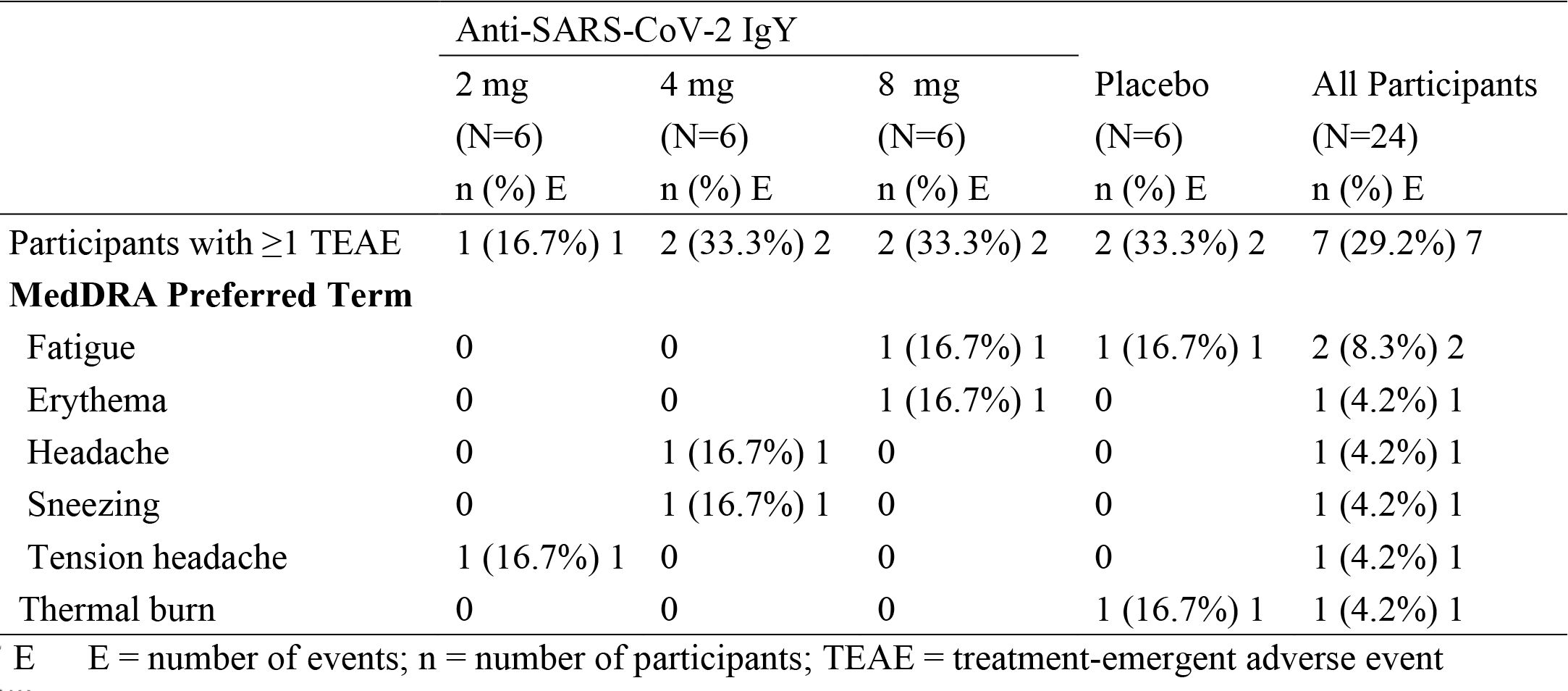
Adverse events by preferred term- single ascending-dose study (Part 1).

|  | Anti-SARS-CoV-2 IgY |  |  | Placebo<br>(N=6)<br>n (%) E | All Participants<br>(N=24)<br>n (%) E |
| --- | --- | --- | --- | --- | --- |
|  | 2 mg<br>(N=6)<br>n (%) E | 4 mg<br>(N=6)<br>n (%) E | 8 mg<br>(N=6)<br>n (%) E |  |  |
| Participants with $\geq 1$ TEAE | 1 (16.7%) 1 | 2 (33.3%) 2 | 2 (33.3%) 2 | 2 (33.3%) 2 | 7 (29.2%) 7 |
| **MedDRA Preferred Term** |  |  |  |  |  |
| Fatigue | 0 | 0 | 1 (16.7%) 1 | 1 (16.7%) 1 | 2 (8.3%) 2 |
| Erythema | 0 | 0 | 1 (16.7%) 1 | 0 | 1 (4.2%) 1 |
| Headache | 0 | 1 (16.7%) 1 | 0 | 0 | 1 (4.2%) 1 |
| Sneezing | 0 | 1 (16.7%) 1 | 0 | 0 | 1 (4.2%) 1 |
| Tension headache | 1 (16.7%) 1 | 0 | 0 | 0 | 1 (4.2%) 1 |
| Thermal burn | 0 | 0 | 0 | 1 (16.7%) 1 | 1 (4.2%) 1 |

E = number of events; n = number of participants; TEAE = treatment-emergent adverse event

**Table 4.** Adverse events by preferred term- multiple-dose study (Part 2).

|  | Anti-SARS-CoV-2 IgY |  |  | Placebo TID<br>(N=6)<br>n (%) E | All Participants<br>(N=24)<br>n (%) E |
| --- | --- | --- | --- | --- | --- |
|  | 2 mg TID<br>(N=6)<br>n (%) E | 4 mg TID<br>(N=6)<br>n (%) E | 8 mg TID<br>(N=6)<br>n (%) E |  |  |
| Participants with $\geq 1$ TEAE | 4 (66.7%) 7 | 2 (33.3%) 3 | 4 (66.7%) 5 | 4 (66.7%) 5 | 14 (58.3%) 20 |
| **MedDRA Preferred Term** |  |  |  |  |  |
| Headache | 0 | 0 | 3 (50.0%) 3 | 2 (33.3%) 2 | 5 (20.8%) 5 |
| Upper respiratory tract infect | 0 | 0 | 1 (16.7%) 1 | 1 (16.7%) 1 | 2 (8.3%) 2 |
| Contusion | 0 | 0 | 0 | 1 (16.7%) 1 | 1 (4.2%) 1 |
| Dental discomfort | 1 (16.7%) 1 | 0 | 0 | 0 | 1 (4.2%) 1 |
| Dizziness | 1 (16.7%) 1 | 0 | 0 | 0 | 1 (4.2%) 1 |
| Ear pain | 1 (16.7%) 1 | 0 | 0 | 0 | 1 (4.2%) 1 |
| Epistaxis | 1 (16.7%) 1 | 0 | 0 | 0 | 1 (4.2%) 1 |
| Eyelid irritation | 0 | 0 | 1 (16.7%) 1 | 0 | 1 (4.2%) 1 |
| Injection site haematoma | 1 (16.7%) 1 | 0 | 0 | 0 | 1 (4.2%) 1 |
| Nasal congestion | 0 | 0 | 0 | 1 (16.7%) 1 | 1 (4.2%) 1 |
| Parosmia | 0 | 1 (16.7%) 1 | 0 | 0 | 1 (4.2%) 1 |
| Presyncope | 1 (16.7%) 1 | 0 | 0 | 0 | 1 (4.2%) 1 |
| Rhinorrhoea | 0 | 1 (16.7%) 1 | 0 | 0 | 1 (4.2%) 1 |
| Skin abrasion | 0 | 1 (16.7%) 1 | 0 | 0 | 1 (4.2%) 1 |
| Tenderness | 1 (16.7%) 1 | 0 | 0 | 0 | 1 (4.2%) 1 |

E = number of events; n = number of participants; TEAE = treatment-emergent adverse event

All adverse events were mild (grade 1) in severity. No serious adverse event or lab-related adverse event was reported, and there was no dose dependency of adverse events observed.

Furthermore, no participant receiving anti-SARS-CoV-2 RBD IgY had an adverse event of nasal irritation or nasal congestion. There were no clinically significant observations or trends noted in laboratory assessments, vital signs, physical exam findings, or electrocardiograms during the study.

#### 3.7.3 Pharmacokinetics

PK analyses indicated no evidence of serum anti-SARS-CoV-2 RBD IgY above the lowest detection levels of 30 ng/mL (using a GLP study at Charles River Laboratories) in the 18 participants who received anti-SARS-CoV-2 RBD IgY in the multiple-dose part of the study.

#### 3.7.4 Serum cytokines

Levels of 80 different cytokines in sera of 19 participants of the multiple-dose part were tested before treatment (D1 pre-dose), 2 hours after dosing on day 1 (D1 H2); day 2, before treatment (D2 pre-dose), and 2 hours after the first dosing on day 2 (D2 H2; **Supplementary Figure S3**). There were slight but statistically significant decreases (red histograms) in 10 of the 80 cytokines (**Supplementary Figure S3**). These slight declines in CCL27, CXCL9, IL23, IL27, LIF, MIP5, RESISTIN, TNFα, TNFα, and TNFRSF6 were noted only in the 12 mg/day group compared to the pretreatment levels. These declines also occurred only 2 hours after the first dose (D1 H2) and were not sustained, except for MIP5. There was also a small decrease in IL3, 2 hours after the first dose of 6 mg/day group anti-SARS-CoV-2 RBD IgY, but there were no changes in this cytokine at any other times or doses. These slight changes, which were also not sustained or dose dependent, were judged to be artifactual. Overall, there were no clinically relevant increases in serum cytokines at any time for any of the treatment groups (6, 12, or 24 mg total daily dose of anti-SARS CoV-2 RBD IgY for 14 days).

#### 3.7.5 IgE and anti-IgE

The anti-SARS-CoV-2 IgY preparation contains ovalbumin. Although participants with egg allergies were excluded from the trial, an exploratory analysis was conducted of pretreatment serum total IgE and anti-IgE antibody (mainly anti-ovalbumin) levels in the 24 participants in the multiple-dose part of the trial. No participant had detectable serum egg-white specific IgE antibodies (all <0.35 kU/L).

## 4. DISCUSSION

Despite recent successes in generating highly effective COVID-19 vaccines, there is an ongoing need for widely available and safe antiviral strategies that reduce infection and transmission worldwide.

Limitations to current vaccines include global vaccine availability and affordability, vaccine hesitancy, and rapidly emerging highly infective viral strains that escape vaccine-induced immunity. This has been particularly apparent following the emergence of Delta and Omicron. The latter variant was first detected in specimens collected on November 8, 2021 (35), within a few weeks became the dominant SARS-CoV-2 variant in the United States (36), and is now the most common strain globally (13). Both convalescent sera from early strain-infected patients and fully vaccinated individuals exhibited a low neutralization capacity against Omicron (9–12); a reduction of 30 to 40- fold in neutralization titers was reported. Furthermore, of eight currently authorized or approved monoclonal antibodies, seven did not neutralize the Omicron variant and one had a 3-fold reduction in neutralization titer (11). These data highlight the need for alternative and complementary approaches to curb COVID-19.

Here, we describe the production of the first chicken egg yolk-derived anti-index SARS-CoV- 2 RBD IgY polyclonal antibodies as an intranasal drop product for humans with equal in vitro activity against all variants of concern. These IgY were raised in SPF hens and showed an excellent safety profile when given intranasally by drops to rats for 28 days (4 mg/day). No toxicity, innate inflammatory response, or systemic exposure to IgY were noted in this GLP study. In 48 healthy adult participants, anti-SARS-CoV-2 RBD IgY given intranasally at single-ascending doses of 2, 4, and 8 mg and as total daily doses of 6, 12, and 24 mg for 14 days also had a highly favorable safety and tolerability profile. Importantly, no participant receiving intranasal anti-SARS-CoV-2 IgY in the multiple-dose phase had measurable levels of anti-SARS-CoV-2 RBD IgY in their sera, reflecting the absence of systemic absorption of topically administered IgY following intranasal application. We also found no evidence of a systemic inflammatory immune response triggered by the topical treatment with anti-SARS-CoV-2 RBD IgY in humans, and no detectable increase in 80 sera cytokines.

Hen-derived IgY antibodies have several advantages for topical use in humans; these antibodies do not bind the Fc receptor or rheumatoid factor or activate the human complement cascade (37), thus greatly reducing the risk of severe immune responses. These features support the clinical applications of IgY for nasal treatment in a wide range of persons, including the elderly, immunocompromised, and children. IgY have been beneficial with favorable safety and tolerability when given prophylactically in both animal models and clinical settings of viral diseases, including respiratory infections [reviewed in (38)]. Overall, available data to date suggest that IgY given by nonparenteral administration do not have unwanted off-target proinflammatory effects and are nontoxic to humans, thus permitting potential clinical applications in diverse populations and diseases.

The potential use of IgY from hens immunized with inactivated virus (39), full-length recombinant S protein (40–43), or N protein (44) has been explored with studies evaluating neutralization of the virus in cells, with IC50 values of 10 mg/mL (39), 1 mg/mL (40), 16.8 mg/mL (41), and 0.27 mg/mL (42). However, the ability of these egg-derived antibodies to neutralize other common SARS-CoV-2 variants has not been evaluated and their safety profile in animals or humans has not been assessed.

Intranasally administered proteins are removed from the mucosal surface through ciliary movement (40, 45), which was the basis for using a 3-times daily (every 4 hours) regimen in our phase 1 study. The anti-SARS-CoV-2 RBD IgY was designed to capture and immobilize SARS-CoV-2 on the nasal mucosa, preventing the virus from binding to and spreading across the nasal mucosa, and also preventing the transmission of the virus to other individuals. Intranasal delivery of mammalian immunoglobulins as antiviral agents has been extensively evaluated in humans (46–51). Human immunoglobulins G (IgG) and A given intranasally are well tolerated (46–51), including in pediatric populations. Our decision to protect from viral entry at the nasal mucosa stems from the observation that levels of lung hACE2 are much lower than in the nose; infection of lung tissue is >5 orders of magnitude lower compared with nasal mucosa (52, 53). Therefore, inhibition of viral entry at the nose is likely the correct target site for optimal efficacy.

Our product is egg-derived immunoglobulins, which could contain potentially antigenic residual chicken proteins. However, it is not indicated for those who are allergic to egg yolks. Note also that most humans are exposed to egg-derived antigens through their diet and are not allergic. Furthermore, anaphylaxis for those who consume eggs regularly is rare. However, the safety and tolerability of hen-derived IgY as intranasal treatment in humans have not been described despite their extensive use in a variety of routes in animals and aquaculture (38, 54).

Several other studies have examined anti-COVID-19 intranasal prophylaxis, mostly in animal models (55–61). These prophylaxes include polymer barriers, active vaccines, existing antiviral drugs, inhibitors of protease-induced activation of the virus, antiseptics, antimicrobial agents, and antibodies. Most relevant for comparison with our study is the use of neutralizing antibodies. In one study, intranasal treatment with a monoclonal human antibody (500 µg in 100 µL/nare) 12 hours after infection in hamsters inoculated with 5 x 10^4^ median tissue culture infective dose (TCID50) of SARS-CoV-2 decreased clinical disease signs and improved recovery during 9 days of infection compared with control antibody-treated hamsters (61). However, in contrast to our study, there was a substantial systemic exposure to the human antibody 24 hours after a single intranasal treatment with 2.5 mg; serum levels of the treated antibodies in that study were 210 ng/mL vs. below detection levels (30 ng/mL) in our study following administration of 4 mg IgY antibodies intranasally daily for up to 28 days. Therefore, the benefit of the treatment in their study (61) could have been due to neutralization of the virus that had entered the body rather than blocking entry of the virus at the nasal mucosa. Similarly, a single intranasal monoclonal immunoglobin M (IgM) antibody administration in a mouse model of COVID-19 was highly efficacious when mice were infected with 10^4^ PFU (59). Human IgM systemic exposure was also noted in mice treated with human IgM monoclonal antibody anti-SARS-CoV-2, although the study attributed the protection to the persistent presence of the antibody at the nasal cavity for over 48 hours based on fluorescent tag measurement (59). Such long persistence of levels of IgM in the nasal cavity is at odds with other studies, including when using 99mTc-labeled albumin particles or fluorescently labeled IgY antibodies that showed residence time of 2-4 hours (40). If the long persistence is not an artifact of the method, it may suggest a unique benefit of IgM treatment as COVID-19 prophylaxis. Note, however, that with one exception (59), none of these studies assessed the cross-reactivity of antibodies against the variants.

Our work shows that, although the IgY was raised against the ancestral (index) strain RBD, the repertoire of the antibodies raised in hens was diverse and polyclonal so that binding affinities measured by ELISA for the single (Alpha), double (Delta), and triple amino acid (Beta) substitutions, or the Omicron variant with 15 amino acid substitutions in the RBD were similar to the affinity for the index RBD or full-length S protein (**Figure 4C, H**). We then confirmed that there was no difference between Alpha, Beta, and Delta variants, and the index strain in a neutralization assay in culture using a VSV-S pseudovirus or live virus (**Figure 4 D, E, I**), whereas a reduced neutralization activity of human serum was observed in side-by-side experiments (**Figure 4F, G**). The viral neutralization studies reported here were conducted by a commercial provider (RetroVirox) and by an established laboratory at the USAMRIID, both comparing the results with either convalescent sera or an immunized human, for relative titer evaluation.

The culture neutralization titer of anti-SARS-CoV-2 RBD IgY is lower than the human anti-SARS-CoV-2 sera (e.g., **Figures 4F vs. 4D**). However, this may reflect the need for protease-induced RBD exposure in the full-length S protein for binding by IgY, which might not occur effectively in the culture model. A comparison of titer values between our product and sera from an immunized person can also be calculated based on values of IgG levels in human sera (∼15 mg/mL); a titer of 1:2,000 (**Figure 4F**; dashed line) is equivalent to ∼7 ug/mL or 100-fold higher IgG titer than our IgY (**Figure 4D**; ∼600 ug/mL). However, as the dose of the IgY anti-SARS-CoV-2 preparation in humans is planned to be 4 mg/dose, more important is the equal potency of the IgY towards the various variants when used even at ∼1/10 of the intended IgY dose (**Figure 4E, I**).

Another potentially important difference between our findings and the previously published study is the antigen used to raise the antibodies. When expressed in mammalian cells, the full-length S1 protein has at least 22 glycosylation sites per S monomer (62). As glycosylated amino acids are more immunogenic, the affinity of the human antisera may reflect binding to the glycosylated determinants of the protein. However, as glycosylation sites in the S1 protein are heavily mutated and new sites may be formed in many of the variants (31), immune reactivity that is biased towards glycosylated sites may lead to loss of activity as the virus mutates. This will not occur when the non-glycosylated RBD is used as the immunogen, as we have done in our study using cell-free expressed RBD (23, 24). Supporting the negative impact of glycosylated antigen, increased immunogenicity of protein antigens after removal of glycosylation sites has been previously shown for hepatitis C virus envelope antigen-based vaccines (63). Furthermore, the apparent higher titer in the neutralization assay may be biased if the tested virus has the same glycosylation sites as that used as an immunogen in vaccinated individuals; many studies use the original viral isolates rather than the common current variants.

An important feature of our product is the ease of developing prophylaxis that can be quickly and inexpensively produced. We found that 24 mg total daily dose (divided into three equal doses) of intranasal anti-SARS-CoV-2 IgY for 14 days had an excellent safety profile in humans; this daily dose represents ∼1/20 of one egg of immunized SPF hen and ∼1/5 of an egg of commercial hen, underscoring that such an IgY dose is feasible for both production cost and effort.

There are some limitations to our studies. Our phase 1 clinical trial in healthy volunteers was to assess initial safety, tolerability, and PK of anti-SARS-CoV-2 IgY and was not designed to evaluate efficacy. In addition, we were unable to obtain in vivo data showing viral neutralization (**Supplementary Figure S2**). This may reflect using too much virus in this animal model of COVID- 19; our study used 8 x 10^5^ or 4 x 10^6^, vs. 1 or 5 x 10^4^ TCID50 (58, 60). In addition, our intranasal formulated IgY preparation was viscous to obtain better delivery in humans. As hamsters are obligatory nose-breathers, they may have blown out the formulated IgY. Finally, the virus that was delivered in 50 µL of liquid directly into each nare may have washed out some antibodies. Another limitation in our study is that the neutralization studies comparing the hen IgY vs. human sera were not comprehensive and included only 1 or 2 human samples. Nevertheless, our study is the first to demonstrate that anti-SARS-CoV-2 IgY against all the current variants of concern had a favorable safety profile when used chronically as intranasal drops in rats and humans.

There are also several advantages for the use of IgY as prophylaxis against other pathogens besides SARS-CoV-2 that cause disease in humans. As we noted above, IgY generation is inexpensive and fast; one egg of an SPF hen produces 20-80 daily doses (at 6 mg/dose) within 3 weeks from the first injection (1 week after the first boost). We found a limited variability between individual immunized hens as determined by ELISA and Western blot analyses and a batch-to-batch consistency (**Figure 3A-F**). IgY is also easy to distribute; besides the known long-term stability of purified IgY (64), we also found excellent stability of the formulated material at 2-8°C for at least 6 months (maximum time point measured so far) and greater than 2 weeks when stored at room temperature. This is in contrast to vaccines, some of which require cold-chain storage at -80°C that complicates the coordination of global distribution and to resource-poor regions, in particular.

Another important advantage of IgY-based prophylaxis is the ease of local production, including in low- and middle-income countries. As hen immunization is a standard procedure around the world, IgY purification and formulation do not require expensive equipment and are simple to conduct. In a separate study, we developed a step-by-step protocol for IgY purification for two settings in low- and middle-income countries where electricity is available using inexpensive, readily available materials in place of costly specialized laboratory equipment and chemicals (65). Therefore, anti-SARS-CoV-2 IgY can be readily made available worldwide as an additional means of reducing SARS-CoV-2 infection. Each day, one immunized hen can produce the daily dose required for prophylaxis of a family. In addition, by reducing viral mobility and anchoring the virus to the nasal mucus, transmission of the virus from infected to healthy individuals may be reduced.

## 5. Conclusion

The current COVID-19 pandemic illustrates the need for prophylactics that can be produced rapidly at low cost, are technically accessible anywhere in the world, and complement traditional vaccine development. The safety and benefit of IgY and the ease to produce it at low cost are well described for animal farms. In contrast, the clinical adaptation of IgY for human use has been slow, likely hampered by a lack of economic benefit that has hindered commercial development by industry. For that reason, we undertook the effort of establishing the ability of anti-SARS-CoV-2 IgY to neutralize variants of concern and the initial safety of the IgY preparation using industry GLP and GMP standards. We suggest that until vaccination that is highly effective against prevalent variants becomes available worldwide or herd immunity is achieved, intranasal delivery of anti-SARS-CoV-2 IgY may provide passive immunization, including for use as an add-on to personal protective equipment and other preventive measures for the general population. This IgY may also provide short-term protection in addition to vaccines in less well-ventilated environments, such as trains, airplanes, and lecture halls. We also suggest that this approach has the potential to provide a means to curb new threats of epidemics by airborne infectious agents; by providing the relevant immunogen for hen immunization at the geographical site where the threat was detected, an effective passive immunity can be initiated locally to stop the spread of the airborne infectious agent before it becomes an epidemic. We hope that this study will trigger further work to evaluate the safety and efficacy of anti-SARS-CoV-2 IgY in those at risk of SARS-CoV-2 infection.

## Data Availability

The original contributions presented in the study are included in the article/supplementary material. Further inquiries can be directed to the corresponding author.

## Author Contributions

Contributions were made by MW, DM, and LF to project conceptualization; LF, ML, CS, NO, AY, TH, AY, KB, CC, JD, BM, AB, and TS to methodology; ML, NO, GY, CC, CB, SA, TB, AJ, SP, BK, and LL to the investigations; LL and DM to visualization; DM and SK to funding acquisition; MA, RB, LL, JR to project administration; JR, JD, BM, TH, AB, SK, and DM to supervision; DM and LF to writing the original draft; and DM, LF, BK, and TS to editing. All authors reviewed and approved the submitted version.

## Acknowledgments

We thank our many SPARK at Stanford advisors for their support, both financially and through their invaluable advice. We also thank Miao Wen, Nina A. Carlos, Lawrence Huang, Cuong Tran, Stephanie Armstrong, and Daniel Calarese of Sutro Biopharma for their help in the production of the cell-free expressed RBD, Roxanne Lopardo at Avian Vaccine Services, Charles River Laboratories for her work in IgY testing, and David Goodkin for helpful advice and critical review. We also thank the study staff at Linear Clinical Research Ltd and Resolutum Global for their outstanding conduct and analysis of the phase 1 clinical study and Yael Rosenberg-Hasson, Tyson H Holmes, Prof. Mark Davis, and other members of the Immunoassay Team at the Human Immune Monitoring Center (Stanford University) for cytokine analyses and statistical consultation. Linda MacKeen provided invaluable editing support. This manuscript has previously appeared online as a preprint (66).

## Disclaimer

Opinions, conclusions, interpretations, and recommendations are those of the authors and are not necessarily endorsed by the U.S. Army. The mention of trade names or commercial products does not constitute endorsement or recommendation for use by the Department of the Army or the Department of Defense.

## Abbreviations

BSA, bovine serum albumin; COVID-19, coronavirus disease 2019; CPE, cytopathic effect; DMEM, Dulbecco’s Modified Eagle Medium; ELISA, enzyme-linked immunosorbent assay; FBS, fetal bovine serum; GLP, Good Laboratory Practice; GMP, Good Manufacturing Practice; hACE2, human angiotensin-converting enzyme 2; HPLC, high-performance liquid chromatography; HRP, horseradish peroxidase; IC50, half-maximal inhibitory concentration; IgA, immunoglobulin A; IgE, immunoglobulin E; IgG, immunoglobulin G; IgM, immunoglobulin M; IgY, immunoglobulin Y; MEM, Minimum Essential Media; NT50, 50% neutralization titer; PBS, phosphate-buffered solution; PFU, plaque-forming units; PK, pharmacokinetic; RBD, receptor-binding domain; RLU, relative light unit ratio; RU, replication units; S, spike; SARS-CoV-2, severe acute respiratory syndrome coronavirus 2; SDS-PAGE, sodium dodecyl sulfate-polyacrylamide gel electrophoresis; SPF, specific pathogen-free; SMC, Safety Monitoring Committee; SUMO, small ubiquitin-related modifier; TCID50, tissue culture infective dose; USAMARIID, United States Army Medical Research Institute of Infectious Diseases; VSV, vesicular stomatitis virus; VSV-S, VSV expressing spike protein on its surface

## Supplementary Material

**Supplementary Table S1.**
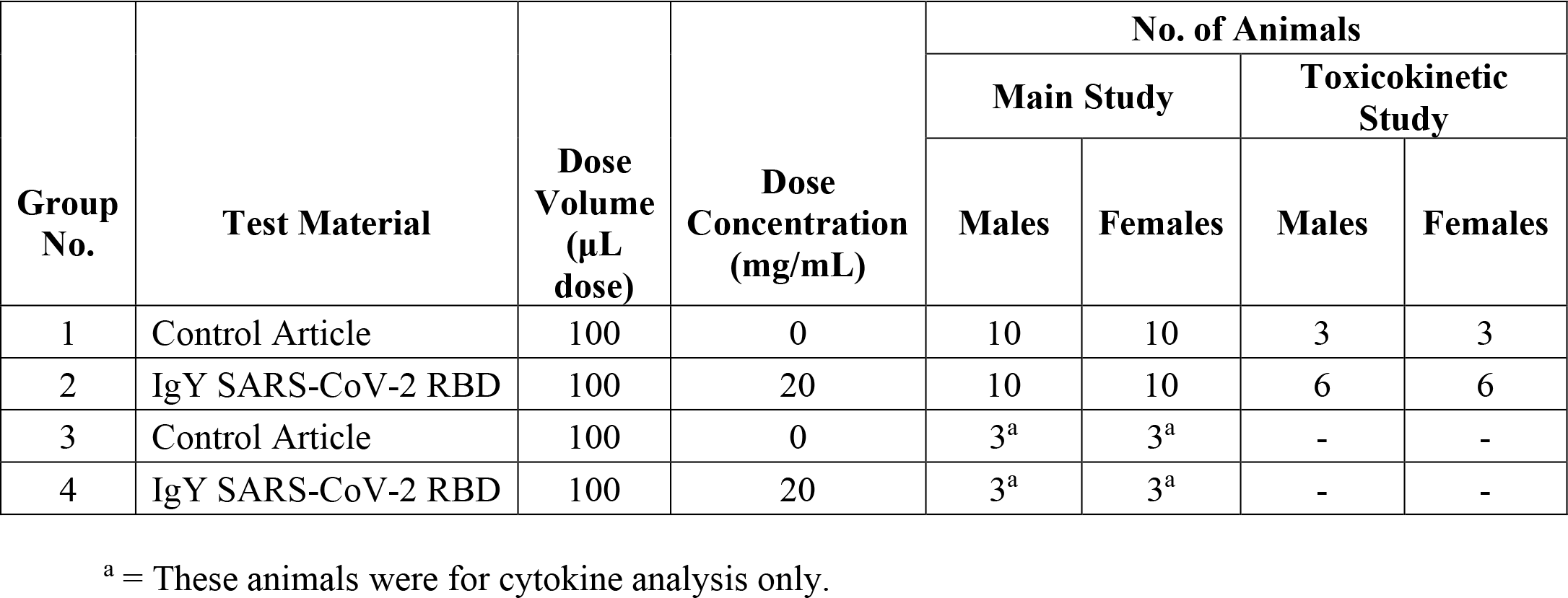
Experimental protocol of the rat toxicity study

| Group No. | Test Material | Dose Volume (µL dose) | Dose Concentration (mg/mL) | No. of Animals |  |  |  |
| --- | --- | --- | --- | --- | --- | --- | --- |
|  |  |  |  | Main Study |  | Toxicokinetic Study |  |
|  |  |  |  | Males | Females | Males | Females |
| 1 | Control Article | 100 | 0 | 10 | 10 | 3 | 3 |
| 2 | IgY SARS-CoV-2 RBD | 100 | 20 | 10 | 10 | 6 | 6 |
| 3 | Control Article | 100 | 0 | 3<sup>a</sup> | 3<sup>a</sup> | - | - |
| 4 | IgY SARS-CoV-2 RBD | 100 | 20 | 3<sup>a</sup> | 3<sup>a</sup> | - | - |

<sup>a</sup> = These animals were for cytokine analysis only.

**Supplementary Table S2.**
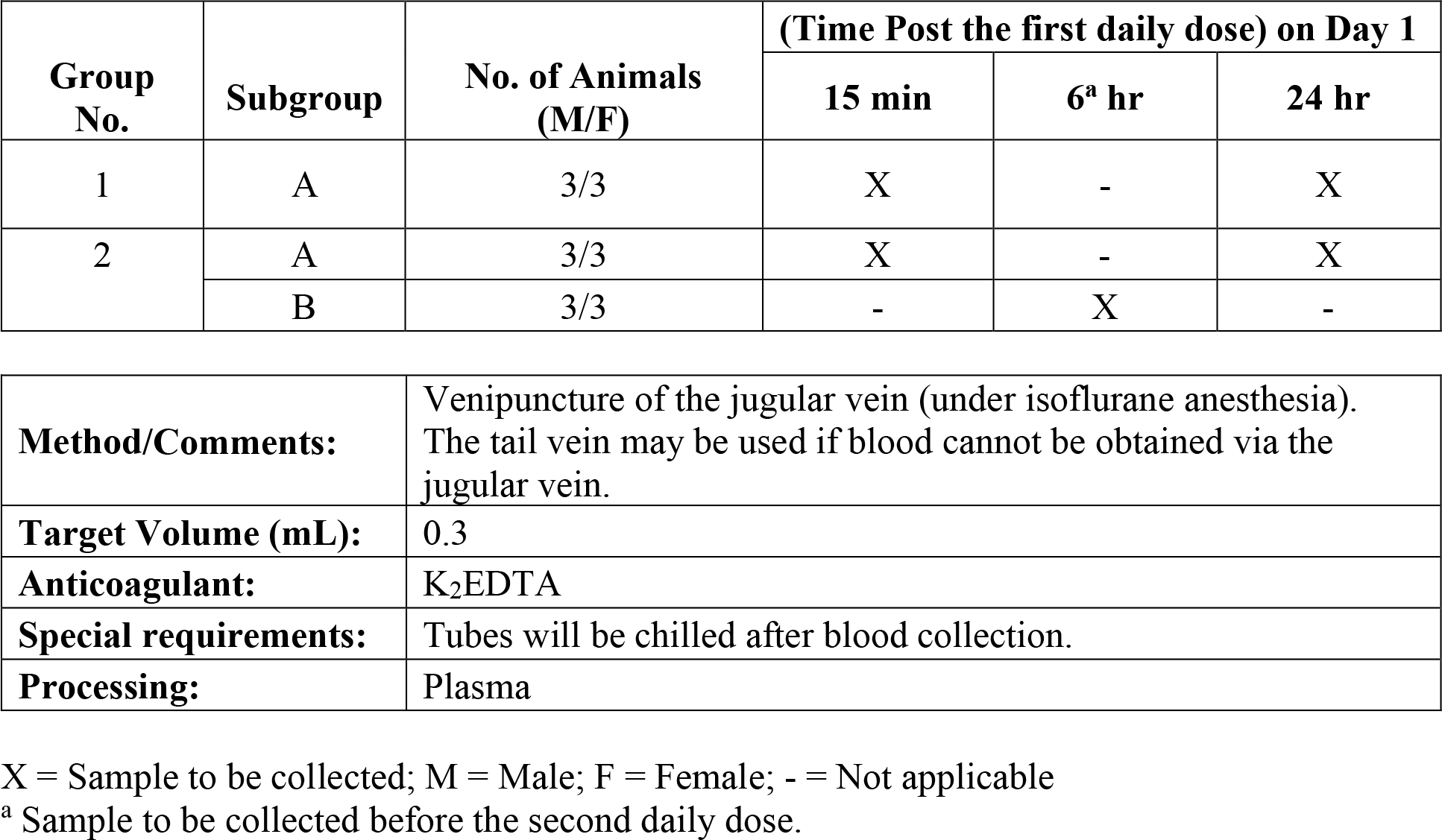
Bioanalytical sample collection from the rat toxicity study

| Group No. | Subgroup | No. of Animals (M/F) | (Time Post the first daily dose) on Day 1 |  |  |
| --- | --- | --- | --- | --- | --- |
|  |  |  | 15 min | 6<sup>a</sup> hr | 24 hr |
| 1 | A | 3/3 | X | - | X |
| 2 | A | 3/3 | X | - | X |
|  | B | 3/3 | - | X | - |

|  |  |
| --- | --- |
| **Method/Comments:** | Venipuncture of the jugular vein (under isoflurane anesthesia).<br>The tail vein may be used if blood cannot be obtained via the jugular vein. |
| **Target Volume (mL):** | 0.3 |
| **Anticoagulant:** | K<sub>2</sub>EDTA |
| **Special requirements:** | Tubes will be chilled after blood collection. |
| **Processing:** | Plasma |

X = Sample to be collected; M = Male; F = Female; - = Not applicable
<sup>a</sup> Sample to be collected before the second daily dose.

**S3 Table.**
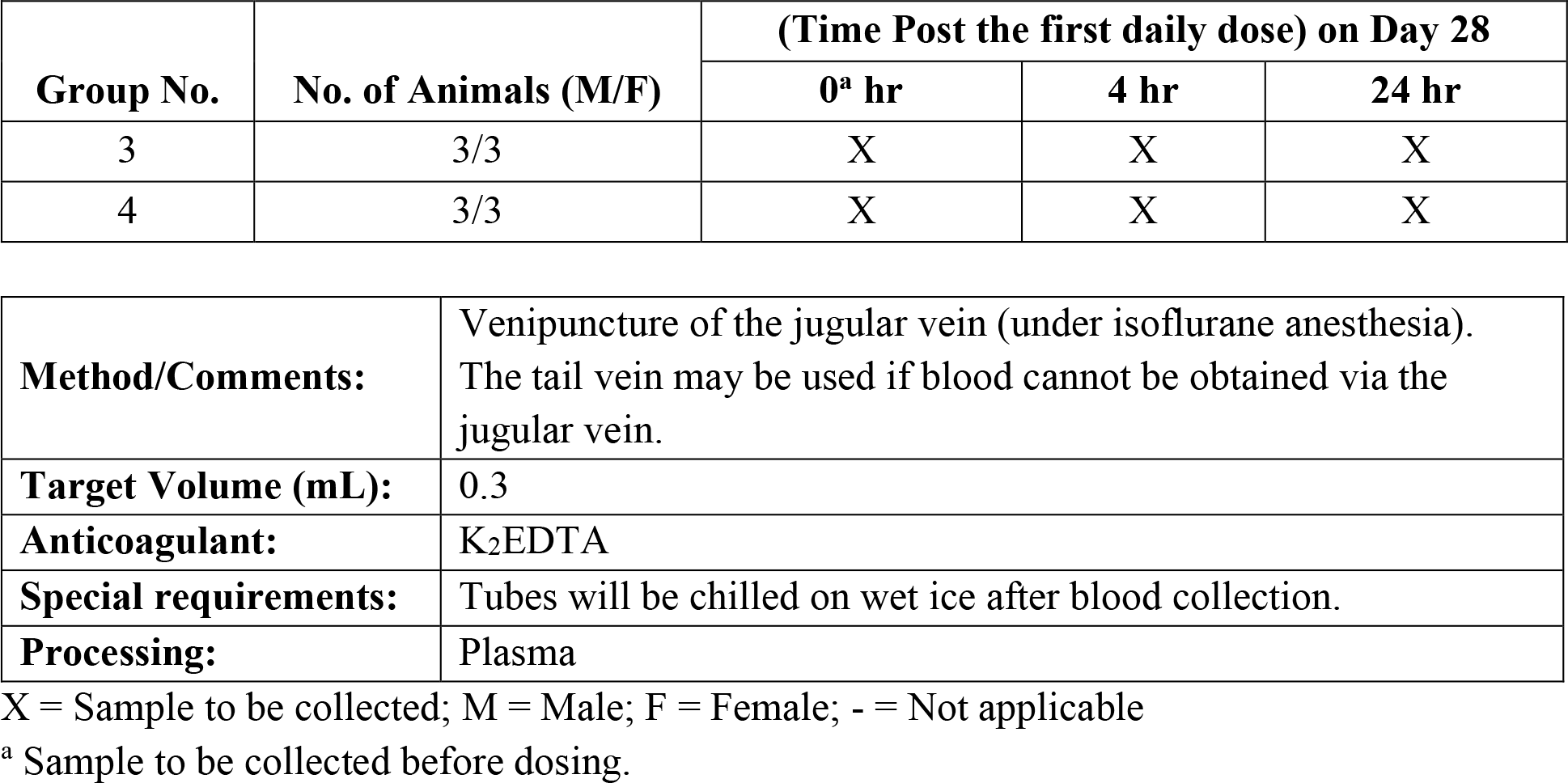
Cytokine sample collection for the rat toxicity study

| Group No. | No. of Animals (M/F) | (Time Post the first daily dose) on Day 28 |  |  |
| --- | --- | --- | --- | --- |
|  |  | 0<sup>a</sup> hr | 4 hr | 24 hr |
| 3 | 3/3 | X | X | X |
| 4 | 3/3 | X | X | X |

|  |  |
| --- | --- |
| **Method/Comments:** | Venipuncture of the jugular vein (under isoflurane anesthesia).<br>The tail vein may be used if blood cannot be obtained via the jugular vein. |
| **Target Volume (mL):** | 0.3 |
| **Anticoagulant:** | K<sub>2</sub>EDTA |
| **Special requirements:** | Tubes will be chilled on wet ice after blood collection. |
| **Processing:** | Plasma |

X = Sample to be collected; M = Male; F = Female; - = Not applicable
<sup>a</sup> Sample to be collected before dosing.

**Supplementary Table S4.**
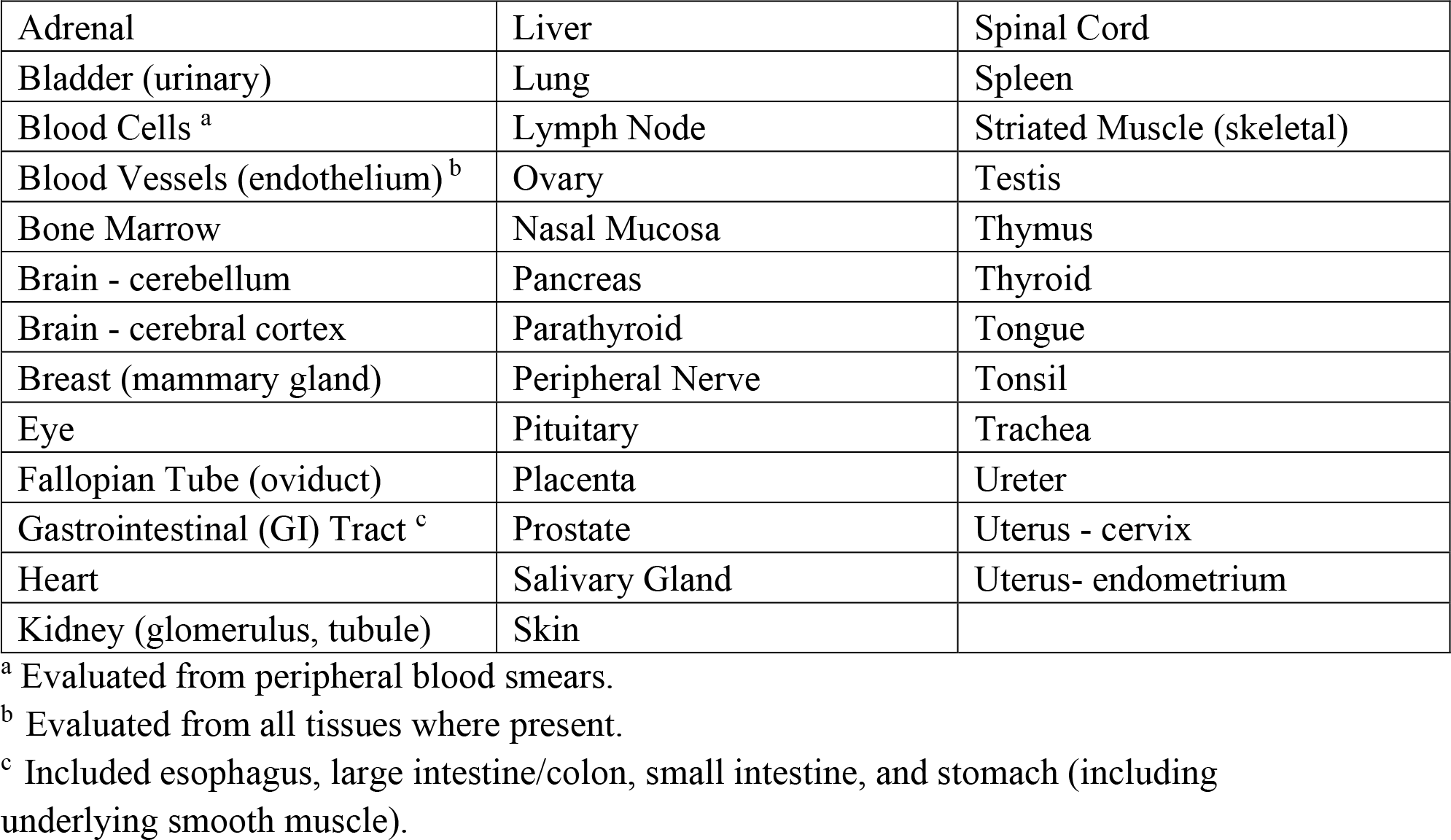
Human tissue cross-reactivity; list of tissues examined. Binding of anti-SARS-CoV-2 RBD IgY was determined using the following normal human tissue from 3 separate donors.

|  |  |  |
| --- | --- | --- |
| Adrenal | Liver | Spinal Cord |
| Bladder (urinary) | Lung | Spleen |
| Blood Cells <sup>a</sup> | Lymph Node | Striated Muscle (skeletal) |
| Blood Vessels (endothelium) <sup>b</sup> | Ovary | Testis |
| Bone Marrow | Nasal Mucosa | Thymus |
| Brain - cerebellum | Pancreas | Thyroid |
| Brain - cerebral cortex | Parathyroid | Tongue |
| Breast (mammary gland) | Peripheral Nerve | Tonsil |
| Eye | Pituitary | Trachea |
| Fallopian Tube (oviduct) | Placenta | Ureter |
| Gastrointestinal (GI) Tract <sup>c</sup> | Prostate | Uterus - cervix |
| Heart | Salivary Gland | Uterus- endometrium |
| Kidney (glomerulus, tubule) | Skin |  |

<sup>a</sup> Evaluated from peripheral blood smears.
<sup>b</sup> Evaluated from all tissues where present.
<sup>c</sup> Included esophagus, large intestine/colon, small intestine, and stomach (including underlying smooth muscle).

**Supplementary Figure S1.**
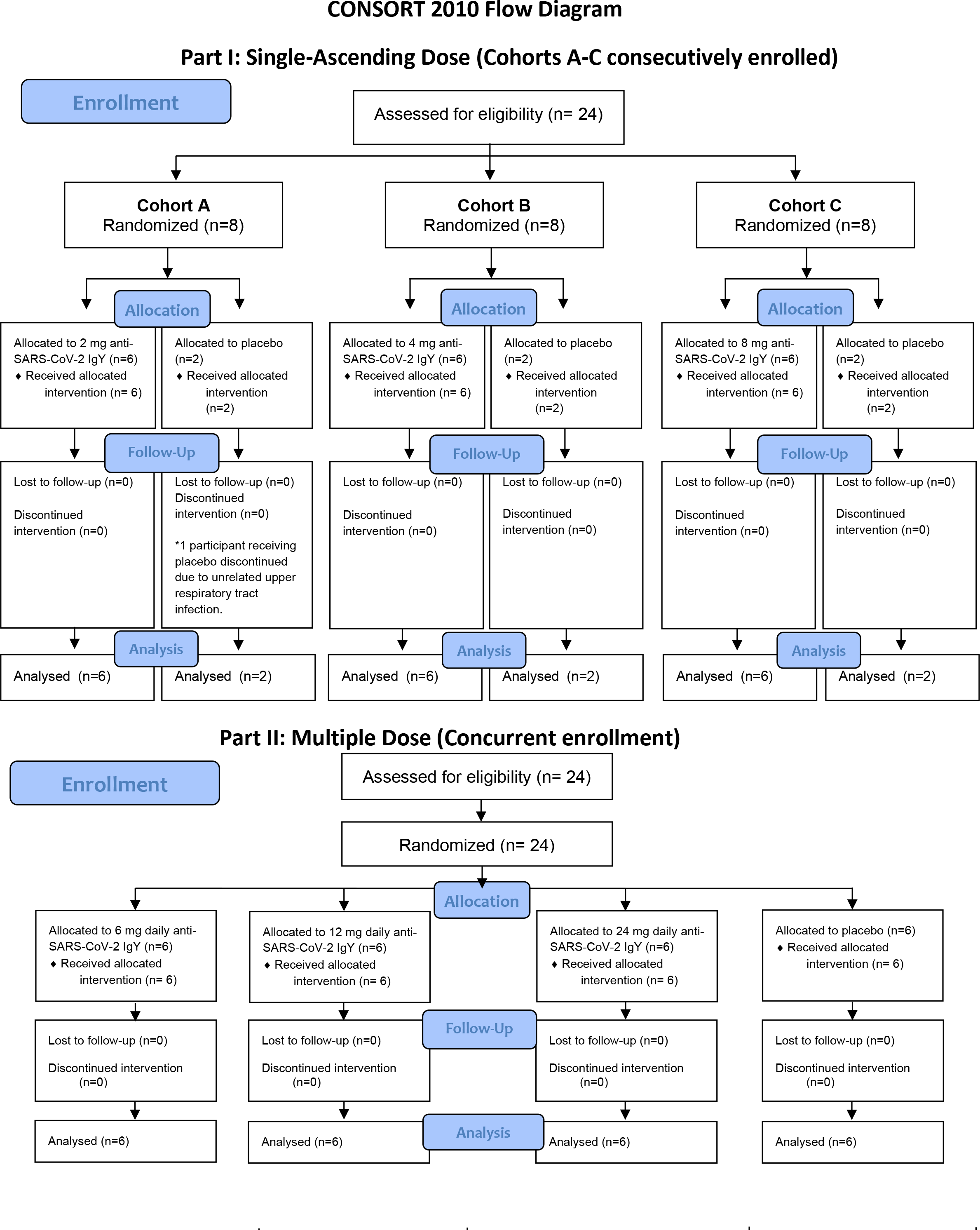
CONSORT flow diagram of phase 1 single-ascending and multiple-dose study.

**Supplementary Figure 2.**
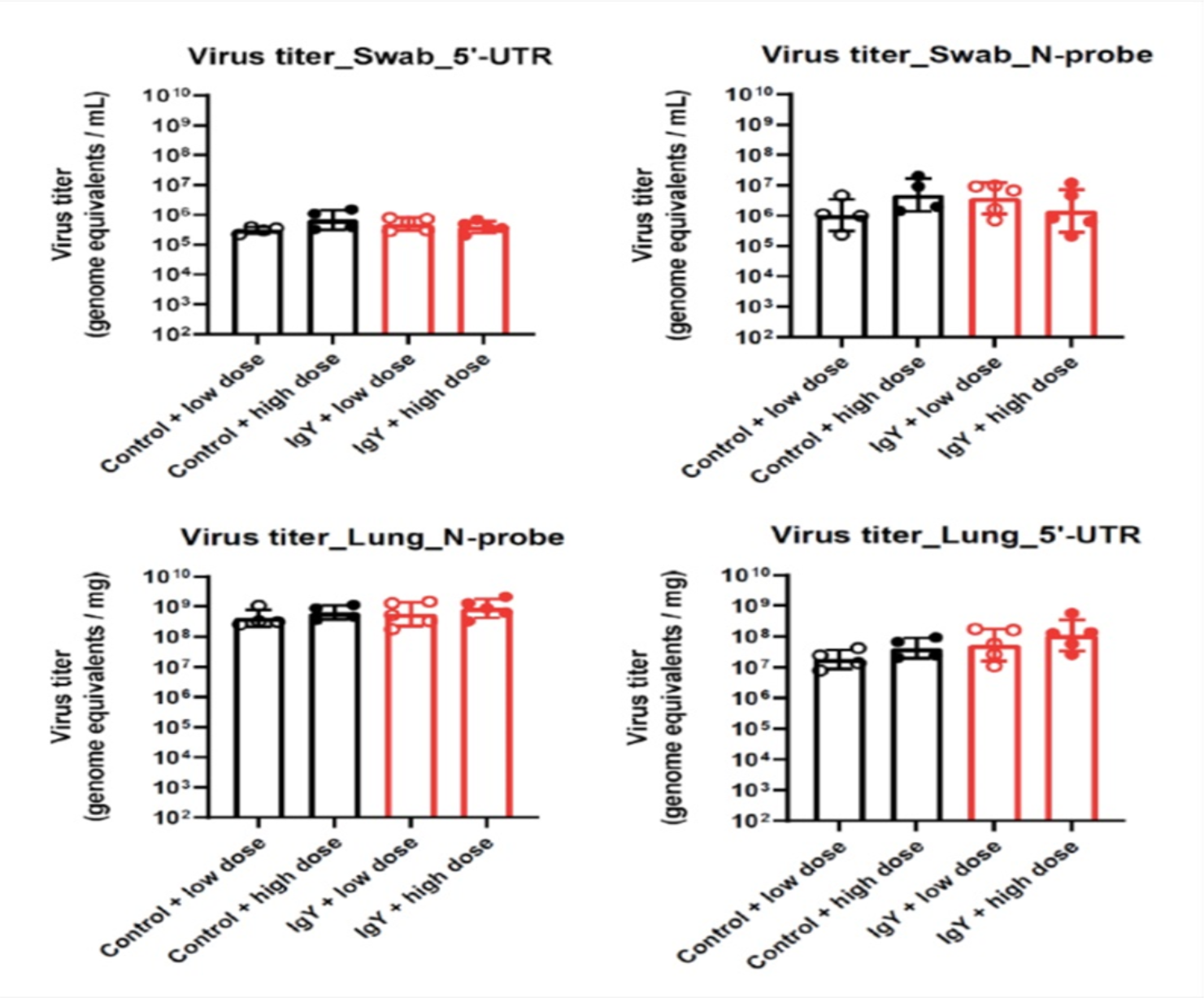
Hamster efficacy study Placebo control and IgY-treated Syrian hamsters were challenged with a high (4 x 10^6^) or ‘low’ (8 x 10^5^) dose of SARS-CoV-2. Infectious virus titers were measured in the lungs 3 days post-infection by plaque assay. Each symbol is a single animal. One animal in the placebo group challenged with a low dose of the virus did not have any detectable virus in the lung homogenate. This was ruled an outlier based on the viral RNA levels in other assays. Viral RNA levels were quantified RT-PCR in lung homogenates of nasal swabs using 2 different primer-probe sets. Each symbol is a single animal and the bars represent the geometric mean with geometric standard deviation.

**Supplementary Figure S3.**
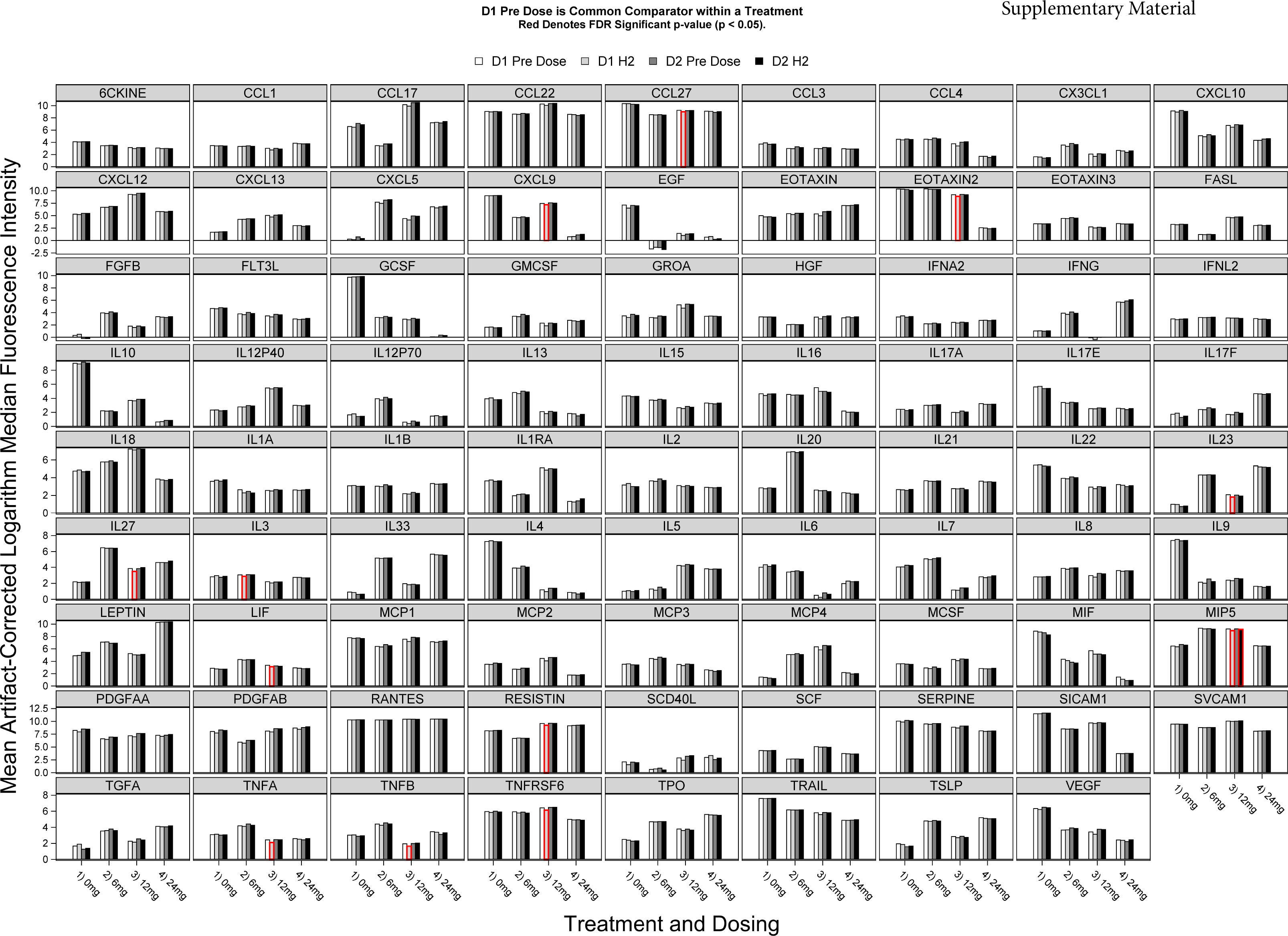
(next page). Serum cytokines for human participants in the phase 1 study. Blood cytokine levels before treatment (D1 pre-dose), 2 hours after the first treatment (D1H2), 24 hours after 3-times daily intranasal (D2 pre-dose), and 2 hours after the first dose on the second day (D2H2) of anti-SARS-CoV-2 RBD IgY (0, 2, 4 and 8 mg/dose or 0, 6, 12, and 24 mg/day). D1 pre-dose is common comparator within a treatment. Red denotes false discovery rate (FDR) significant p-value (p<0.05).

**Supplementary Methods.** Protocol for “Phase 1 Study in Healthy Participants to Evaluate the Safety, Tolerability, and Pharmacokinetics of Single-Ascending and Multiple Doses of an Anti-Severe Acute Respiratory Syndrome Coronavirus 2 (SARS-CoV-2) Chicken Egg Antibody (IgY)”

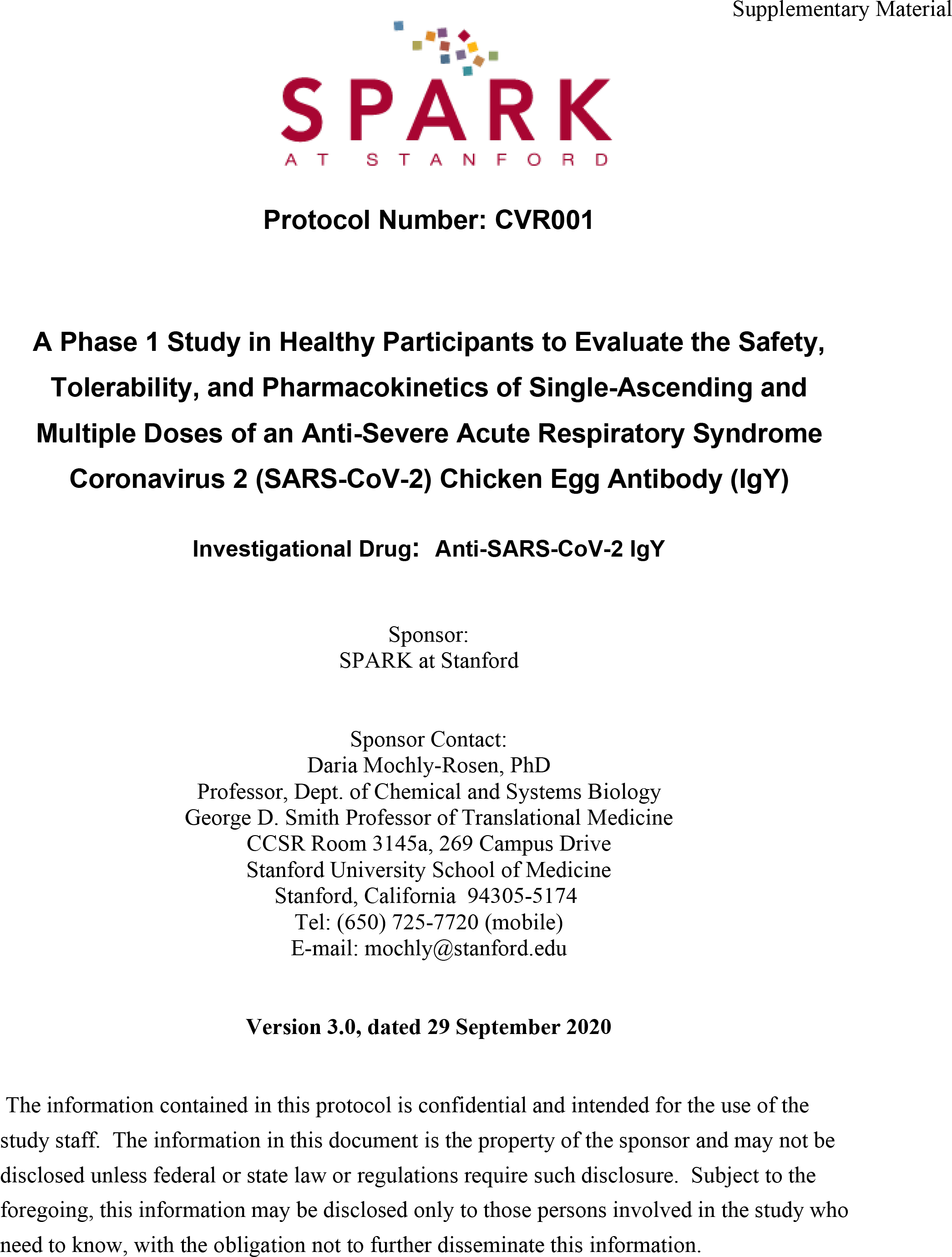
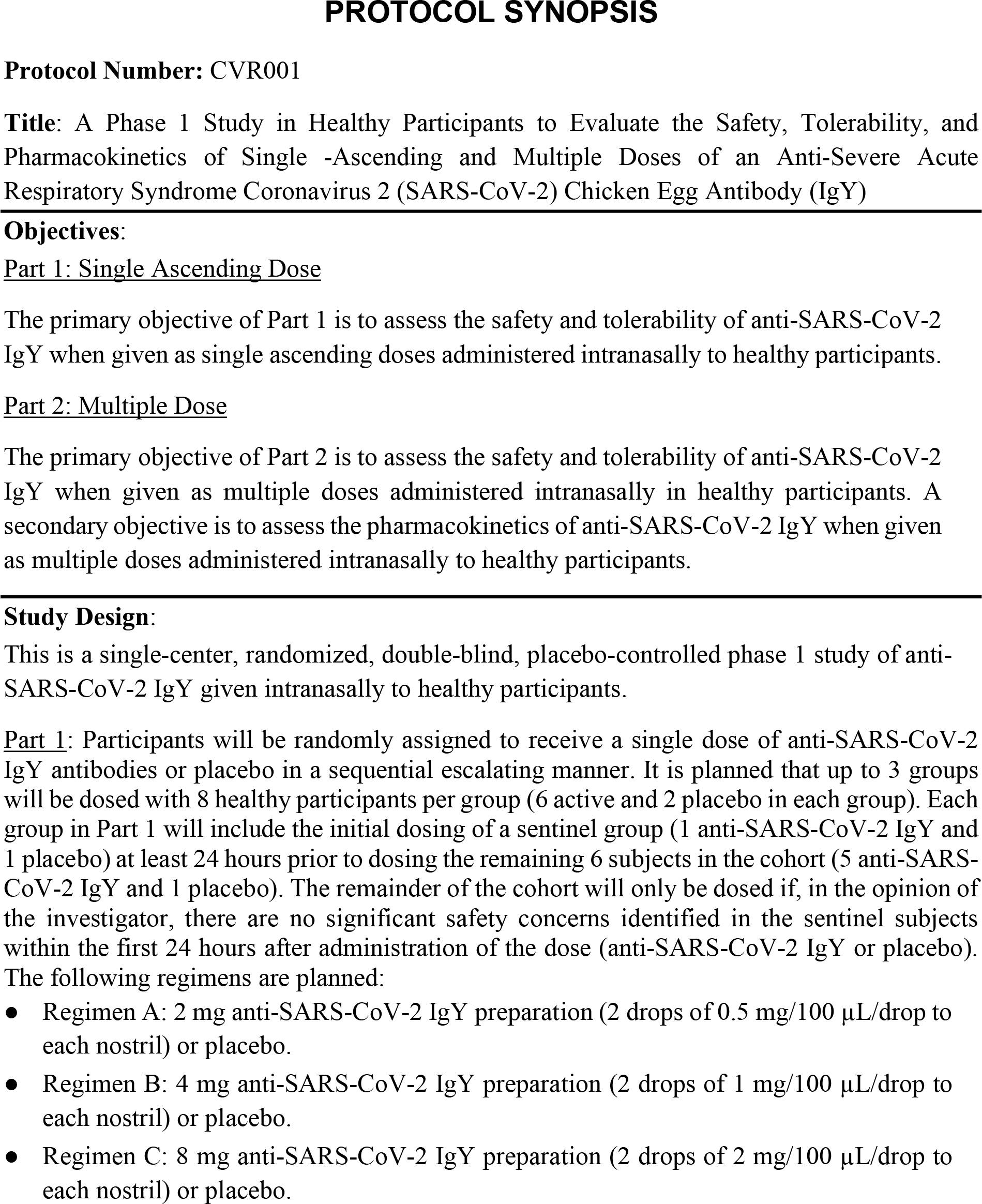
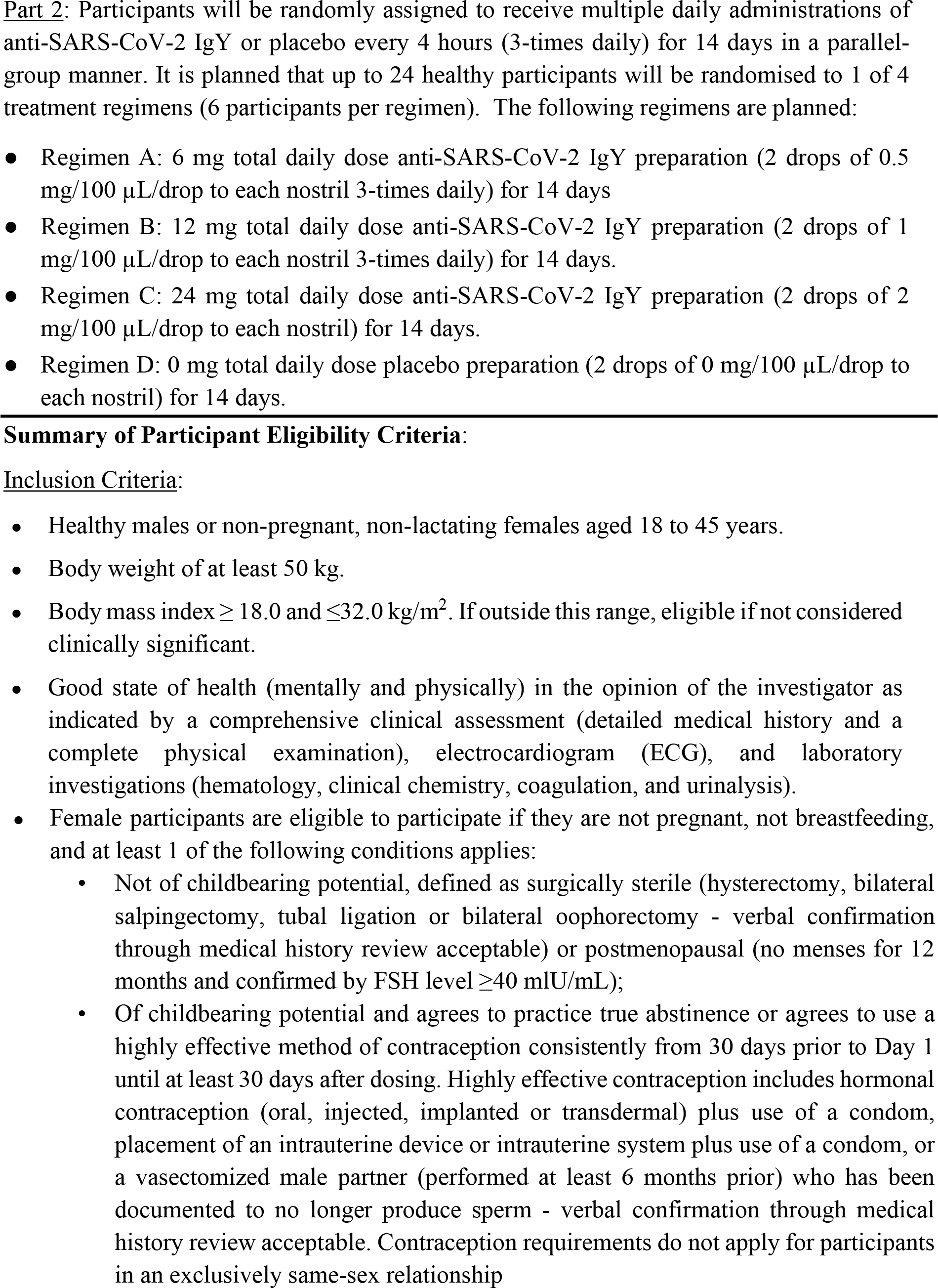
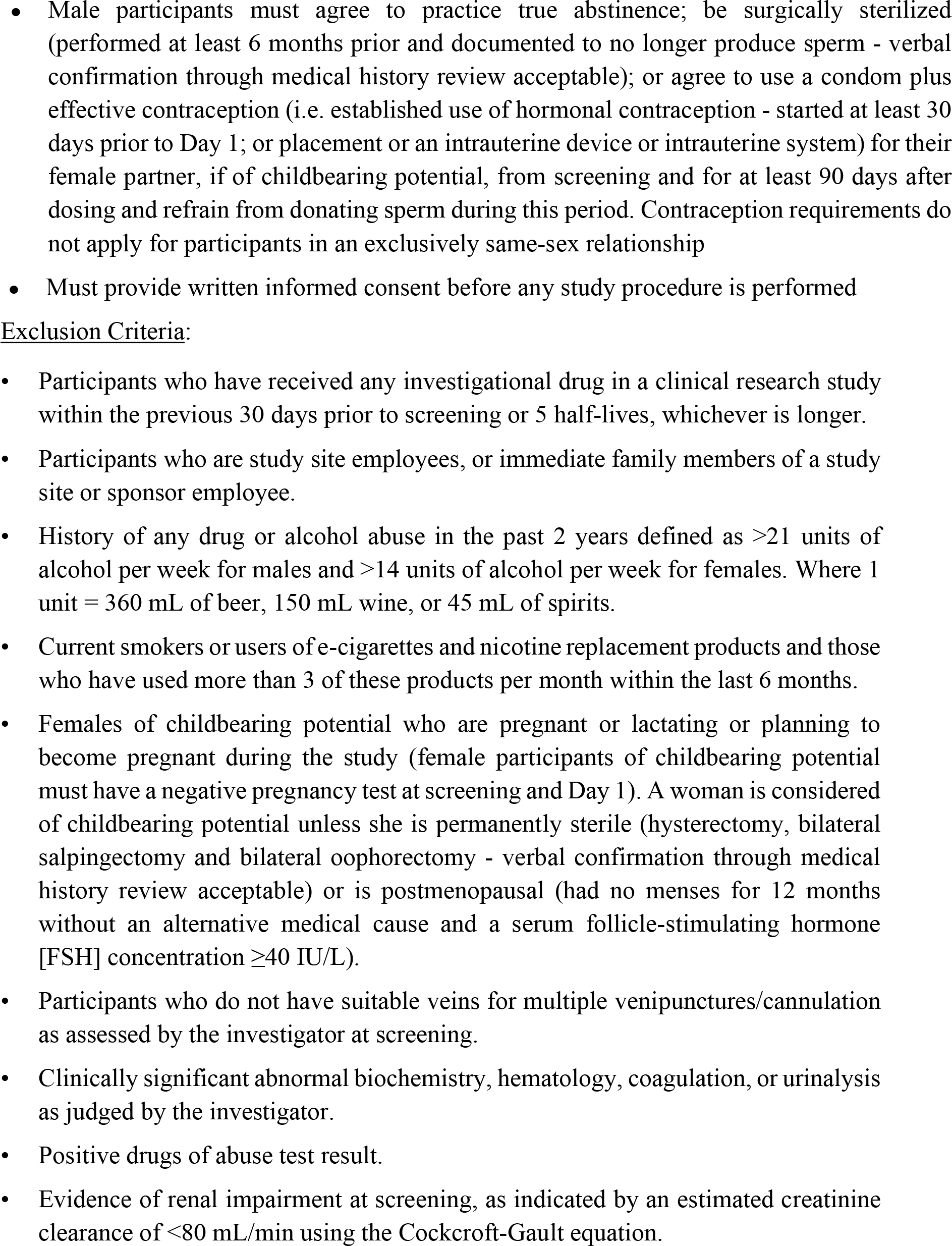
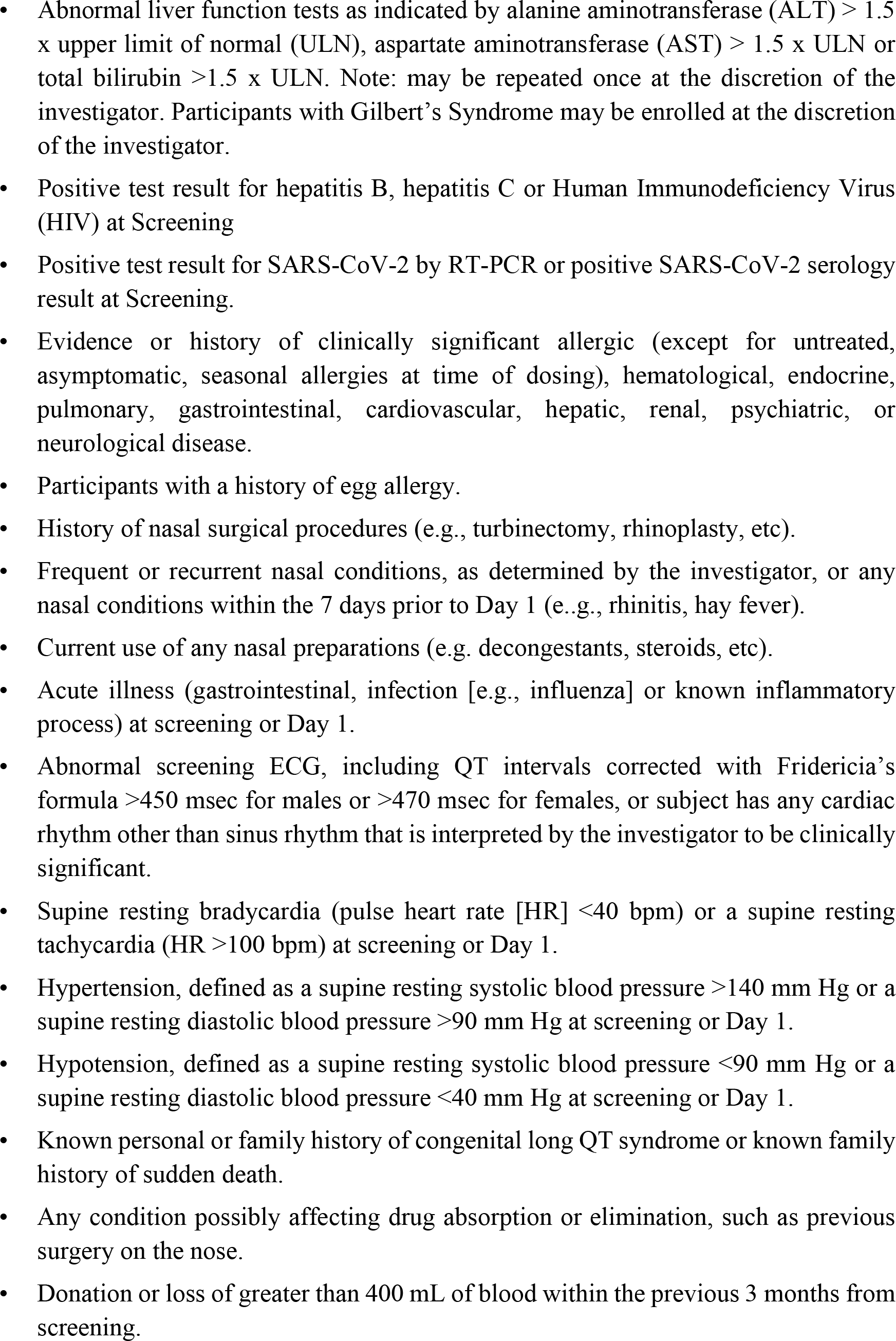
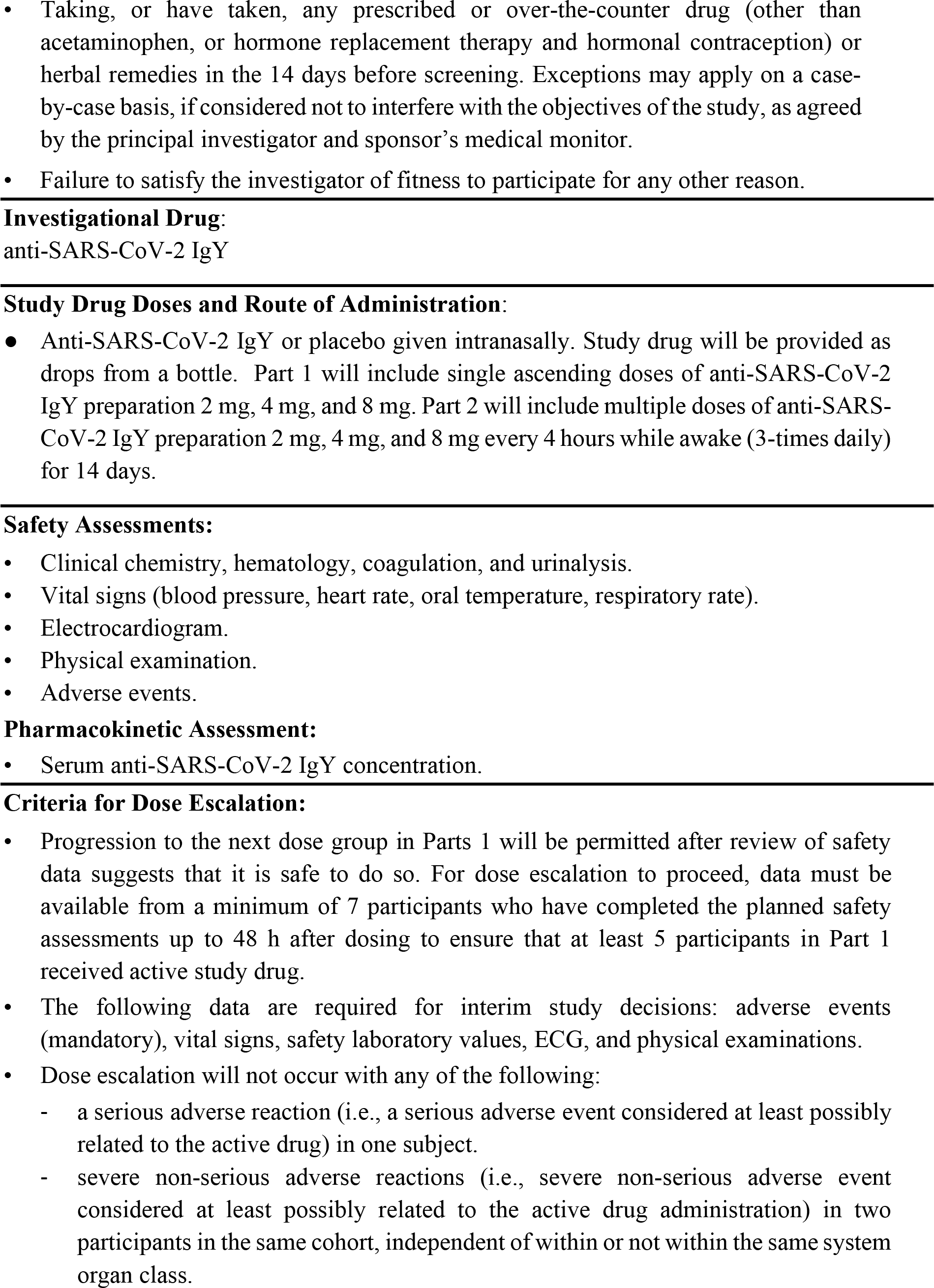
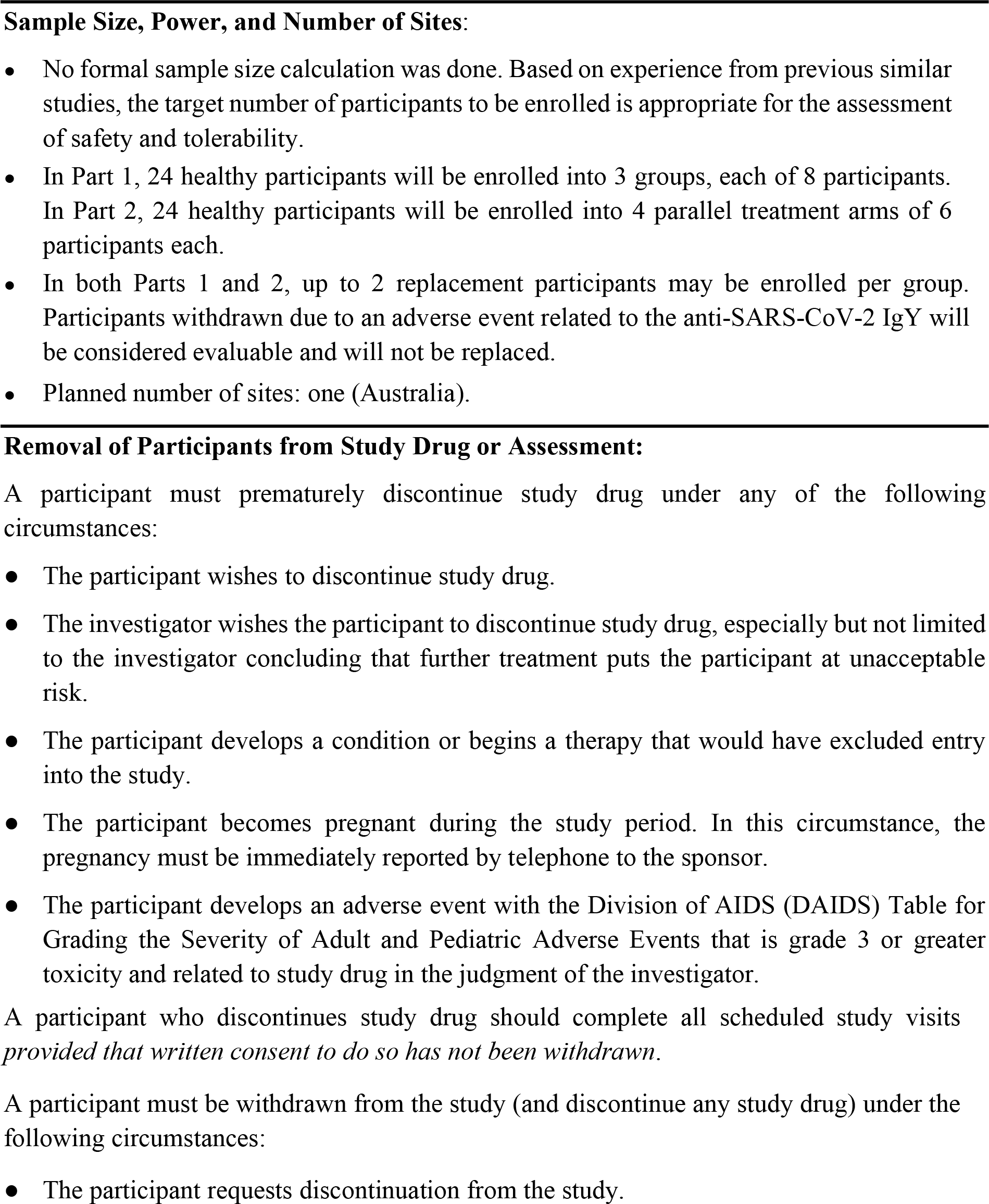
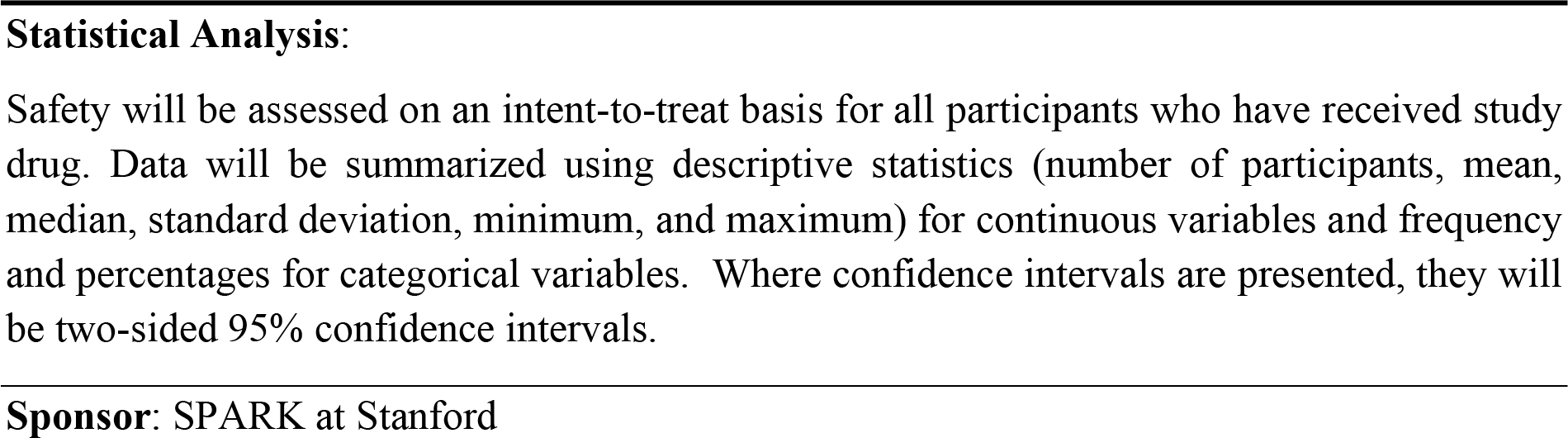

## 1. LIST OF ABBREVIATIONS

### ABBREVIATION DEFINITION

ACE2: angiotensin-converting enzyme 2
AE: adverse event
ALT: alanine transaminase
AST: aspartate transaminase
BUN: blood urea nitrogen
CLIA: Clinical Laboratory Improvement Amendments
COVID-19: coronavirus disease 2019
CRF: case report form
DAIDS: Division of AIDS
EC: Ethics Committee
FSH: follicle-stimulating hormone
hCG: human chorionic gonadotropin
HIPAA: Health Insurance Portability and Accountability Act
HIV: human immunodeficiency virus
IgG: immunoglobulin G
IgM: immunoglobulin M
IgY: immunoglobulin Y
INR: international normalized ratio
ITT: intent-to-treat
MedDRA: Medical Dictionary for Regulatory Activities
Mg: milligram
mL: milliliter
μL: microliter
PT: prothrombin time
aPTT: activated partial thromboplastin time
rt-PCR: reverse transcription-polymerase chain reaction
SAE: serious adverse event
SARS-CoV-2: severe acute respiratory syndrome coronavirus 2
SDS-PAGE SMC: sodium dodecyl sulfate-polyacrylamide gel electrophoresis Safety Monitoring Committee
WHO: World Health Organization

## 2. INTRODUCTION

### 1.1 Severe Acute Respiratory Syndrome Coronavirus 2 (SARS-CoV-2) Infection

Coronaviruses are a large family of single-stranded RNA viruses that infect mammals and birds, causing respiratory infection. In December 2019, a novel coronavirus was identified as the cause of a cluster of pneumonia cases in Wuhan, China. Its rapid spread resulted in an epidemic throughout China that was followed by a dramatically increased number of cases worldwide. In February 2020, the World Health Organization (WHO) designated COVID-19 (coronavirus disease 2019) to be caused by a novel coronavirus termed severe acute respiratory syndrome coronavirus 2 (SARS-CoV-2) (World Health Organization 2020). As of August 3, 2020, over 18 million persons with COVID-19 had been identified in 215 countries with an estimated 692,000 deaths (World Health Organization 2020). The progression from isolated cases in China in late 2019 to the current worldwide pandemic is underscored by asymptomatic (Chan et al., 2020; Li et al., 2020; Mizumoto and Chowell, 2020) and presymptomatic (Arons et al., 2020; He et al., 2020; Kimball et al., 2020) infection and transmission, in addition to potential transmission through environmental contamination (Ong et al., 2020). Viral entry is most prominent through the nasal passage, where binding of the viral spike protein to the human angiotensin-converting enzyme 2 (ACE2) receptor facilitates cell entry (Sungank et al., 2020; Zhou et al., 2020). The ACE2 receptor is localized on mucosal epithelial cells (Hamming et al., 2004; Jia et al., 2006). There is currently no drug or vaccine that prevents or reduces the risk of acquiring SARS-CoV-2 infection.

### 1.2 Immunoglobulin Y

Antibody-based passive immunization with immunoglobulin G (IgG) has a long history of effectiveness in both preventing and treating human infectious diseases (Keller and Stiehm, 2000). The possibility of protective immunity following infection with SARS-CoV-2 has led to uncontrolled studies in a small number of critically ill hospitalized patients with COVID-19 that suggest potential benefit from the effect of convalescent plasma containing neutralizing antibodies (Duan et al., 2020; Salazar et al., 2020; Shen et al., 2020). As such, the potential of passive immunization with egg yolk antibodies against SARS-CoV-2 infection has also been raised (Constantin et al., 2020; Somasundaram et al., 2020).

Egg yolk antibodies, termed immunoglobulin Y (IgY), are present in birds, reptiles, and amphibians, and considered the evolutionary precursor of immunoglobulins such as IgG present only in mammals (Kovas-Nolan and Mine, 2012). Immunoglobulin Y does not cross-react with mammalian antibodies or other unintentional targets or activate the mammalian complement system or Fc receptors (Warr et al., 1995; Zhang et al., 2017).

Immunoglobulin Y has been beneficial with favorable safety and tolerability when given prophylactically in both animal models and clinical settings of viral diseases, including respiratory infections (Kovas-Nolan and Mine, 2012). For example, animal studies have shown that the prophylactic treatment of calf and newborn piglets with orally administered IgY was highly effective and well tolerated against coronaviruses and rotavirus (Ikemori et al., 1997; Vega et al., 2011). Intranasal administration of IgY was beneficial and well tolerated in mouse models of influenza infection (Nguyen at al., 2010; Wallach et al., 2011; Yang et al., 2014) and *P. aeruginosa* acute pneumonia (Ranjbar et al., 2019), and a guinea pig model of allergic rhinitis (Guo-Zhu et al., 2015; Wei-Xu et al., 2016). Inhaled anti-*Pseudomonas* IgY were protective and well tolerated in a ventilated piglet model of *P. aeruginosa* infection (Otterbeck et al., 2019).

In humans, continuous prophylactic treatment by daily mouth rinse with specific IgY against. *P. aeruginosa* in 17 patients with cystic fibrosis for up to 12 years (114 patient-years) significantly reduced pulmonary *P. aeruginosa* infections compared with 23 cystic fibrosis control patients, with no adverse events (Kollberg et al., 2003; Nilsson et al., 2008). A randomized, double-blind, placebo-controlled phase 3 trial of 164 patients with cystic fibrosis conducted at 47 European sites from 2011 to 2015 evaluated nightly treatment with a gargling solution of anti- *P. aeruginosa* IgY (Schuster et al., 2019). No clinically meaningful differences in adverse events were reported between the treatment arms. Additional clinical studies evaluating IgY given orally include (a) treatment with a gel containing anti-*Candida* IgY, which improved tongue coating and the number of *Candida* colonies (Takeuchi et al., 2016),. (b) treatment with lozenges containing anti-*P. gingivalis* IgY, which improved gum pocket depth, gingival bleeding, and bacterial levels (Yokoyama et al., 2007; Nguyen et al., 2018), (c) ingestion of anti-rotavirus IgY, which reduced stool frequencies, viral excretion, and lowered hydration needs in children with rotavirus diarrhea (Sarker et al., 2001; Rahman et al., 2012), and (d) suppression of *H. pylori* after ingestion of yogurt containing anti-*H.pylori* IgY (Horie et al., 2004).

Overall, available data to date suggest that IgY given systemically do not have off-target inflammatory effects and are nontoxic to humans, thus permitting clinical applications in a wide range of vulnerable populations including the elderly, immunocompromised, and young children (Rahman et al., 2013).

### 2.3 Use of anti-SARS-CoV-2 IgY for Prevention of SARS-CoV-2 Infection

There are no published studies that have evaluated the use of anti-SARS-CoV-2 IgY to prevent human SARS-CoV-2 infection. Active vaccination requires the induction of an immune response that takes time to develop and varies depending on the vaccine and recipient. In contrast, passive antibody administration may provide immediate immunity to susceptible persons.

Passive immunization with sera from recovered patients infected with SARS-CoV-2 is currently under investigation and requires the availability of a population of donors who have recovered from the disease and can donate convalescent serum, blood banking facilities to process the serum donations, availability of assays to detect SARS-CoV-2 in serum, virologic assays to measure viral neutralization, virology laboratory support to perform these assays, and clinical protocols that are well designed and comply with country-specific and local regulatory approval (Casadevall and Pirofski, 2020). Passive immunization with anti-SARS-CoV-2 IgY may offer advantages in the known safety profile of IgY, applicability to a wide array of individuals, high yield per egg, and rapid mass production at a low cost given the large production of eggs for human consumption (Constantin et al., 2020; Somasundaram et al., 2020). In addition, anti-SARS-CoV-2 IgY would not be expected to elicit antibody-dependent enhancement of infection due to an inability to bind to human IgG Fc receptors on immune cells.

Intranasal antibody prophylaxis is an especially effective means against multiple viral pathogens (Hemmingsson and Hammarström, 1993; Giraudi et al., 1997; Heikkinen et al., 1998; Weltzin and Monath, 1999). Further, key characteristics of the mechanism of action of anti-SARS-CoV-2 IgY may be ideal for effective immunization: a) anti-SARS-CoV-2 IgY may bind to the spike protein on the surface of the virus, competing with the binding of the viral spike protein to the human ACE2 receptor to prevent cell entry and infection, b) anti-SARS-CoV-2 IgY may also agglutinate SARS-CoV-2 on the surface of the mucosa, thus preventing viral entry across the mucosa, and c) intranasal administration can deliver anti-SARS-CoV-2 IgY to the nasal passage and throat mucosa (through mucociliary clearance), the main routes of viral entry and replication.

In the absence of an active vaccine, this approach may be especially valuable for immediate and short-lived protection when used with personal protective equipment and other preventative measures to provide added protection for frontline health care personnel or other service workers at increased risk of infection.

Neutralization test results showed that SARS-CoV-2 IgY was effective in neutralizing SARS-CoV-2 (SPARK at Stanford, data on file). More detailed information about nonclinical and preclinical studies can be found in the Investigator’s Brochure.

## 2. OBJECTIVES

### Part 1: Single Ascending Dose

The primary objective of Part 1 is to assess the safety and tolerability of anti-SARS-CoV-2 IgY when given as single-ascending doses administered intranasally to healthy participants.

### Part 2: Multiple Dose

The primary objective of Part 2 is to assess the safety and tolerability of anti-SARS-CoV-2 IgY when given as multiple doses administered intranasally to healthy participants. A secondary objective is to assess the pharmacokinetics of anti-SARS-CoV-2 IgY when given as multiple doses administered intranasally to healthy participants.

Safety will be evaluated using adverse event (AE), physical examination (including vital signs), electrocardiogram, and clinical laboratory data. Pharmacokinetics will be evaluated by serum anti-SARS-CoV-2 IgY concentration.

## 3. INVESTIGATIONAL PLAN

### 3.1 Summary of Study Design

This is a single-center, randomized, double-blind, placebo-controlled phase 1 study of anti-SARS-CoV-2 IgY given intranasally to healthy participants at a single site in Australia. Participants will be recruited from the Linear Clinical Research Ltd panel or by direct advertising to the public.

<u>Part 1</u>: Participants will be randomly assigned to receive a single dose of anti-SARS CoV-2 IgY or placebo in a sequential escalating manner. It is planned that up to 3 groups will be dosed with 8 healthy participants per group (6 active and 2 placebo in each group). Each group in Part 1 will include the initial dosing of a sentinel group (1 anti-SARS-CoV-2 IgY and 1 placebo) at least 24 hours prior to dosing the remaining 6 subjects in the cohort (5 anti-SARS-CoV-2 IgY and 1 placebo). The remainder of the cohort will only be dosed if, in the opinion of the investigator, there are no significant safety concerns identified in the sentinel subjects within the first 24 hours after administration of the dose (anti-SARS-CoV-2 IgY or placebo). The following regimens are planned:

- Regimen A: 2 mg anti-SARS-CoV-2 IgY preparation (2 drops of 0.5 mg/100 µL/drop to each nostril) or placebo.
- Regimen B: 4 mg anti-SARS-CoV-2 IgY preparation (2 drops of 1 mg/100 µL/drop to each nostril) or placebo.
- Regimen C: 8 mg anti-SARS-CoV-2 IgY preparation (2 drops of 2 mg/100 µL/drop to each nostril) or placebo.

<u>Part 2</u>: Participants will be randomly assigned to receive multiple daily administrations of anti-SARS-CoV-2 IgY or placebo every 4 hours (3-times daily) for 14 days in a parallel-group manner. It is planned that up to 24 healthy participants will be randomised to 1 of 4 treatment regimens (6 participants per regimen). The following regimens are planned:

- Regimen A: 6 mg total daily dose anti-SARS-CoV-2 IgY preparation (2 drops of 0.5 mg/100 µL/drop to each nostril 3-times daily) for 14 days.
- Regimen B: 12 mg total daily dose anti-SARS-CoV-2 IgY preparation (2 drops of 1 mg/100 µL/drop to each nostril 3-times daily) for 14 days.
- Regimen C: 24 mg total daily dose anti-SARS-CoV-2 IgY preparation (2 drops of 2 mg/100 µL/drop to each nostril) for 14 days.
- Regimen D: 0 mg total daily dose placebo preparation (2 drops of 0 mg/100 µL/drop to each nostril) for 14 days

### 3.2 Outline of Visit Schedule

#### 3.2.1 Screening Evaluations

Screening assessments must be done to determine participant eligibility. Written consent must be obtained before conducting any study procedures.

##### 4.2.1.1 Part 1: Single-Ascending Dose Schedule of Events

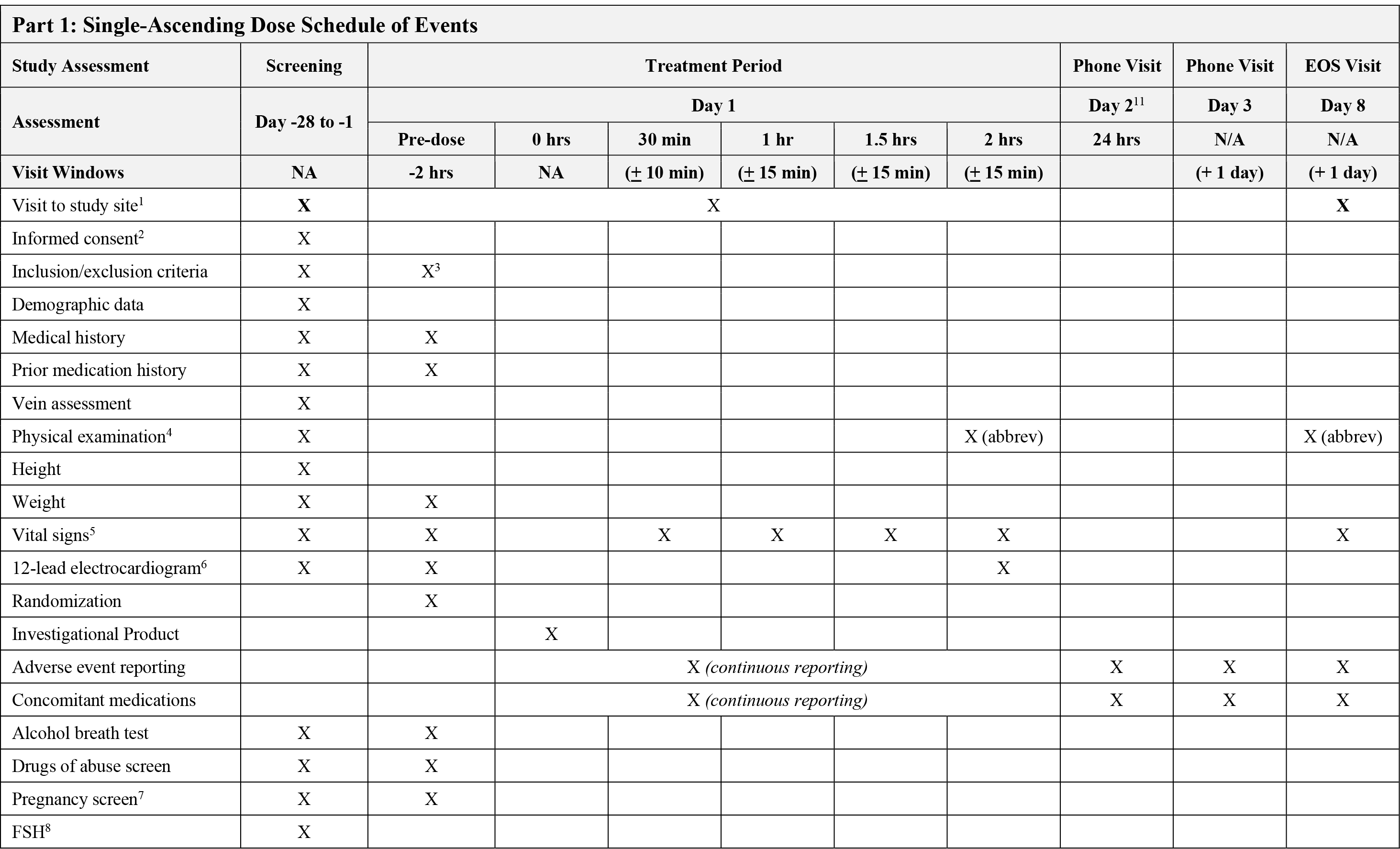
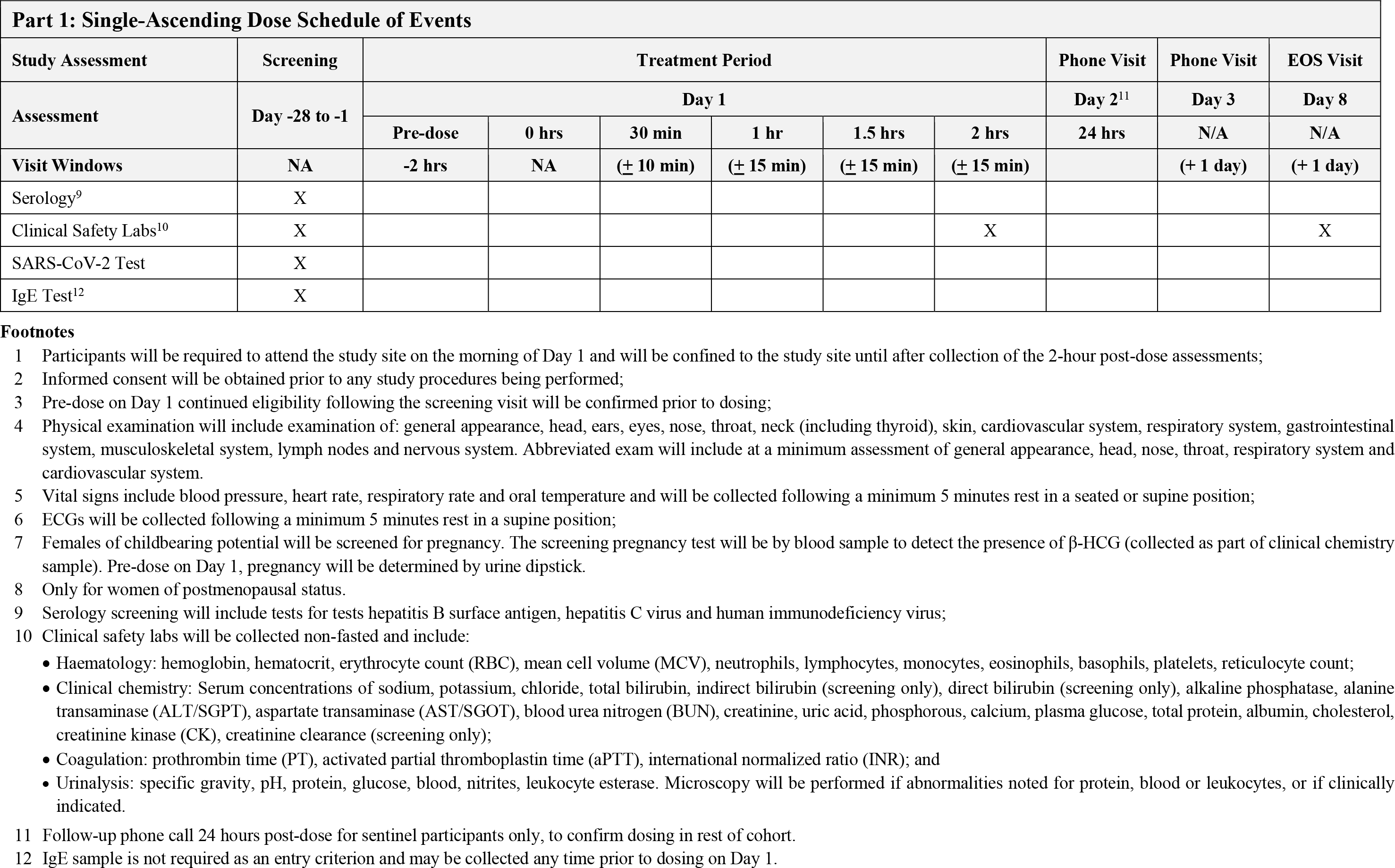

##### 4.2.1.2 Multiple Dose Schedule of Events

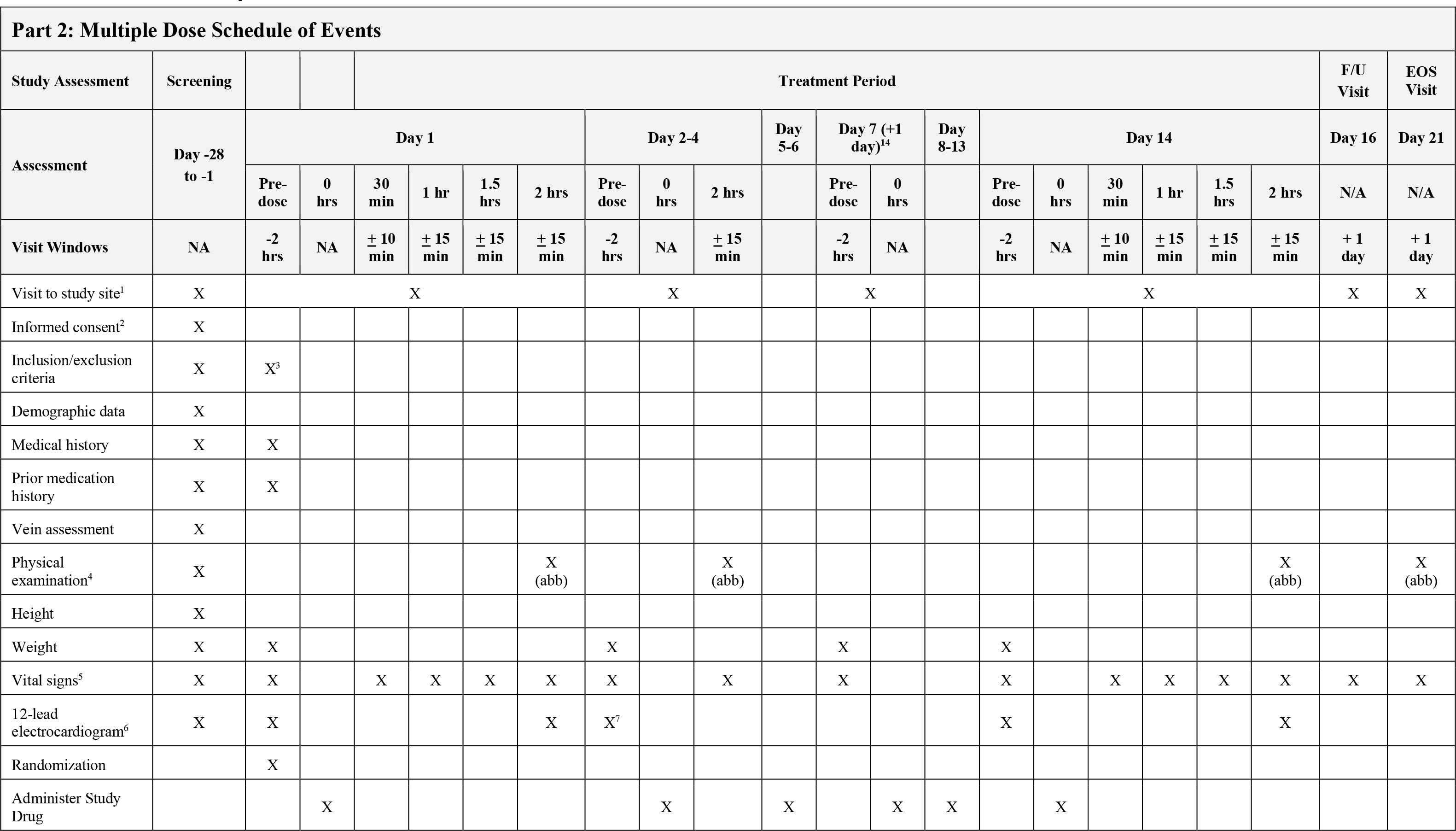
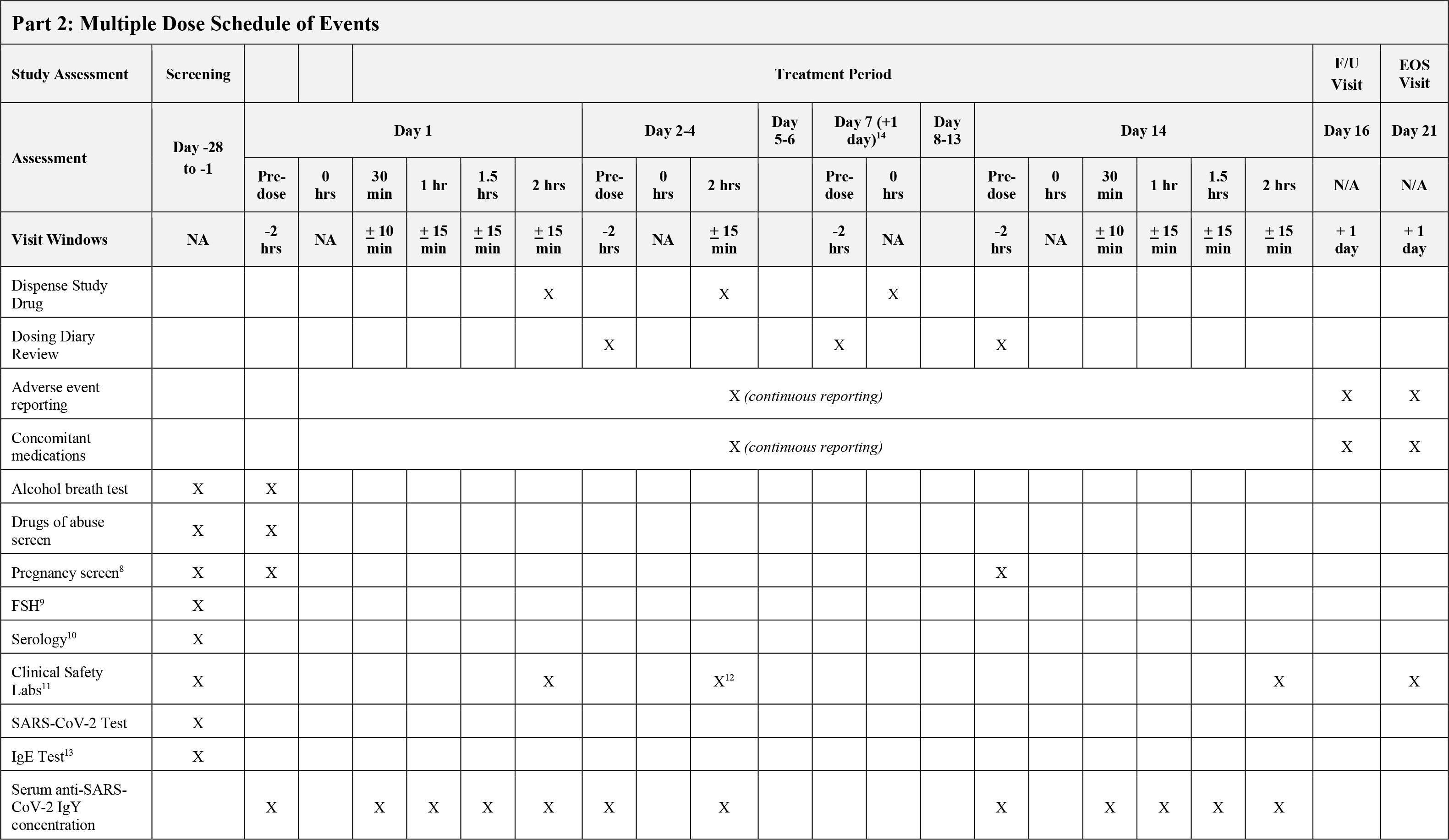
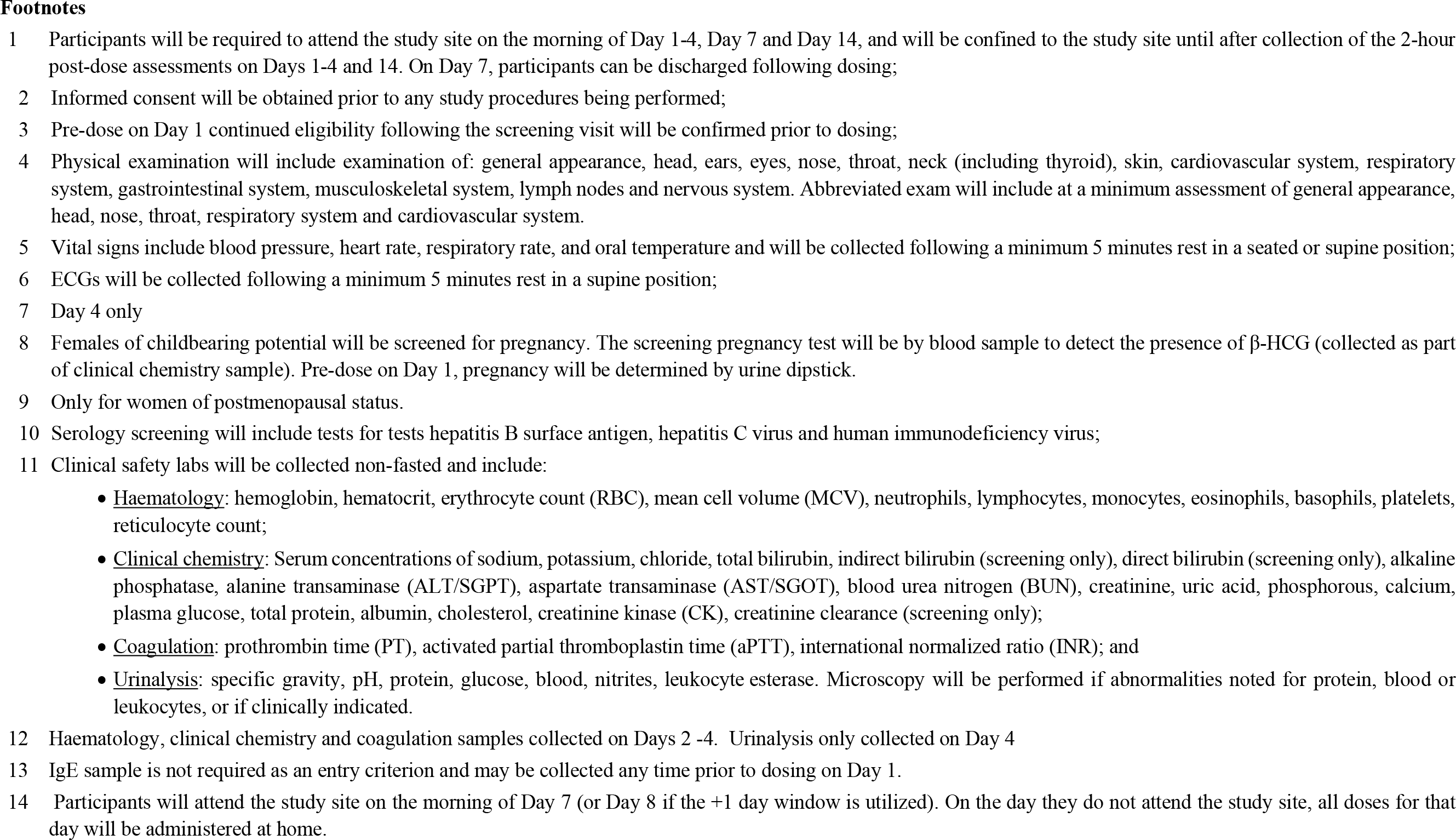

## 5 DISCUSSION OF DESIGN

This is a single-center, randomized, double-blind, placebo-controlled phase 1 study of anti-SARS-CoV-2 IgY given intranasally to healthy participants.

<u>Part 1</u>: Participants will be randomly assigned to receive a single dose of anti-SARS-CoV-2 IgY or placebo in a sequential escalating manner. It is planned that up to 3 groups will be dosed with 8 healthy participants per group (6 active and 2 placebo in each group).

<u>Part 2</u>: Participants will be randomly assigned to receive multiple daily administrations of anti-SARS-CoV-2 IgY antibodies or placebo every 4 hours (3-times daily) for 14 days in a parallel-group manner. It is planned that up to 24 healthy participants will be randomised to 1 of 4 treatment regimens (6 participants per regimen).

<u>Dose</u>: The maximum feasible solubility of IgY is approximately 20 mg/mL. The total maximum daily dose of anti-SARS-CoV-2 IgY to be used in the present study (24 mg daily) is based in part on solubility considerations and is less than the daily dose of anti- *P. aeruginosa* IgY given prophylactically as continuous oral treatment for up to 12 years (114 patient-years) to prevent pulmonary infections in 17 patients with cystic fibrosis (Nilsson et al., 2008). This study showed a significant reduction in *P. aeruginosa* compared with 23 control patients with cystic fibrosis, with no adverse events (Nilsson et al., 2008). Further, an equivalent of ½ egg/treatment was used/dose, which translates to about 50 mg IgY/dose given once daily (Dr. Anders Larsson, Uppsala University, Sweden, personal communication). As a result, the maximum daily dose in the planned Phase 1 study of 24 mg given intranasally to healthy volunteers for 14 days is significantly less than the oral daily dose given continuously to patients with cystic fibrosis over long periods. For the maximum anti-SARS-CoV-2 IgY dose of 4 mg/nare, we calculate a favorable ratio of IgY to viral particles, even when viral amount covers 100% of the nasal pathway and 100% of the mucosa thickness.

- <u>Entry Criteria</u>: Participants will be healthy adults and receive standard assessments to determine eligibility.
- <u>Safety Endpoints</u>: The safety assessments of physical examination, vital signs, clinical laboratories, electrocardiogram, and adverse event collection used in this trial are standard assessments for phase 1 studies evaluating the safety of single and repeated dosing.
- <u>Pharmacokinetic Endpoint</u>: Serum levels of anti-SARS-CoV-2 IgY will be measured by enzyme-linked immunosorbent assay to determine the presence of systemic exposure.

## 6 STUDY POPULATION

The investigators participating in this study have expertise in the conduct of phase 1 studies. The participants will comprise healthy adults between the ages of 18 and 45.

Participation in this study is voluntary. The nature of the study will be fully explained to each participant during the informed consent. The participants will have the opportunity to ask questions. An informed consent document will then be signed by the participant and the person performing the consent discussion and retained by the investigator according to Good Clinical Practice. A copy of the signed informed consent document will be given to the participant.

Eligibility for enrollment will be based on the results of screening for the following inclusion and exclusion criteria.

### 6.1 Inclusion & Exclusion Criteria Inclusion Criteria

- Healthy males or non-pregnant, non-lactating females aged 18 to 45 years.
- Body weight of at least 50 kg.
- Body mass index between ≥18.0 and ≤32.0 kg/m^2^. If outside this range, eligible if not considered clinically significant.
- Good state of health (mentally and physically) in the opinion of the investigator as indicated by a comprehensive clinical assessment (detailed medical history and a complete physical examination), electrocardiogram (ECG), and laboratory investigations (hematology, clinical chemistry, coagulation, and urinalysis).
- Female participants are eligible to participate if they are not pregnant, not breastfeeding, and at least 1 of the following conditions applies:

- Not of childbearing potential, defined as surgically sterile (hysterectomy, bilateral salpingectomy, tubal ligation or bilateral oophorectomy - verbal confirmation through medical history review acceptable) or postmenopausal (no menses for 12 months and confirmed by follicle-stimulating hormone (FSH) level ≥40 mlU/mL);
- Of childbearing potential and agrees to practice true abstinence or agrees to use a highly effective method of contraception consistently from 30 days prior to Day 1 until at least 30 days after dosing. Highly effective contraception includes hormonal contraception (oral, injected, implanted or transdermal) plus use of a condom, placement of an intrauterine device or intrauterine system plus use of a condom, or a vasectomised male partner (performed at least 6 months prior) who has been documented to no longer produce sperm - verbal confirmation through medical history review acceptable. Contraception requirements do not apply for participants in an exclusively same-sex relationship
- Male participants must agree to practice true abstinence; be surgically sterilised (performed at least 6 months prior and documented to no longer produce sperm - verbal confirmation through medical history review acceptable); or agree to use a condom plus effective contraception (i.e. established use of hormonal contraception - started at least 30 days prior to Day 1; or placement or an intrauterine device or intrauterine system) for their female partner, if of childbearing potential, from screening and for at least 90 days after dosing and refrain from donating sperm during this period. Contraception requirements do not apply for participants in an exclusively same-sex relationship
- Must provide written informed consent before any study procedure is performed.

#### Exclusion Criteria

1. Participants who have received any investigational drug in a clinical research study within the previous 30 days prior to screening or 5 half-lives, whichever is longer.
2. Participants who are study site employees, or immediate family members of a study site or sponsor employee.
3. History of any drug or alcohol abuse in the past 2 years defined as >21 units of alcohol per week for males and >14 units of alcohol per week for females. Where 1 unit = 360 mL of beer, 150 mL wine, or 45 mL of spirits.
4. Current smokers or users of e-cigarettes and nicotine replacement products and those who have used more than 3 of these products per month within the last 6 months.
5. Females of childbearing potential who are pregnant or lactating or planning to become pregnant during the study (female participants of childbearing potential must have a negative pregnancy test at screening and Day 1). A woman is considered of childbearing potential unless she is permanently sterile (hysterectomy, bilateral salpingectomy, and bilateral oophorectomy) or is postmenopausal (had no menses for 12 months without an alternative medical cause and a serum follicle-stimulating hormone [FSH] concentration ≥40 IU/L).
6. Participants who do not have suitable veins for multiple venipunctures/cannulation as assessed by the investigator at screening.
7. Clinically significant abnormal biochemistry, hematology, coagulation or urinalysis as judged by the investigator.
8. Positive drugs of abuse test result.
9. Evidence of renal impairment at screening, as indicated by an estimated creatinine clearance of <80 mL/min using the Cockcroft-Gault equation.
10. Abnormal liver function tests as indicated by alanine aminotransferase (ALT) > 1.5 x upper limit of normal (ULN), aspartate aminotransferase (AST) > 1.5 x ULN or total bilirubin >1.5 x ULN. Note: may be repeated once at the discretion of the investigator. Participants with Gilbert’s Syndrome may be enrolled at the discretion of the investigator.
11. Positive test result for hepatitis B, hepatitis C or human immunodeficiency virus (HIV) at Screening
12. Positive test result for SARS-CoV-2 by RT-PCR or positive SARS-CoV-2 serology result at Screening.
13. Evidence or history of clinically significant allergic (except for untreated, asymptomatic, seasonal allergies at time of dosing), hematological, endocrine, pulmonary, gastrointestinal, cardiovascular, hepatic, renal, psychiatric, or neurological disease.
14. Participants with a history of egg allergy.
15. History of nasal surgical procedures (e.g., turbinectomy, rhinoplasty, etc).
16. Frequent or recurrent nasal conditions, as determined by the investigator, or any nasal conditions within the 7 days prior to Day 1 (e..g., rhinitis, hay fever).
17. Current use of any nasal preparations (e.g. decongestants, steroids, etc).
18. Acute illness (gastrointestinal, infection [e.g., influenza] or known inflammatory process) at screening or Day 1.
19. Abnormal screening ECG, including QT intervals corrected with Fridericia’s formula >450 msec for males or >470 msec for females, or subject has any cardiac rhythm other than sinus rhythm that is interpreted by the investigator to be clinically significant.
20. Supine resting bradycardia (pulse heart rate [HR] <40 bpm) or a supine resting tachycardia (HR >100 bpm) at screening or Day 1.
21. Hypertension, defined as a supine resting systolic blood pressure >140 mm Hg or a supine resting diastolic blood pressure >90 mm Hg at screening or Day 1.
22. Hypotension, defined as a supine resting systolic blood pressure <90 mm Hg or a supine resting diastolic blood pressure <40 mm Hg at screening or Day 1
23. Known personal or family history of congenital long QT syndrome or known family history of sudden death.
24. Any condition possibly affecting drug absorption or elimination, such as previous surgery on the nose.
25. Donation or loss of greater than 400 mL of blood within the previous 3 months from screening.
26. Taking, or have taken, any prescribed or over-the-counter drug (other than acetaminophen, or hormone replacement therapy and hormonal contraception) or herbal remedies in the 14 days before screening. Exceptions may apply on a case-by-case basis, if considered not to interfere with the objectives of the study as agreed by the principal investigator and sponsor’s medical monitor.
27. Failure to satisfy the investigator of fitness to participate for any other reason.

### 6.2 Removal of Participants from Study Drug or Assessment

#### 6.2.1 Early Discontinuation of Study Drug

A participant must prematurely discontinue study drug under any of the following circumstances:

- The participant wishes to discontinue study drug.
- The investigator wishes the participant to discontinue study drug, especially but not limited to the investigator concluding that further treatment puts the participant at unacceptable risk.
- The participant develops a condition or begins a therapy that would have excluded entry into the study.
- The participant becomes pregnant during the study period. In this circumstance, the pregnancy must be immediately reported by telephone to the sponsor.
- The participant develops an adverse event with the Division of AIDS (DAIDS) Table for Grading the Severity of Adult and Pediatric Adverse Events that is grade 3 or greater toxicity and related to study drug in the judgment of the investigator.

#### 6.2.2 Participant Withdrawal from the Study

A participant must be withdrawn from the study (and discontinue any study drug) if:

- the participant requests such study discontinuation.

The reason for study withdrawal must be recorded in the participant’s case report form (CRF).

## 7 TREATMENTS

### 7.1 Participant Assignment

After signing the informed consent documents, participants who meet all eligibility criteria will be assigned to study drug assignment.

### 7.2 Method of Assignment to Treatment

Participants who have met all the inclusion criteria for enrollment and none of the exclusion criteria will be randomized to receive either anti-SARS-CoV-2 IgY or placebo, with dosage determined based on cohort assignment. Computer-generated random numbers will be used by the study statistician to generate the randomization allocation sequence. The randomization number assigned to the participant, which will be used to link the participant to the unique medication number for the bottle of study drug to be administered to the participant.

After signing the informed consent form, each participant will be given a screening number according to the screening order. On Day 1, following confirmation of eligibility, participants will be randomized to receive anti-SARS-CoV-2 IgY or placebo.

At the time of randomization, the participant will be assigned a unique randomization number, which will be allocated sequentially based on the pre-determined randomization schedule, and according to their chronological order of inclusion in the study. Confirmation of the treatment number allocated will be documented in the drug accountability records and recorded in the electronic case report form (eCRF).

Both the screening and randomization numbers will be used to identify the participant throughout the study period and on all study-related documentation.

### 7.3 Materials and Supplies

#### 7.3.1 Formulation, Packaging and Labeling

Study drug (anti-SARS-CoV-2 IgY or placebo) will be administered to the participant in a (personal use) bottle with a dropper (Bravado Pharma, Inc, Lutz, FL) through the site pharmacy. The isolation of IgY from individual eggs was performed using the protocols similar to the one described by Akita and Nakai (1993) and Fu et al (2012). Briefly, egg white was separated from the egg yolk and discarded. The yolk was mixed vigorously with sterilized water and left standing overnight at 4 °C after a slight acidification step. The mixture was then centrifuged at 1200×g for 30 minutes at 4 °C, and the supernatant was lyophilized. The protein concentration in harvested eluate was determined according to Bradford protein assay. Each egg yolk produced ∼35-40 mg of IgY-enriched preparation.

The IgY preparation was subjected to analysis by sodium dodecyl sulfate-polyacrylamide gel electrophoresis (SDS-PAGE) and Western blot. The isolated IgY had a high purity as confirmed by SDS-PAGE and purified IgY had good biological activity as confirmed by Western blot. The activity of IgY in yolks diluted at up to 1:128,000 in phosphate buffer saline was assessed using an indirect enzyme-linked immunosorbent assay as described previously (Huang et al., 2005). After immunization of specific-pathogen-free chickens with SARS coronavirus antigen, there were no detectable antibodies against SARS coronavirus in the yolks of eggs laid in the first 3 weeks after the first immunization. The production of anti-SARS antibody in yolks was found to commence 2 weeks after the IgY was produced in serum.

Drug substance (IgY preparation) is formulated with an inactive excipient (2% Carboxymethylcellulose Sodium), at neutral pH at ∼300mosm. Each bottle of drug product (anti-SARS-CoV-2 IgY preparation) and matched placebo will have liquid identical in appearance and smell.

**Table 7.1.** Study Drug Treatment and Packaging (Part 1, Single-Ascending Dose)

| **Treatment** | **Study Drug** | **Frequency** | **Packaging** |
| --- | --- | --- | --- |
| anti-SARS-CoV-2 IgY | anti-SARS-CoV-2 IgY preparation in liquid (0.5 mg/100 µL/drop, 1 mg/100 µL/drop, and 2 mg/100 µL/drop ) | 2 drops to each nostril as a single administration | Nose drop bottle |
| Placebo | liquid (100 µL/drop) | 2 drops to each nostril 3-times daily while awake (every 4 hours) | Nose drop bottle |

**Table 7.2.** Study Drug Treatment and Packaging (Part 2, Multiple Dose)

| **Treatment** | **Study Drug** | **Frequency** | **Packaging** |
| --- | --- | --- | --- |
| anti-SARS-CoV-2 IgY | anti-SARS-CoV-2 IgY preparation in liquid (0.5 mg/100 µL/drop, 1 mg/100 µL/drop, and 2 mg/100 µL/drop ) | 2 drops to each nostril every 4 hours (3-times daily) for 14 days | Nose drop bottle |
| Placebo | liquid (100 µL/drop) | 2 drops to each nostril every 4 hours (3-times daily) for 14 days | Nose drop bottle |

Study drug bottle will contain the following information:

- Protocol number (CVR001).
- Dose: Each vial contains 1.5 mL anti-SARS-CoV-2 IgY Preparation Nasal Suspension, 5, 10, or 20 mg/mL, or Placebo, for Intranasal application.
- Lot number (BP0001).
- Retest Date.
- Storage: 2 °C - 8 °C (36 °F - 46 °F).
- A precautionary statement that drug is limited to clinical trial use only.
- Keep out of reach of children.
- Company name (SPARK).
- Manufacturer: Bravado Pharmaceuticals, LLC, 44212 Cypress Gulch Drive, Lutz, FL 33559.

#### 7.3.2 Storage and Handling

The study drug will be in bottles. The study drug will be stored at 2 °C - 8 °C (36 °F - 46 °F). No other special handling is required.

#### 7.3.3 Final Disposition of Clinical Supplies

At the end of the study, study drug supply and accountability records will be reconciled as to drug shipped, drug consumed, and drug remaining. Any discrepancies noted will be documented. Final drug accountability reconciliation will be performed at the visit occurring at the end of treatment or early discontinuation. Unused study drug will be destroyed.

### 7.4 Dosage Administration

The investigator (or designee) will administer a single dose of study drug in Part 1 and the morning dose on Days 1-4, Day 7 and Day 14 in Part 2. All other doses in Part 2 will be self-administered by the participant. Participants will be instructed with the following language:

“Gently blow your nose before using this drug. Then tilt your head back while sitting or lying down. After the study drug is administered, keep your head tilted for a few minutes. Try not to blow your nose for at least 5 minutes after study drug administration”

In Part 2, participants will be dispensed study drug in a dropper bottle to complete the subsequent doses at home, 4 hours apart for a total of 3 doses per day. They will retain the empty bottles to return to the site at the following visit for accountability. Doses performed at home will be recorded in a dosing diary. On Day 14 there will only be a single morning dose.

The investigator (or designee) is responsible for the correct use of the study drug to the participants, confirming that instructions are followed properly, maintaining accurate records of study drug dispensing, and collection of all unused study drug, including empty drug packaging.

Participants will be instructed to contact the investigator as soon as possible if he or she has a complaint or problem with the study drug or drug delivery system so that the situation can be assessed.

### 7.5 Blinding

This is a randomized, double-blind, placebo-controlled study. Neither the participants nor the study personnel with direct contact with the participant will know which study drug (anti-SARS-CoV-2 IgY vs. placebo) is being administered during the treatment period. Placebo and active drug liquid for intranasal administration will be similar in appearance and smell, and the amount of study drug will be the same for each treatment group.

Emergency unblinding for AEs will be performed through the code break envelopes. Emergency unblinding for AEs may be used only if a participant’s medical care requires knowledge of the participant’s treatment assignment. The investigator should make every effort to contact the sponsor before unblinding a participant’s treatment assignment. If a participant’s treatment assignment is unblinded, the sponsor must be immediately notified by telephone.

### 7.6 Concomitant Therapy

Nasal treatments that may interfere with administration of study drug in the judgment of the investigator are prohibited during the study.

### 7.7 Dosing Diary

Participants will be issued a dosing diary to record dosing times for treatment compliance. Participants will be instructed to bring their diary with them during all visits to the clinic.

## 8 ADVERSE EVENT REPORTING

### 8.1 Definition of Adverse Event

For purposes of this trial, an AE will be defined as **any** new unfavorable or unintended sign, symptom, or disease or change of an existing condition, which occurs during or after treatment, whether or not considered treatment-related. If clinically significant laboratory values lead to or are associated with clinical symptoms(s), the diagnosis should be reported as an adverse event. Lack of drug effect is not an adverse event in clinical trials because the purpose of the clinical trial is to establish drug effect.

Before enrollment, study site personnel will note the occurrence and nature of each participant’s medical condition(s) in the appropriate section of the CRF. During the remainder of the study, site personnel will again note any change in the condition(s) and the occurrence and nature of any adverse events.

If the study drug is discontinued for a participant, study site personnel must report and clearly document the circumstances and data leading to any such discontinuation, using designated case report forms. For adverse events, the participant should be followed until the event resolves or stabilizes, with frequency of follow-up at the discretion of the investigator.

In cases where the investigator notices an unanticipated benefit to the participant, study site personnel should enter “unexpected benefit” with the actual event term (for example, the complete actual term would be “unexpected benefit—sleeping longer”).

Cases of pregnancy that occur during maternal or paternal exposures to study drug should be reported for tracking purposes. Data on fetal outcome and breastfeeding are collected for regulatory reporting and drug safety evaluation.

#### 8.1.1 Reporting Procedures for All Adverse Events

Investigators are responsible for monitoring the safety of participants who have entered this study and noting any event that seems unusual, even if this event may be considered an unanticipated benefit to the participant. The investigator is responsible for appropriate medical care of participants during the study.

The investigator remains responsible for following, through an appropriate health care option, adverse events that are serious or that caused the participant to discontinue the study. The participant should be followed until the event is resolved or explained. Frequency of follow-up is left to the discretion of the investigator.

Adverse event information will be collected through Day 8 (Part 1) and Day 21 (Part 2). Participants who discontinue study drug at any time will have adverse events collected through an additional 1 week, *provided consent to continue in the study has not been withdrawn*.

The investigator is responsible for assessing and recording all adverse experiences. Each AE will be recorded and classified for intensity, seriousness, and causality. All AEs either observed by the investigator or reported by the participant will be recorded regardless of causality. The investigator will follow the participant until an AE resolves or stabilizes.

#### 8.1.2 Adverse Event Severity

The Division of AIDS (DAIDS) Table for Grading the Severity of Adult and Pediatric Adverse Events version 2.1 (July 2017) will be used to assess and grade AE severity, including laboratory abnormalities judged to be clinically significant. If the experience is not covered in the DAIDS criteria, the following guidelines should be used to grade severity:

− Mild (grade 1): Mild symptoms causing no or minimal interference with usual social and functional activities with intervention not indicated.

− Moderate (grade 2): Moderate symptoms causing greater than minimal interference with usual social and functional activities with intervention indicate.

− Severe (grade 3): Severe symptoms causing inability to perform usual social and functional activities with intervention or hospitalization indicated.

− Life-threatening (grade 4): Potentially life-threatening symptoms causing inability to perform basic self-care functions with intervention indicated to prevent permanent impairment, persistent disability, or death.

The term “severe” is a measure of intensity and a severe AE is not necessarily serious.

#### 8.1.3 Adverse Event Relationship to Study Drug

The relationship of an AE to the study drug should be based on the judgment of the investigator and assessed using the following guidelines:

− Definitely: Previously known toxicity of agent; or an event that follows a reasonable temporal sequence from administration of the drug; that follows a known or expected response pattern to the suspected drug; that is confirmed by stopping or reducing the dosage of the drug; and that is not explained by any other reasonable hypothesis.

− Probably: An event that follows a reasonable temporal sequence from administration of the drug; that follows a known or expected response pattern to the suspected drug; that is confirmed by stopping or reducing the dosage of the drug; and that is unlikely to be explained by the known characteristics of the participant’s clinical state or by other interventions.

− Possibly: An event that follows a reasonable temporal sequence from administration of the drug; that follows a known or expected response pattern to that suspected drug; but that could readily have been produced by a number of other factors.

− Unrelated: An event that can be determined with certainty to have no relationship to the study drug.

#### 8.1.4 Serious Adverse Event Definition and Reporting Procedures

Any AE that meets the definition of serious noted below and occurs in a participant during the course of the study must be reported to the sponsor by telephone within 24 hours of the investigator becoming aware of the event. In addition, a serious adverse event form (SAE) must be completed by the investigator and faxed to the study sponsor within 24 hours of the investigator becoming aware of the event. In addition, the site investigator must report SAEs to their local Ethics Committee (EC) in accordance with the EC’s standard operating procedures and policies.

An SAE is defined as an adverse event that suggests a significant hazard or side effect, regardless of the relationship to study drug. An SAE includes, but may not be limited to, any event that:

- Results in death.
- Is life-threatening. This definition implies that the participant, in the view of the investigator, is at immediate risk of death from the event. It does not include an event that, had it occurred in a more serious form, might have caused death.
- Requires inpatient hospitalization or prolongs existing hospitalization.
- Results in persistent or significant disability/incapacity.
- Results in a congenital anomaly or birth defect. This serious criterion applies if a participant exposed to an investigational product gives birth to a child with a congenital anomaly or birth defect.

Medical and scientific judgment will be exercised in deciding whether classification of an AE as serious is appropriate in other situations, such as important medical events that may not be immediately life-threatening or result in death or hospitalization, but may jeopardize the participant or require intervention to prevent one of the outcomes listed in the definition above. These should also usually be considered serious. Examples of such events include allergic bronchospasm requiring intensive treatment in an emergency room or at home, blood dyscrasias or convulsions that do not result in inpatient hospitalization, or the development of drug dependence or abuse.

Serious adverse events occurring after a participant is discontinued from the study will only be reported if the investigator believes that the event may have been caused by the study drug or a protocol procedure.

For the purpose of expedited reporting to regulatory agencies, an investigator will be responsible for identifying any adverse event that is serious, unexpected, and believed to be related to study drug. An adverse event or suspected adverse reaction is considered

“unexpected” if it is not consistent with the risk information described in the general investigational plan or elsewhere in the current investigational new drug application. For example, under this definition, hepatic necrosis would be unexpected (by virtue of greater severity) if the investigator brochure or investigational drug application referred only to elevated hepatic enzymes or hepatitis. Similarly, cerebral thromboembolism and cerebral vasculitis would be unexpected (by virtue of greater specificity) if the investigator brochure or investigational drug application listed only “cerebral vascular accidents.

#### 8.1.5 Laboratory Tests

Clinical laboratory tests will be performed at the times specified in the Study Schedule (see Section 4.2). All clinical laboratory assessments will be analyzed at the participating site’s local laboratory. Investigators must document their review of each laboratory report by signing or initialing and dating each report.

#### 8.1.6 Safety Monitoring

The sponsor or designee will monitor blinded safety data throughout the study. The sponsor (or designee) will review SAEs within time frames mandated by regulatory requirements and will review trends and adverse events at periodic intervals.

Prior to dose escalation in the SAD cohorts, the Safety Monitoring Committee (SMC) will review all available safety and tolerability data for a minimum of 7 participants who have completed the planned safety assessments up to 48 hours after dosing. The SMC will be composed of the independent medical monitor, principal investigator, and Sponsor’s medical representative. Other members of the investigational team may join the SMC as deemed appropriate.

The data will be reviewed blinded, unless the SMC considers it necessary to unblind the data for safety concerns. Before breaking the code per standard procedures, the potential decisions and actions will be determined.

SMC decisions on dose escalation will be taken in consensus between the members of the Safety Monitoring Committee. If consensus cannot be reached, the investigator, who has the ultimate responsibility for the safety of participants, will make the final decision on whether to continue or stop the study. The SMC’s decisions and their rationale will be documented.

## 9 QUALITY CONTROL AND QUALITY ASSURANCE

The investigator agrees to be responsible for implementing and maintaining quality control and quality assurance systems to ensure that trials are conducted and data are generated, documented, and reported in compliance with the protocol, accepted standards of Good Clinical Practice, and all applicable federal, state, and local laws, rules and regulations relating to the conduct of the clinical study.

The investigator also agrees to conduct the study in an efficient and diligent manner and in conformance with this protocol; generally accepted standard of Good Clinical Practice; and all applicable federal, state, and local laws, rules, and regulations relating to the conduct of the clinical study.

The investigator must allow study-related monitoring, audits, and inspection by the EC, sponsor (or designee), government regulatory agencies, and, if applicable, University compliance and quality assurance groups of all trial-related documents and procedures.

The investigator shall prepare and maintain accurate study documentation in compliance with Good Clinical Practice standards and applicable federal, state, and local laws, rules, and regulations.

## 10 DATA ANALYSIS METHODS

### 10.1 Determination of Sample Size

No formal sample size calculation was done. Based on experience from previous similar studies, the target number of participants to be enrolled is appropriate for the assessment of safety and tolerability.

### 10.2 Safety Variables

Safety evaluations will include collection of AE, physical examination, vital sign, electrocardiogram, and clinical laboratory data. Laboratory data will include evaluation of:

- Hematology: hemoglobin, hematocrit, erythrocyte count, mean cell volume, neutrophils, lymphocytes, monocytes, eosinophils, basophils, platelets, reticulocyte count.
- Metabolic panel: Serum concentrations of sodium, potassium, chloride, total bilirubin, alkaline phosphatase, alanine transaminase (ALT), aspartate transaminase (AST), blood urea nitrogen (BUN), creatinine, uric acid, phosphorous, calcium, plasma glucose, total protein, albumin, cholesterol, creatinine kinase.
- Coagulation: prothrombin time (PT), activated partial thromboplastin time (aPTT), international normalized ratio (INR).
- Urinalysis: specific gravity, pH, protein, glucose, blood, nitrites, leukocyte esterase. Microscopy will be performed if abnormalities noted for protein, blood or leukocytes, or if clinically indicated.
- Urine human chorionic gonadotropin (hCG) test.

### 10.3 Pharmacokinetic Variable

Serum anti-SARS-CoV-2 IgY concentration.

### 10.4 Immunoglobulin E

Immunoglobulin E (IgE) blood samples will be collected from each participant. Samples will be stored and may be used for future allergen specific IgE testing.

### 10.5 Statistical and Analytical Plans

#### 10.5.1 General Considerations

The analysis of safety variables will include all participants who receive study drug.

All variables will be summarized by descriptive statistics for each treatment group. The statistics for continuous variables will include mean, median, standard deviation, and number of observations. Categorical variables will be tabulated using frequencies and percentages. Where confidence intervals are presented, they will be two-sided 95% confidence intervals.

#### 10.5.2 Handling of Missing Data

Missing data will not be imputed for safety analyses.

#### 10.5.3 Participant Disposition

Study participant disposition will be summarized by treatment group. Participants who discontinued study drug prematurely or withdrew from the study will be summarized and listed, with reason for early termination/withdrawal.

#### 10.5.4 Participant Characteristics

Demographic and other baseline characteristics will be summarized by treatment group.

#### 10.5.5 Treatment Compliance

Treatment compliance will be evaluated. This will include the date the study drug was dispensed and the final amount of liquid returned.

#### 10.5.6 Safety Analyses

Treatment-emergent AEs are defined as AEs occurring after the first dose of study drug until Day 8 in Part 1 and Day 21 in Part 2. Duration of treatment will be summarized by treatment group.

The incidence of all reported AEs and treatment-related AEs will be tabulated by treatment group. Adverse events will be classified by system organ class and preferred term using the Medical Dictionary for Regulatory Activities (MedDRA).

Adverse events will be listed and summarized by treatment group, MedDRA preferred term, severity, seriousness, and relationship to study drug. In the event of multiple occurrences of the same AE with the same preferred term in one participant, the AE will be counted once as the worst occurrence. The incidence of AEs will be tabulated by system organ class and treatment group. AEs leading to premature discontinuation of study drug or withdrawal from the study will be summarized and listed in the same manner.

Summary statistics for actual values and for change from baseline will be summarized for laboratory results by treatment group and scheduled visit. Participants with laboratory values outside of the normal reference range at any post baseline assessment will be identified.

Data to be listed by subject and summarized by treatment will include AEs, vital signs, ECG parameters, and clinical laboratory evaluations. All values outside the clinical reference ranges will be flagged on the data listings. Other data to be listed by subject will include physical examination findings and concomitant medications.

#### 10.5.7 Pharmacokinetic Analysis

Summary statistics for actual values and for change from baseline will be summarized by treatment group and scheduled visit.

#### 10.5.8 Interim Analyses

No interim analyses are planned for this study.

## 11 ADMINISTRATIVE, ETHICAL, AND REGULATORY CONSIDERATIONS

The investigator is responsible for presenting the risks and benefits of study participation to the participant in simple terms using the informed consent document. The investigator will ensure that written informed consent is obtained from each participant by obtaining the appropriate signatures and dates on the informed consent document before the performance of protocol evaluations or procedures.

### 11.1 Ethical Review

The investigator will obtain documentation of the EC approval of the protocol and the informed consent document before the study may begin at the investigative site(s). The name and address of the reviewing EC are provided in the investigator file.

The sponsor will supply the following to the investigative site’s EC:

- Protocol and amendments.
- Informed consent document and updates.
- Investigational New Drug Application.
- Relevant curricula vitae, if required.
- Required safety and SAE reports.
- Any additional submissions required by the site’s EC.

The investigator must provide the following documentation to the sponsor or its designee:

- The EC periodic (e.g., quarterly, annual) reapproval of the protocol.
- The EC approvals of any amendments to the protocol or revisions to the informed consent document.
- The EC receipt of safety and SAE reports, as appropriate.

### 11.2 Regulatory Considerations

This study will be conducted in accordance with the protocol and ethical principles stated in the 2013 version of the Declaration of Helsinki or the applicable guidelines on Good Clinical Practice, and all applicable federal, state, and local laws, rules, and regulations.

All data recorded in the CRF for participants participating in this study will be transcribed from medical records. After reading the protocol, the investigator will sign the protocol signature page and return it to the sponsor or designee.

#### 11.2.1 Investigator Information

The contact information and qualifications of the principal investigator and sub-investigators, and name and address of the research facilities are included in the investigator file.

#### 11.2.2 Protocol Amendments and Study Termination

The sponsor will initiate changes to the protocol as necessary (except for changes to eliminate an immediate hazard to a study participant) and seek approval by the EC before implementing. The investigator is responsible for enrolling participants who have met protocol eligibility criteria. Protocol violations must be reported to the local EC in accordance with EC policies.

The sponsor may terminate the study at any time. The EC must be advised in writing of study completion or early termination.

#### 11.2.3 Study Documentation, Privacy, and Records Retention

Government agency regulations and directives require that all study documentation pertaining to the conduct of a clinical trial must be retained by the investigator for a minimum of 5 years (or longer if required by the local EC).

To protect the safety of participants in the study and to ensure accurate, complete, and reliable data, the investigator will keep records of laboratory tests, clinical notes, and participant medical records in the participant files as original source documents for the study. If requested, the investigator will provide the applicable regulatory agencies and applicable EC with direct access to original source documents.

Records containing participant medical information must be handled in accordance with the requirements of the Health Insurance Portability and Accountability Act (HIPAA) Privacy Rule and consistent with the terms of the participant authorization contained in the informed consent document for the study. Care should be taken to ensure that such records are not shared with any person or for any purpose not contemplated by the informed consent document. Furthermore, CRFs and other documents should be completed in strict accordance with the instructions provided by the sponsor, including the instructions regarding the coding of participant identities.

### 11.3 Study Finances

This study is financed through Linear Clinical Research Ltd and SPARK at Stanford.

### 11.4 Publications

Neither the complete nor any part of the results of the study carried out under this protocol, nor any of the information provided by the sponsor for the purposes of performing the study, will be published or passed on to any third party without the consent of the study sponsor. Any investigator involved with this study is obligated to provide the sponsor with complete test results and all data derived from the study.

---

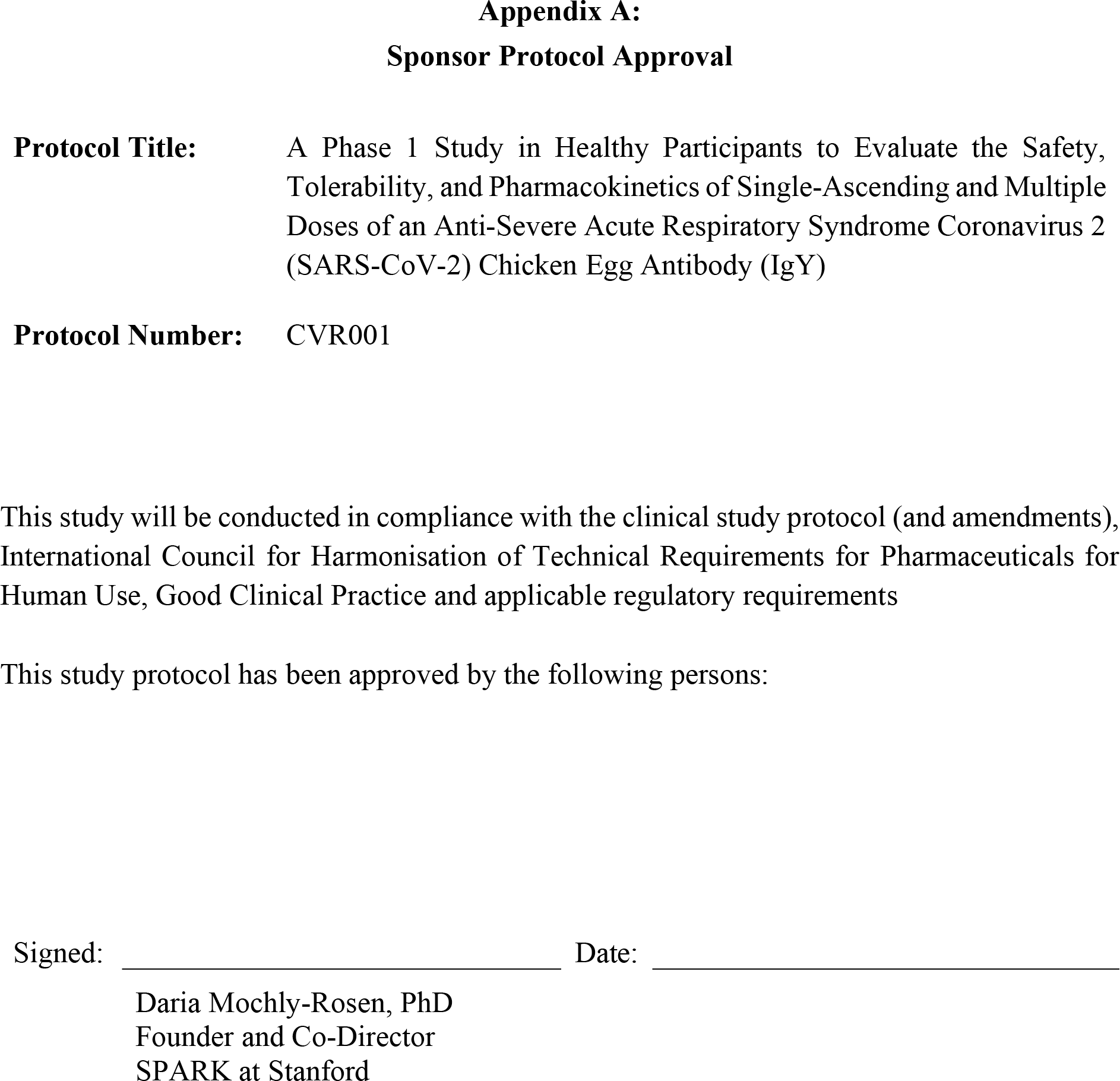
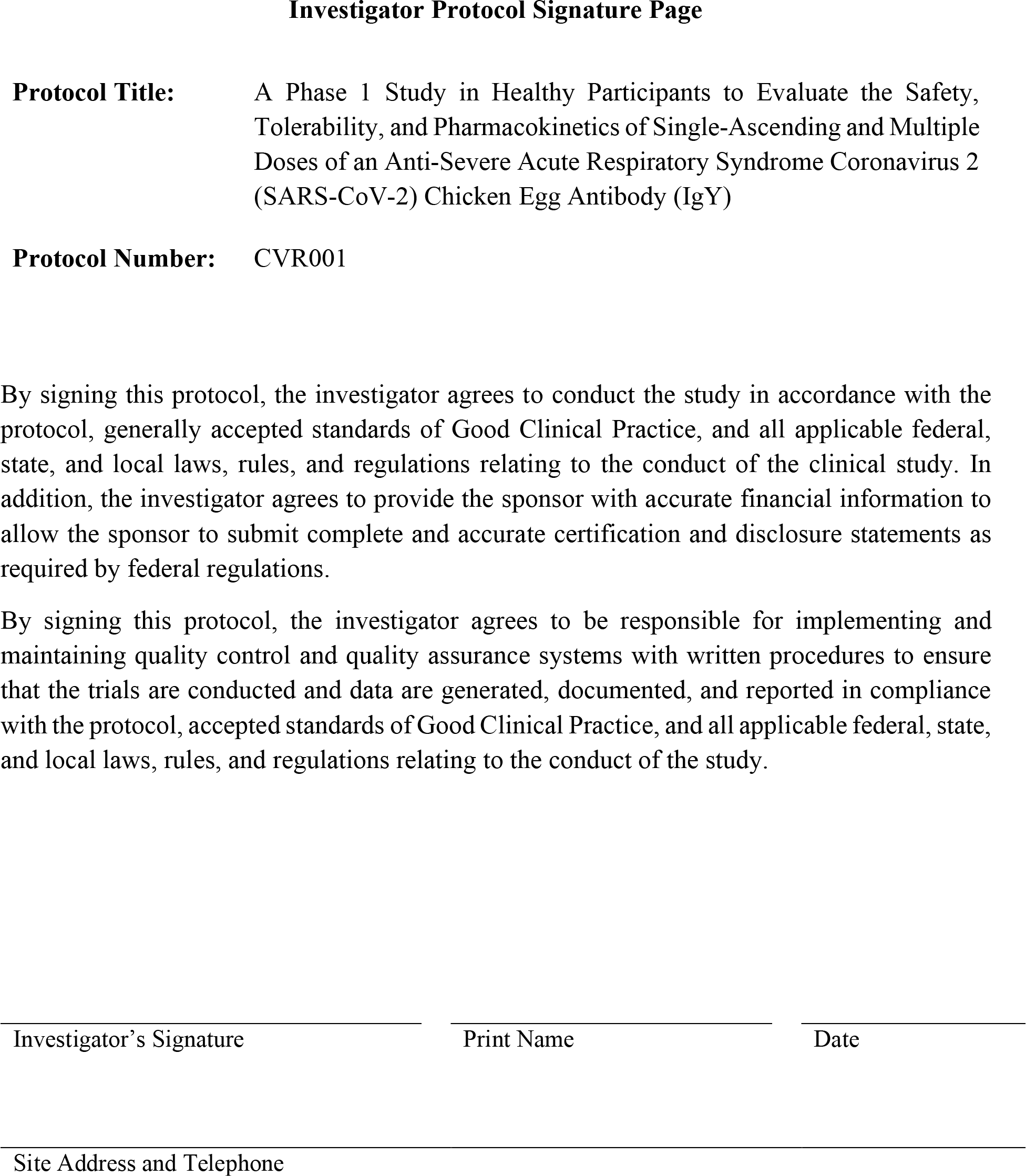

